# Trends of use of drugs with suggested shortages and their alternatives across 52 real world data sources and 18 countries in Europe and North America

**DOI:** 10.1101/2024.08.28.24312695

**Authors:** Marta Pineda-Moncusí, Alexandros Rekkas, Álvaro Martínez Pérez, Angela Leis, Carlos Lopez Gomez, Eric Fey, Erwin Bruninx, Filip Maljković, Francisco Sánchez-Sáez, Jordi Rodeiro, Loretta Zsuzsa Kiss, Michael Franz, Miguel-Angel Mayer, Neva Eleangovan, Pericàs Pulido Pau, Pantelis Natsiavas, Selçuk Şen, Steven Cooper, Sulev Reisberg, Katrin Manlik, Beatriz del Pino, Albert Prats Uribe, Ali Yağız Üresin, Ana Danilović Bastić, Ana Maria Rodrigues, Ãngela Afonso, Anna Palomar-Cros, Annelies Verbiest, Antonella Delmestri, Barış Erdoğan, Carina Dinkel-Keuthage, Carmen Olga Torre, Caroline de Beukelaar, Caroline Eteve-Pitsaer, Cátia F. Gonçalves, Costantino de Palma, Cristina Gavina, Daniel Dedman, David Brendan Price, Denisa Gabriela Balan, Dirk Enders, Edward Burn, Elisa Henke, Elyne Scheurwegs, Emma Callewaert, Encarnación Pérez Martínez, Eng Hooi Tan, Fabian Prasser, Francois Antonini, Frank Staelens, Fredrik Nyberg, Geoffray Agard, Gianluigi Galli, Gianmario Candore, Gianny Mestdach, Hadas Shachaf, Harri Rantala, Huiqi Li, Ines Reinecke, Irene López-Sánchez, Jaime E Poquet-Jornet, Javier de la Cruz Bertolo, Jelle Evers, João Firmino-Machado, Jonas Wastesson, Juan Luis Cruz Bermúdez, Juan Manuel Ramírez-Anguita, Kimmo Porkka, Kristina Johnell, Laurent Boyer, Lieselot Cool, Luca Moscetti, Manon Merkelbach, Mariana Canelas-Pais, Massimo Dominici, Máté Szilcz, Matteo Puntoni, Mees Mosseveld, Mina Tadrous, Miquel Oltra-Sastre, Mona Bové, Nadav Rappoport, Noelia García Barrio, Otto Ettala, Paolo Baili, Paula Rubio Mayo, Peter Prinsen, Raeleesha Norris, Ravinder Claire, Reut Sherman Yackob, Roberto Lillini, Salvador Garcia-Torrens, Sampo Kukkurainen, Silvia Lazzarelli, Talita Duarte-Salles, Tiago Taveira-Gomes, Tim Jansen, Ulrich Keilholz, Wai Yi Man, Xintong Li, Zsolt Bagyura, Daniel Prieto-Alhambra, Peter R. Rijnbeek, Theresa Burkard

**Author notes:** **Corresponding author** Prof Daniel Prieto-Alhambra, Pharmaco- and Device Epidemiology Group, Health Data Sciences, Botnar Research Centre, NDORMS, University of Oxford, Oxford, UK.

## Abstract

**Importance:** Drug shortages leave affected patients in a vulnerable position.

**Objective:** To describe incidence and prevalence of use for medicines with suggested shortages in at least one European country, as announced by the European Medicines Agency, and to characterise the users of these drugs including the indication of use, duration of use, and dosage.

**Design:** We performed a descriptive cohort study from 2010 and up to 2024 in a network of databases which have mapped their data to the Observational Medical Outcomes Partnership (OMOP) Common Data Model (CDM).

**Setting:** Settings included primary care, secondary care, claims and various disease registries.

**Participants:** We included all patients with at least 365 days of history on the database.

**Exposures:** All medicines with a suggested shortage in at least one European country for more than 365 days (n=18). We also assessed their key alternatives (n=39).

**Main outcomes and measures:** We estimated annual incidence rates and period prevalence. A drop in incidence or prevalence of >33% after the shortage was announced was considered confirmation of a shortage.

**Results:** Among 52 databases from Europe and the United States, we observed shortages according to decreased incidence of use for 8 drugs and shortages according to prevalence of use for 9 drugs. The drugs varenicline and amoxicillin alone or plus clavulanate were in shortage in the most number of countries.

**Conclusion and relevance:** We compiled and analysed data of annual incidence and prevalence of use plus information on patient characteristics, indication, and dose for 57 medicines among 52 databases in Europe and the United States between 2010 and 2024. We detected shortages and observed a change in the users’ characteristics for several drugs.

We have described timely real-world scenarios of drug shortages and those unobserved in various health care settings and countries which helps to better understand how drug shortages play out in real life.

## Introduction

Drug shortages represent an important global health issue, posing clinical and economic threats to healthcare systems and public health worldwide.^1^ Drug shortages have been identified as a key public health risk since the beginning of the 20^th^ century (e.g., the shortage of insulin in the early 1920s).^2^ A recent example is the fact that several countries in the Northern Hemisphere faced shortages of “basic medicines” such as antibiotics and painkillers for the treatment of respiratory tract infections, including paediatric formulations, during 2022 and 2023 winters.^3, 4^ The World Health Organization (WHO) highlighted the importance of drug shortages as a complex global challenge^5^ and urged the Member States to develop strategies to mitigate their impact.^6^

The causes of drug shortages can differ between countries and regions due to differences regarding health system structure, procurement processes, logistics etc. Therefore, different health authorities, agencies, and governments have developed various strategies, policies, action plans, and guidelines to address drug shortages and mitigate their potential impact. In 2013, the European Medicines Agency (EMA) launched the first catalogue of medicine shortages assessed by the Agency.^7^ Since that date, the EMA has regularly updated this catalogue in line with the current status of medicines under surveillance due to shortages in more than one European country, except for a suspension between March 2020 and December 2021 that was temporarily suspended due to the COVID-19 pandemic. The list includes information such as the reason for the shortage, and recommendations regarding alternatives for patients, healthcare professionals and other stakeholders.

In addition to anxiety, drug shortages can lead to significant harm, including treatment failure, delays in care, suboptimal treatment outcomes, and therapy errors, particularly when alternative medications are unavailable or unfamiliar. Furthermore, the treatment with alternative and potentially suboptimal medications may increase the risk of adverse reactions and/or potential drug interactions.^8^ Despite increased awareness about the importance of drug shortages and announcements of new recommendations/mitigation strategies, broad research directly focused on drug shortages is very limited. The topic is mainly evaluated by reviews and consensus statements. To this end, Real World Data (RWD) can be a powerful research accelerator to improve the understanding of drug shortages in the clinical practice, providing timely and accurate information on the prevalence and incidence of medication use over time, as well as an overview of the characteristics of the users.

The presented study uses a wide network of databases across Europe available in OMOP-CDM (the Observational Medical Outcomes Partnership Common Data Model) format. The European Health Data & Evidence Network (EHDEN, https://www.ehden.eu/) funded and supported the harmonisation of more than 187 European data bases covering over 200 million people in OMOP-CDM. ^9, 10^ After an open call for participation, we analysed 52 real world data sources from 18 countries: Belgium, Bulgaria, Estonia, Finland, France, Germany, Greece, Hungary, Italy, Netherlands, Portugal, Romania, Serbia, Spain, Sweden, Türkiye, UK, and the USA.

It should be noted that the medicines evaluated in the present study were already reported in the existing catalogue of EMA on drug shortages. In this context, this study did not aim to verify whether the drugs reported by the health authority were actually in shortage, but rather to evaluate the reported drug shortages from a different perspective using RWD. Identification of drug shortages according to the incidence/prevalence of usage of respective medicines, the significant changes in the use of respective medicines throughout the study period, reflection of reported drug shortages on RWD (mostly prescription data, not dispensing), changes in indication of use to review the contribution of high or low demand to the shortage, the change in a specific time period such as COVID-19 pandemic, stratification of the study population according to gender and different age groups, and differences between primary and secondary care settings can be listed as key points that were assessed in the present study. To the best of our knowledge, the present work is the first and the largest study using RWD to evaluate the drug shortages in the context of both incidence/prevalence at population-level and drug utilization characteristics at patient-level on real-world data from several European and some other countries.

## Methods

### Study design

We conducted a large-scale observational, retrospective cohort study consisting of two consecutive parts:

1. A population-level cohort study to assess the incidence and prevalence of use for 16 medicines with reported drug shortages in Europe and 39 of their alternatives.
2. A patient-level cohort of new and prevalent users to characterise drug utilisation of those medicines with reported drug shortages in Europe including their indications, duration of use, and dosage.

### Data Sources

We used routinely collected, longitudinal observational healthcare data from data sources standardized to the OMOP CDM. The study was open to all data partners in the EHDEN network, including EHDEN consortium members.

In this large network cohort study, 82 EHDEN data partners, accounting for 90 databases, expressed interest to join; 67 databases attempted the feasibility stage, of which 63 databases passed, whilst one did not pass feasibility because they did not have any drug of interest, and three could not execute the code due to compatibility problems between the study package and the database management system. Finally, two of the databases that passed the feasibility withdrew at that stage, and one was also available from another data partner and therefore excluded. Of the remaining 60 databases, 54 successfully ran the incidence prevalence step, but two of them were not included in this article as they did not obtain data sharing approval on time. Additionally, from the 52 included databases, 42 ran the large-scale patient-level drug utilisation step (**Figure 1**)

**Figure 1.**
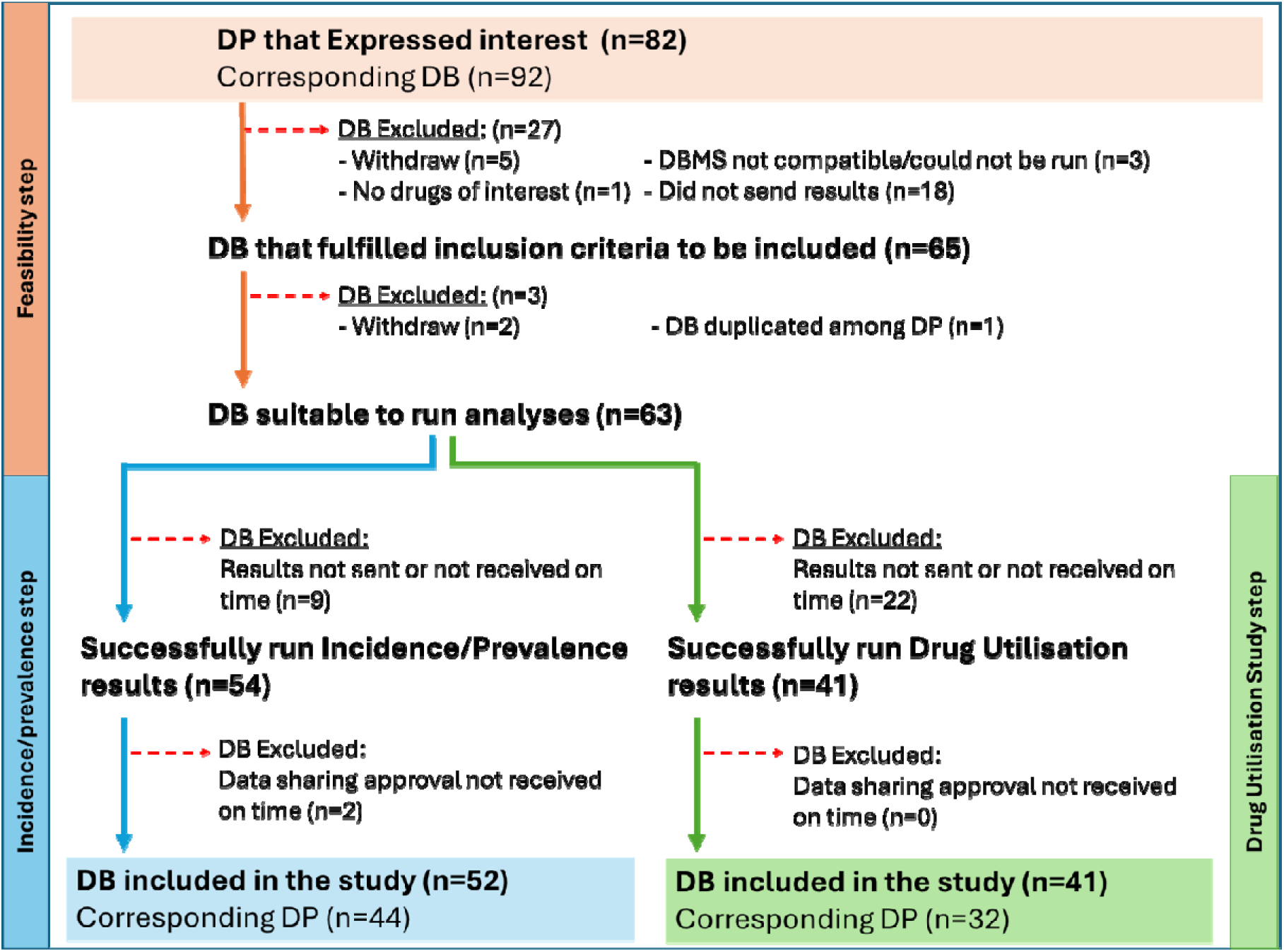
Attrition of the data bases included in the study. Abbreviations: DB, databases; DBMS, database management system; DP data partners.

In the end, 44 EHDEN data partners participated in the study contributing results from 52 data sources from 18 European countries (including Belgium, Bulgaria, Estonia, Finland, France, Germany, Greece, Hungary, Israel, Italy, Netherlands, Portugal, Romania, Serbia, Spain, Sweden, Türkiye, UK) and 3 data sources from the United States (not part of the EHDEN network but provided by the EFPIA partners). (**Figure 1**)

The contributing data sources originated from different health care settings and comprised various types of real-world data including electronic medical records (primary and secondary care), administrative claims, and disease registries. A brief description of all included databases is provided in the **Annex I,** and all database acronyms are available in **Table 1**.

**Table 1.**
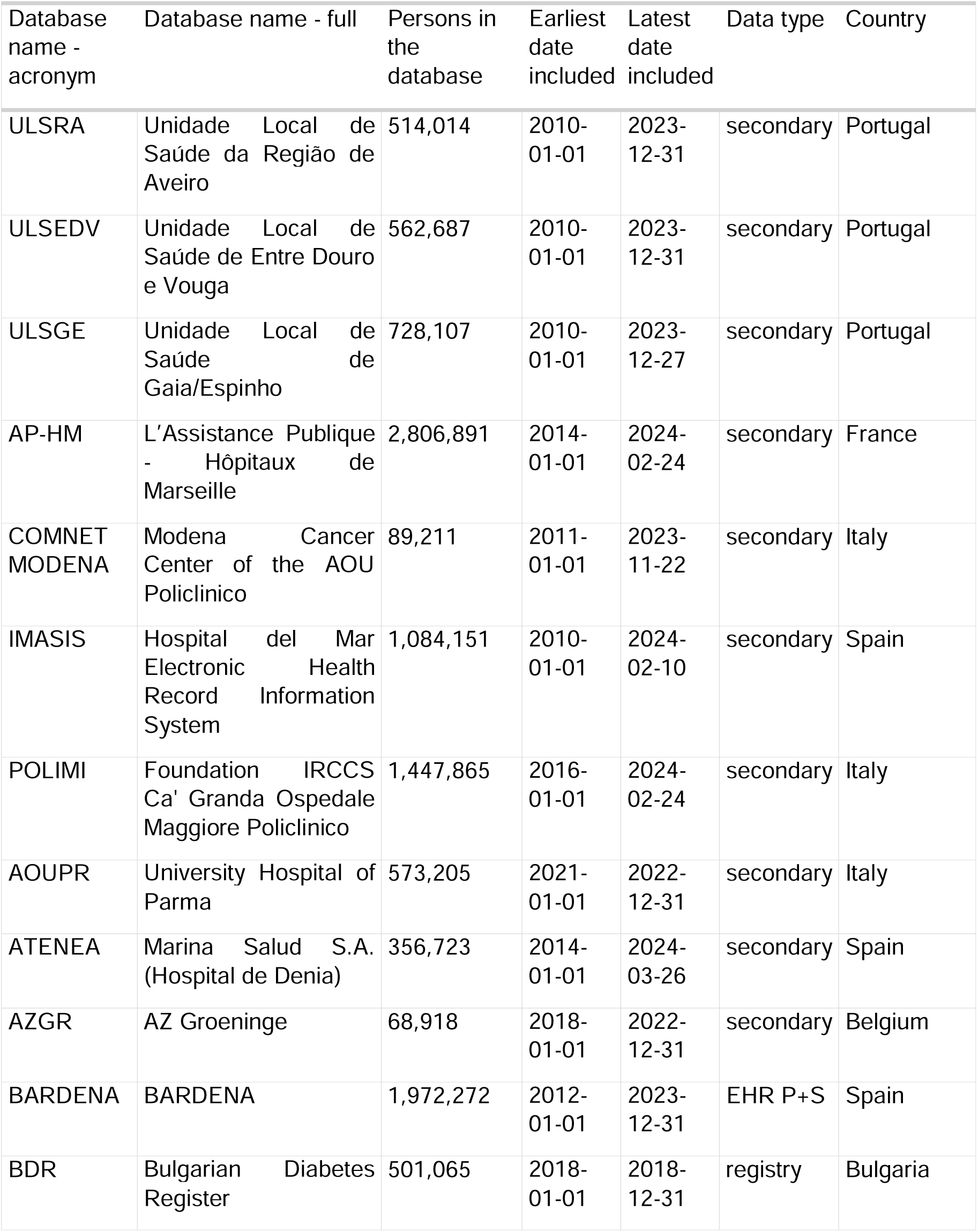
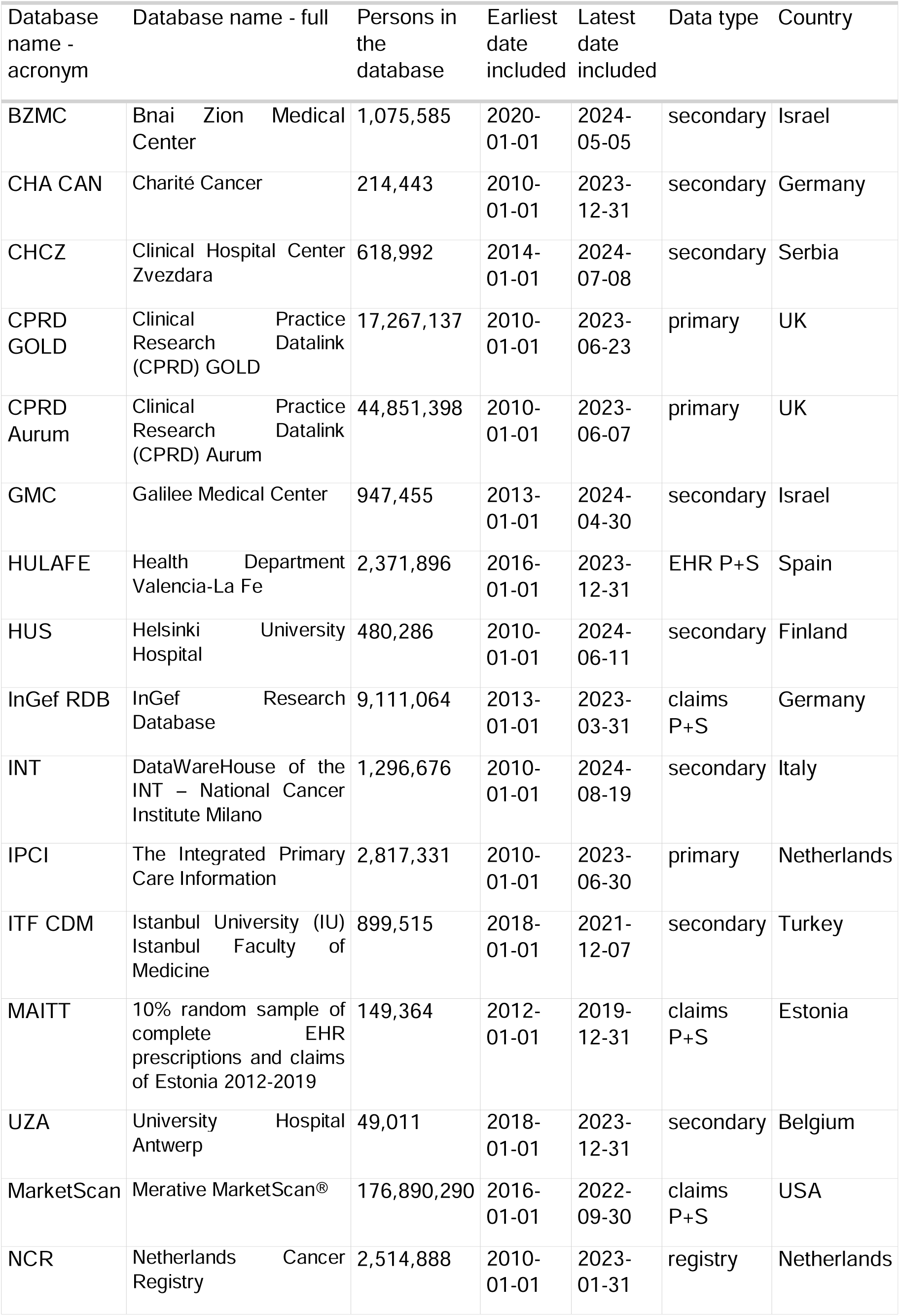
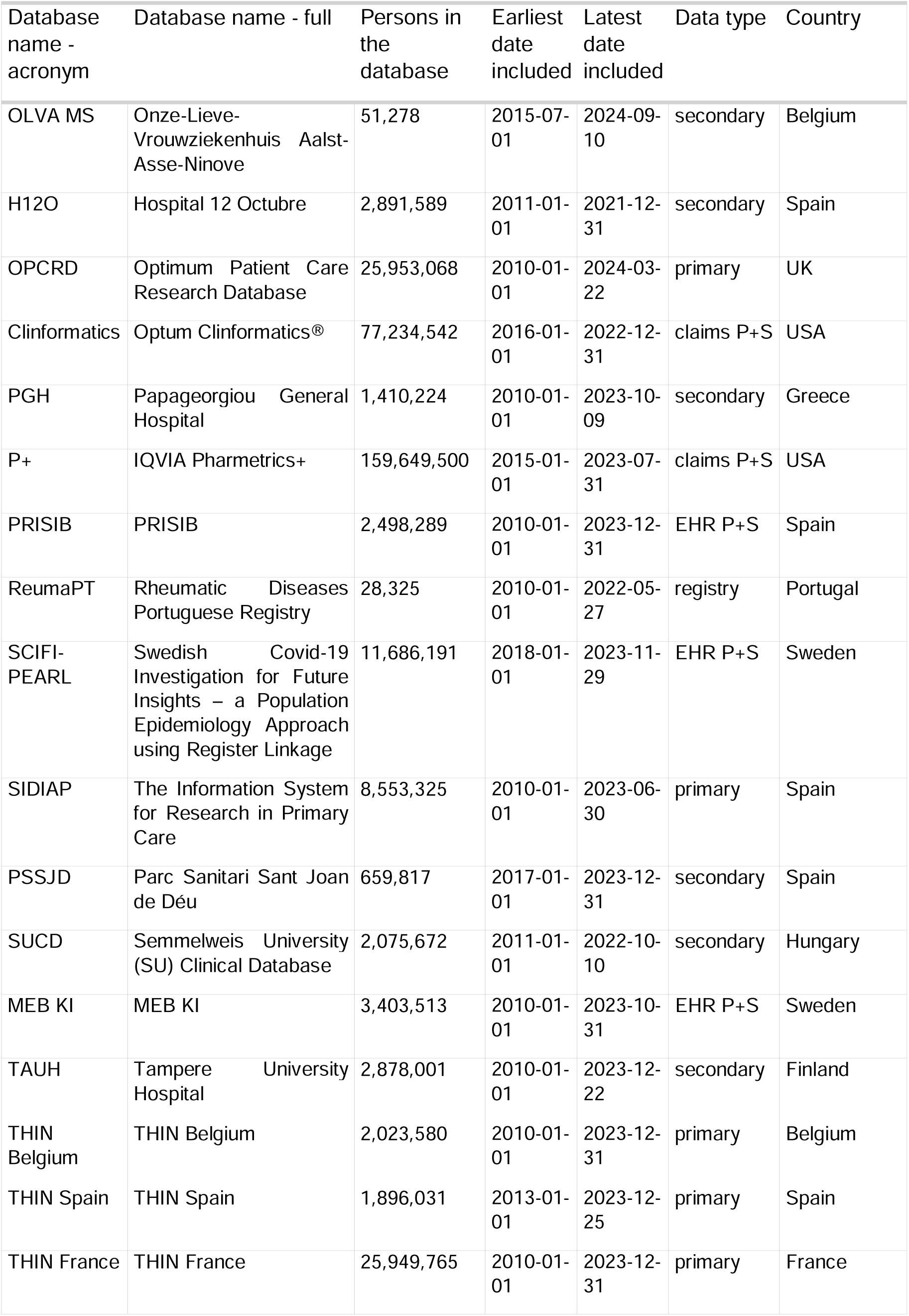
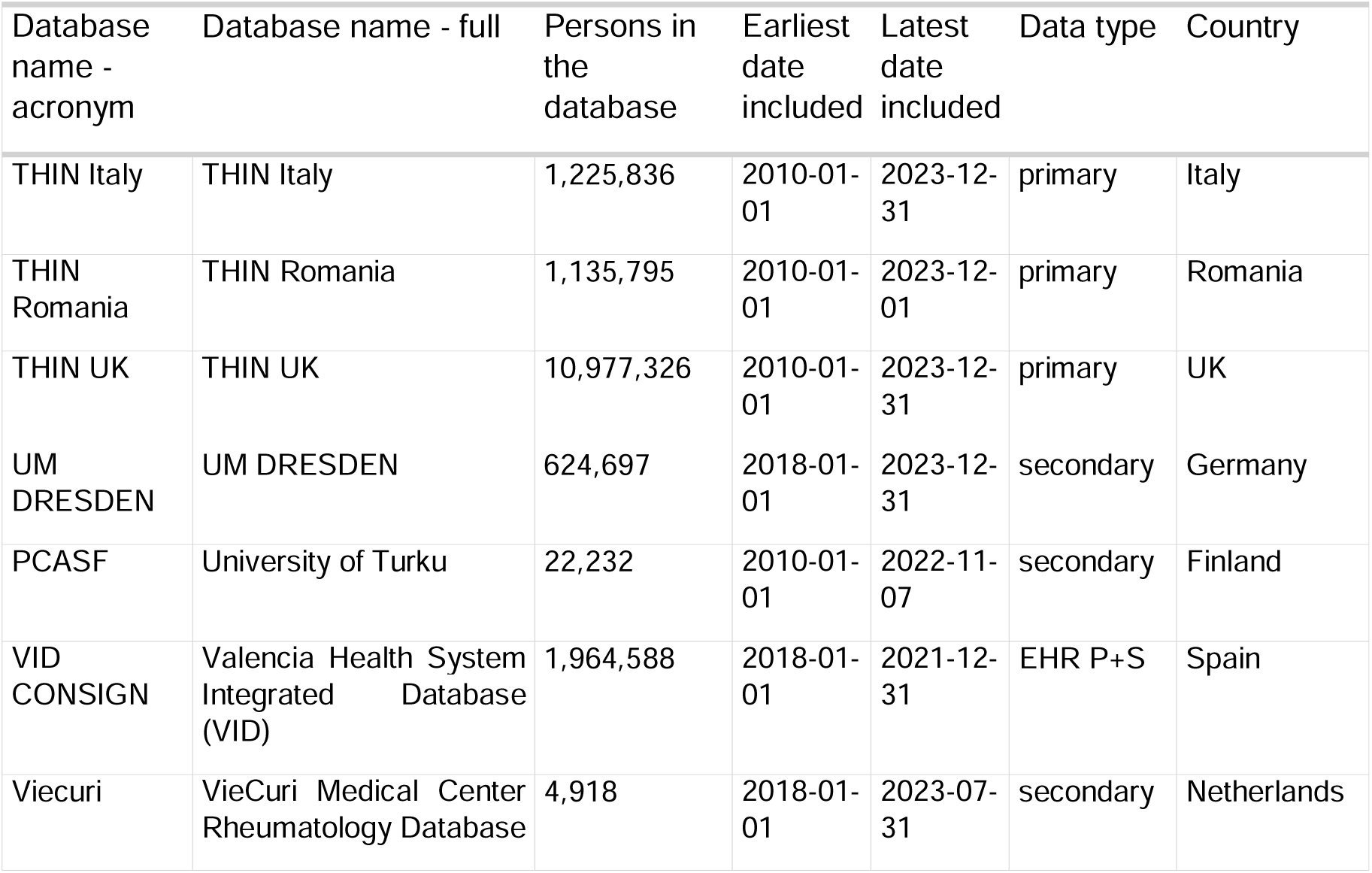
Description of databases included in the study. It contains the list of databases names and acronyms, data type, country, number of persons and period of the study covered in each of them.

### Study setup

The study was set up as a federated network study, where each data partner accessed and analysed their own respective databases locally. Before study initiation, test runs were conducted on all data sources as a feasibility step: to participate partners were required to have at least one drug of interest recorded in more than 100 individuals within the study period. Only after passing the feasibility step successfully were data partners permitted in the second step to execute the analytical package for the Incidence-Prevalence and in the third step the analytical package for the patient-level Drug Utilisation Study (DUS) against their participating data sources. (**Figure 1**)

### Study period

The study period covered the years from 2010 (or the following earliest available data) to the latest data collection date for each data source (**Table 1)**.

### Study population

1. For the population-level drug utilisation study, the study cohort comprised all individuals present in each database during the study period with at least 365 days of data availability before the day they become eligible for study inclusion. This requirement of at least 365 days of data history was not applied for children < 1 year old. Additional eligibility criteria were applied for the calculation of incidence rates, where the observation time of users of the medicine of interest was excluded during use and 30 days afterwards as a wash out period.
2. For the patient level drug utilisation study, the study populations comprised all incident and prevalent users of medicines with suggested shortages and their alternatives in the period between 2010 and the end of the study period. Patients could be eligible more than once during the study period.

### Variables

The list of drugs under study was sourced from the EMA shortages catalogue during the study period.^7^ We included all medicines listed as having had an ongoing shortage for at least 365 days (cut-off end date was September 25^th^, 2023): agalsidase beta, alteplase, amoxicillin, amoxicillin-clavulanate, arsenic trioxide, belatacept, c1 esterase inhibitor, ceftolozane-tazobactam, ganirelix, imiglucerase, sarilumab, tenecteplase, tigecycline, tocilizumab, varenicline and verteporfin.

The list of key alternative drugs which could be prescribed to replace drugs in shortage was drawn either directly from suggestions made by the EMA or by using the ATC group the medicine was in or based on recommendations either from treatment guidelines or from domain experts and included a total of 39 drugs. Cetrorelix, which had been included as alternative for ganirelix, was listed as in shortage in the EMA shortages catalogue. However, its shortage was shorter than 365 days.

The final list of drugs included 13 antibiotics, 7 cancer treatments, 3 eye health treatments, 2 smoking cessation treatments, 22 rare disease medications, 3 thrombolytic agents, 2 assisted reproduction treatments and 5 transplant medications. An exhaustive list of all the drugs included is available in the **Supplementary Table 1.**

### Statistical analysis

All databases had to pass an initial feasibility check before being included in the study to ensure the minimum requirements to participate (described in Study setup section). We assessed patient baseline characteristics in a large-scale characterization study focusing on demographic characteristics (age, sex) and conditions recorded prior to exposure.

Period prevalence of use, with 95% binomial confidence intervals, was calculated by calendar year, defined as the total number of individuals receiving the drug of interest during the given calendar year, divided by the population at risk of being exposed during the same period. Annual incidence rates with 95% Poisson confidence intervals of use for the medicines of interest were calculated as the number of new users after 30 days of no use divided by the time at risk of getting exposed during that period. Incidence was evaluated in terms of annual incidence rates, that is, the number of new users (30 days of no drug use) divided by the time at risk of getting exposed during that period. We also performed stratified analyses based on the following covariates:

- **Period (a.k.a. calendar time)**: We considered stratified analyses based on the calendar year for medicines being listed as (suggested) shortages and their alternatives. When stratifications by calendar year did not allow the observation of changes once shortage was declared, monthly or quarterly periods were considered.
- **Healthcare setting**: We considered stratified analyses based on healthcare settings and data types, including, primary care, secondary care, both combined (primary and secondary care), claims and registries.
- **Demographic characteristics:** We considered stratified analyses based on age bands, that is, younger than 18 versus between 18 and 64 versus older than 65 years old, and sex, that is male versus female patients.

Incidence and prevalence analyses were executed across all participating databases using the *IncidencePrevalence* R-package, part of the DARWIN EU® open-source software library.

In post-hoc analyses, we assessed the difference in average incidence and prevalence between the 3 years after and the 3 years before the announced shortage. A use of < 67% after the announced shortage (i.e. a drop of >33% in use) was considered as indication that the medicine was in shortage in the particular database (“suggested shortage”), health care setting or country. ^11, 12^

The patient-level drug utilisation study performed a characterization of prevalent and new users including the following characteristics:

- Age (median and proportion by age groups: <18, 18-64, 65+) and Sex (proportion male and female patients)
- Indication: We assessed the diagnosis within pre-specified time windows (at index date, and 30 days or any time prior to index date). The number and the percentage of persons with a record of a potential indication within each time window are provided. If a person had records for more than one potential indication, they were included in both indication groups.
- Treatment duration: This was calculated as the duration of drug exposure for the new dispensation/prescription of a medicine of interest. Treatment duration is summarized providing the minimum, p25, median, p75, and maximum duration. For databases where duration could not be calculated due to missing information, treatment duration is not provided.
- Dosage: Daily dosage of treatment was per medicine of interest and administration route. Dose is summarized providing the minimum, p25, median, p75, and maximum dose. For databases, where dose could not be calculated due to missing information, dose is not provided.

The drug utilisation study was executed using the *DrugUtilizationCharacteristics* R-package, part of DARWIN EU® open-source software

### Results interpretation

A full-day webinar was conducted, bringing together medical experts, epidemiologists, and data scientists from all EHDEN data partners who contributed to the study. The incidence and prevalence results were visualized in an R shiny application and presented in a plenary session using one drug group as an example, followed by breakout sessions for each of the 11 drug groups to collaboratively analyse and interpret the results, and to document and summarize the tentative conclusions and limitations. The results were analysed per database and stratified by healthcare setting, age group, sex and country, considering available information from the literature and local health care practice on time windows and potential reasons for drug shortages during the study period.

A second full-day webinar was held three months later in a similar setting to analyse and interpret the results of the second part of the research with the patient-level drug utilisation study.

## Results

We included the results of 52 data sources from 18 European countries and the US. Size of data sources ranged from 4,918 individuals to more than 176 million, but the total number of persons included in the denominators for incidence and prevalence was 617.06 million. (**Table 1**)

### Drugs with suggested shortage

We observed the incident use of 15 drugs with suggested shortages, and the prevalent use of 16 drugs with suggested shortages. (**Figure 2**, **Figure 3**)

**Figure 2.**
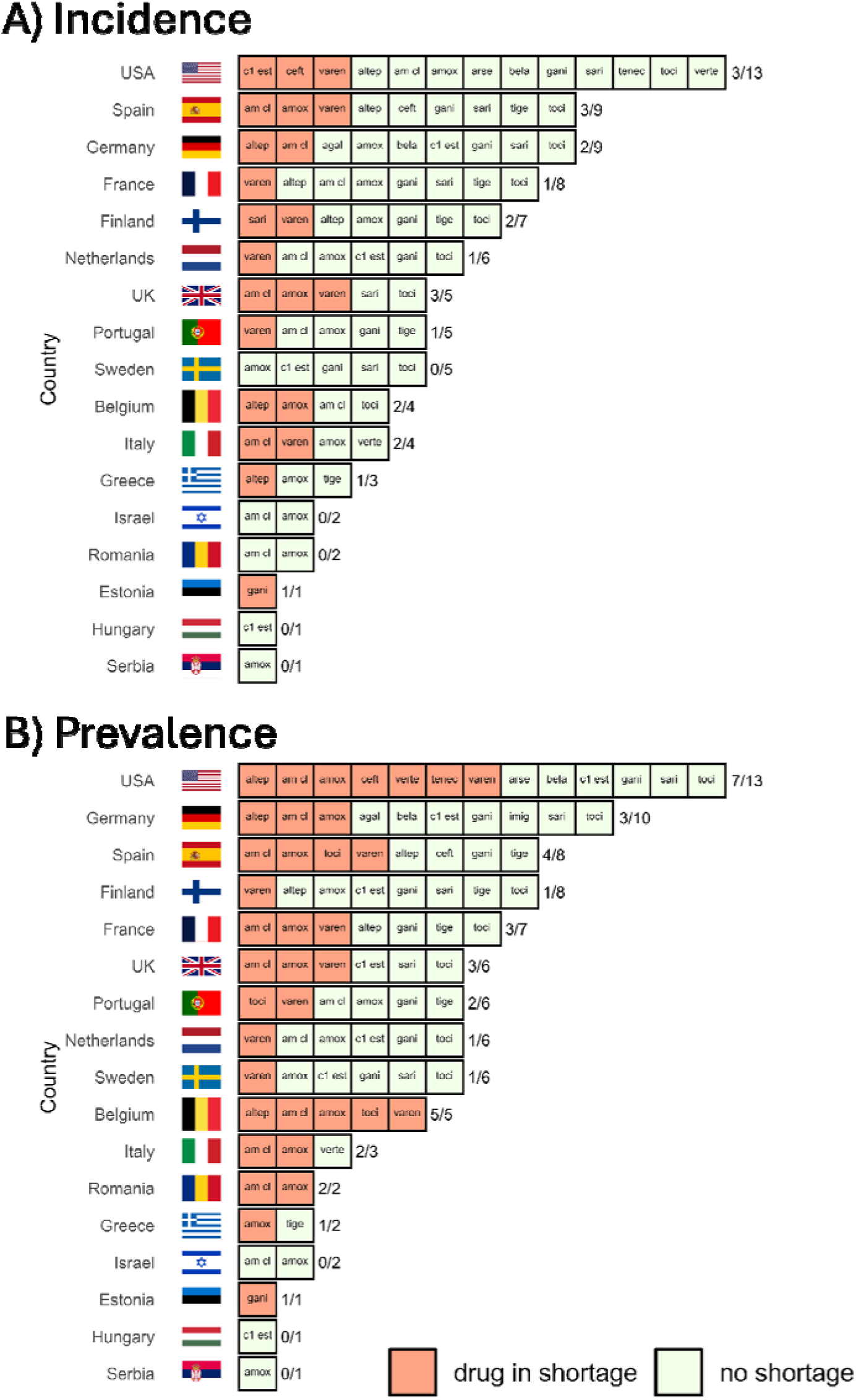
Shortages of drugs by country in A) incident use or B) prevalent use. To be considered as drug in shortage, at least one database of the study in the country had to present a ≥33% drop in use (in incidence or in prevalence). When any of the databases in the country show a ≥33% drop in use after the shortage announcement, we considered it as no shortage observed in the data. Abbreviations: agal, agalsidase beta; altep, alteplase; amox, amoxicillin; am cl, amoxicillin-clavulanate; arse, arsenic trioxide; bela, belatacept; c1 est, c1 esterase inhibitor; ceft, ceftolozane-tazobactam;; gani, ganirelix; imig, imiglucerase; sari, sarilumab; tenec, Tenecteplase; tige, tigecycline; varen, varenicline; verte, verteporfin.

**Figure 3.**
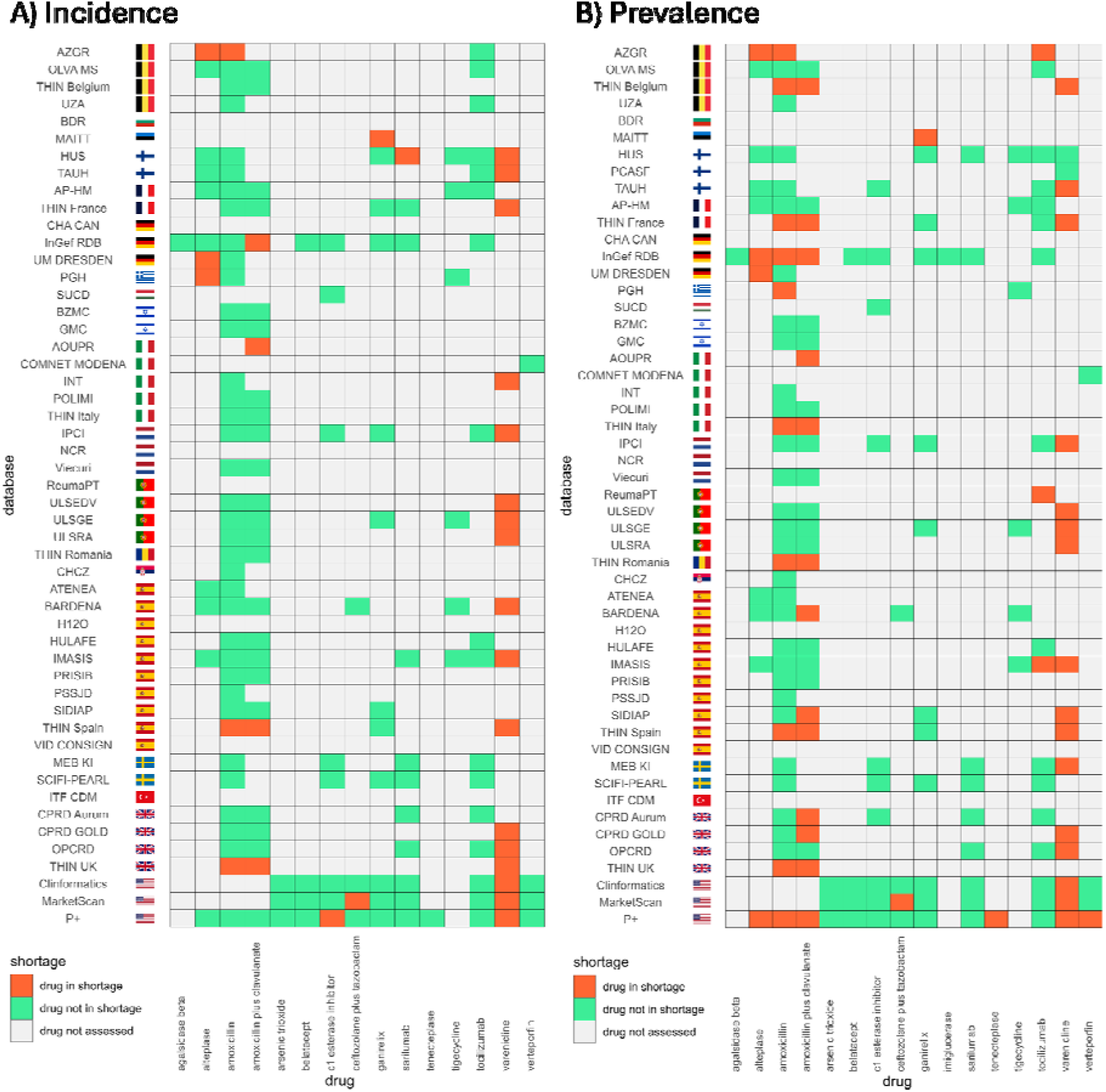
Shortages of drugs by all study database in A) incident use or B) prevalent use. Drug in shortage: the drug presented a ≥33% drop in use (in incidence or in prevalence) after the shortage announcement. Drug not in shortage: the drug did not have a ≥33% drop in use after the shortage announcement. Drug not assessed: drug not observed in the database and therefore not assessed.

The US presented the largest number of drugs in shortages, with 7 drugs out of 13 with a drop in prevalent use larger than ≥33% (post-announcement versus pre-announcement), followed by Belgium with all 5 drugs observed, and Spain with 4 out of 8. The countries with more drugs presenting ≥33% drop in the incident use were UK (3 out of 5), Spain (3 out of 9) and US (3 out of 13). (**Figure 2, Supplementary Table 2**)

Focusing on the drugs with suggested shortages, varenicline was the drug with the largest number of databases (**Figure 3**) and countries (**Supplementary figure 1**) with a ≥33% drop in incidence of use and/or prevalence of use: 17 databases in incidence, 17 in prevalence (with some different databases in respect to the incidence) from up to 10 countries. This was followed by amoxicillin alone and combined with clavulanate, where 3 and 4 databases for incidence of use (respectively), 10 and 13 databases for prevalence of use (respectively for alone and combined), presented a ≥33% drop; affecting up to 9 for amoxicillin alone and 8 when combined with clavulanate.

### Drugs that were more frequently in shortage and its alternatives

The following sections are focused on the drugs that were more frequently in shortage as detected in the databases of the study. The complete results for incidence and prevalence, including all the drugs and their alternatives, are available in a shiny app: https://dpa-pde-oxford.shinyapps.io/MegaStudy_IncidencePrevalence/

#### Smoking cessation treatments

Shortage of varenicline was announced on the 8th of July 2021 by the EMA. We observed a shortage in incidence of use in 12 databases (**Figure 4**), whilst in prevalence of use was 13 databases (**Figure 5**). The shortage was more prevalent in primary care (56% [5 out of 9] databases assessed presented a shortage), followed by the other types of data containing primary and secondary combined, claims and registries (present in 50% [5 out of 10]), and hospitals (present in 20% [3 out of 15]).

**Figure 4.**
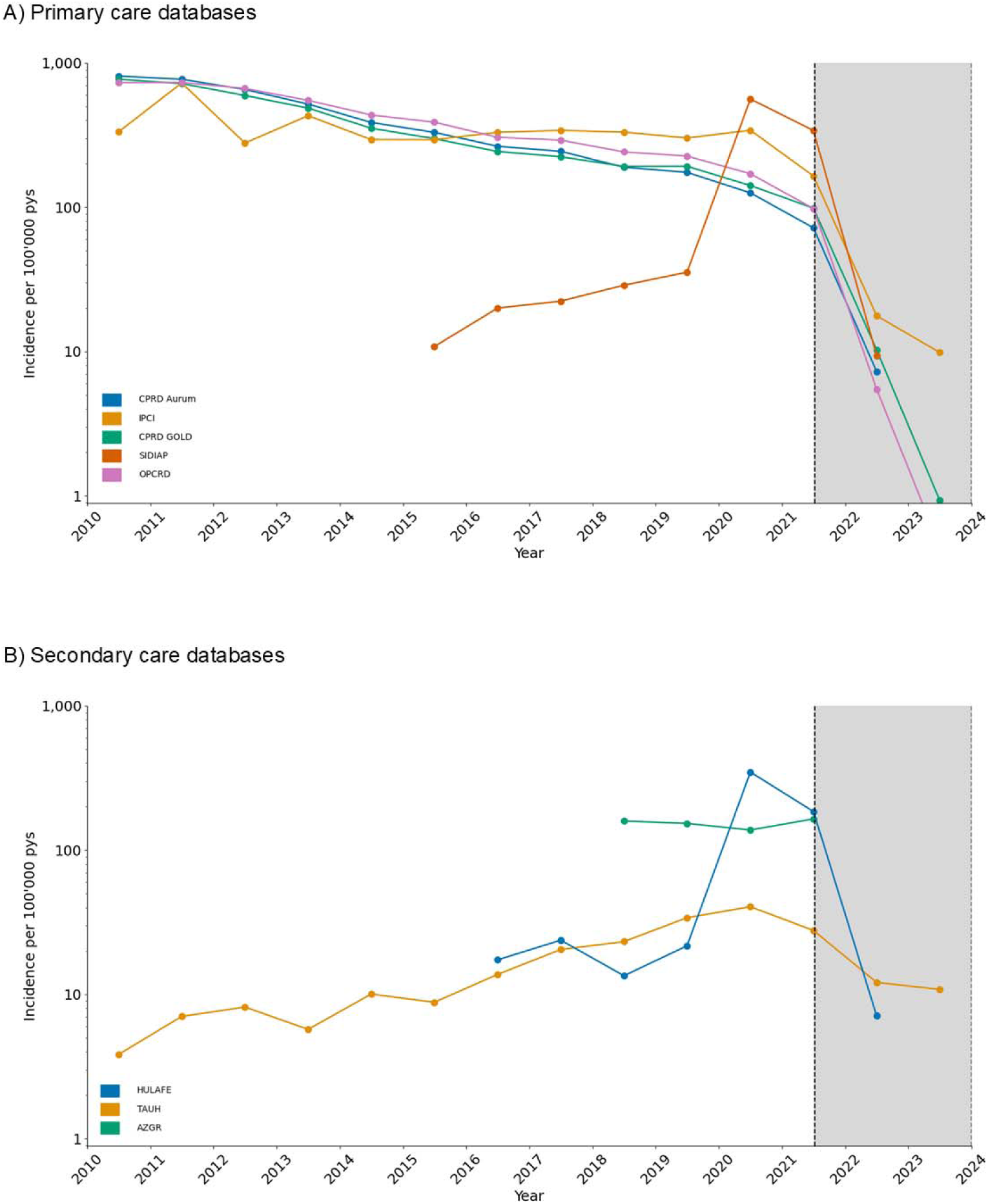
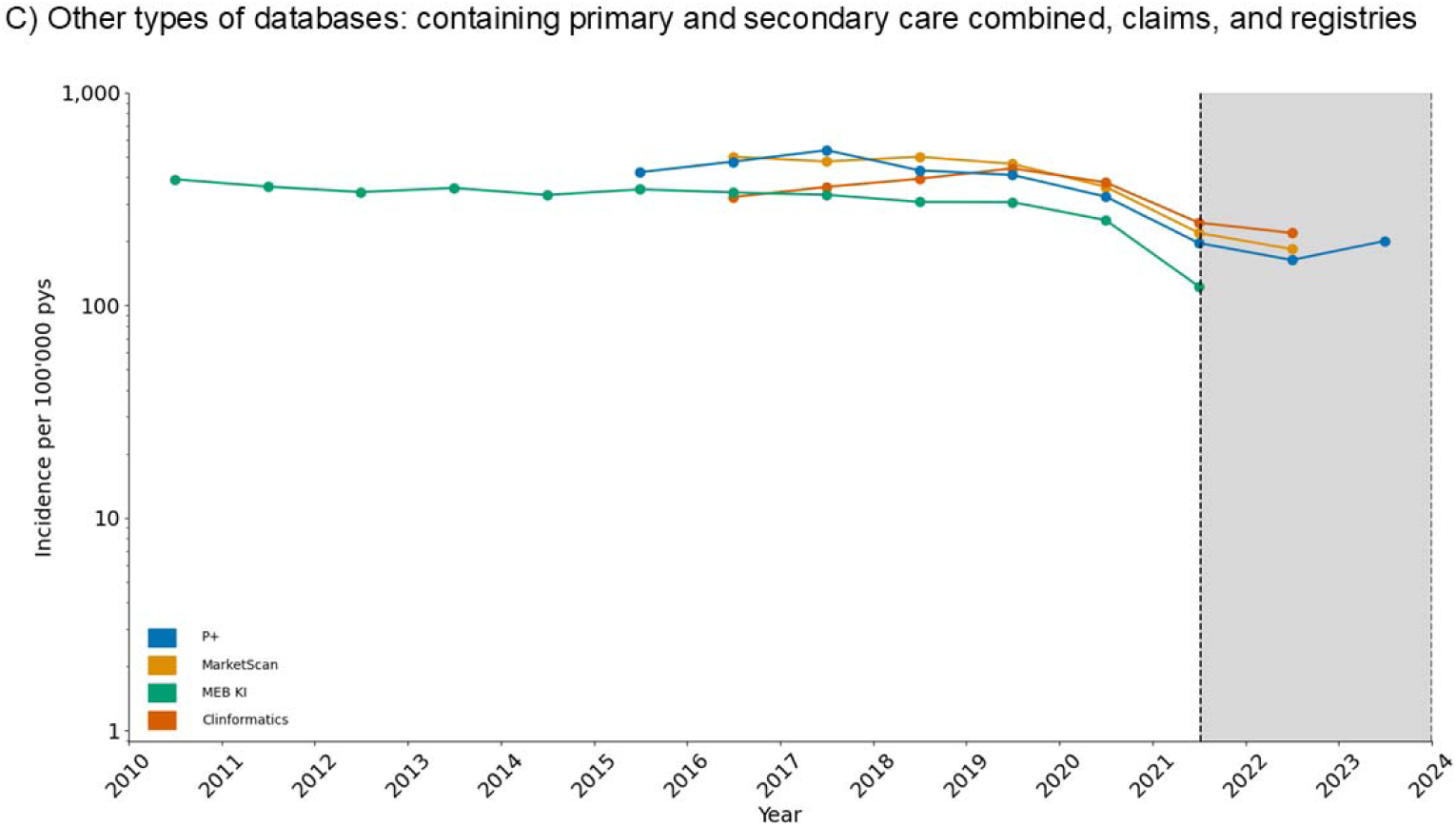
Incidence of use for varenicline per 100,000 person-years in the databases presenting. **a** ≥30% drop in incident use after shortage was announced (area in grey): A) Primary care databases, B) Secondary care databases and C) Other types of databases containing primary and secondary combined, claims and registries. Claims databases contain records from primary and secondary care data. Secondary care databases shown are all hospitals.

**Figure 5.**
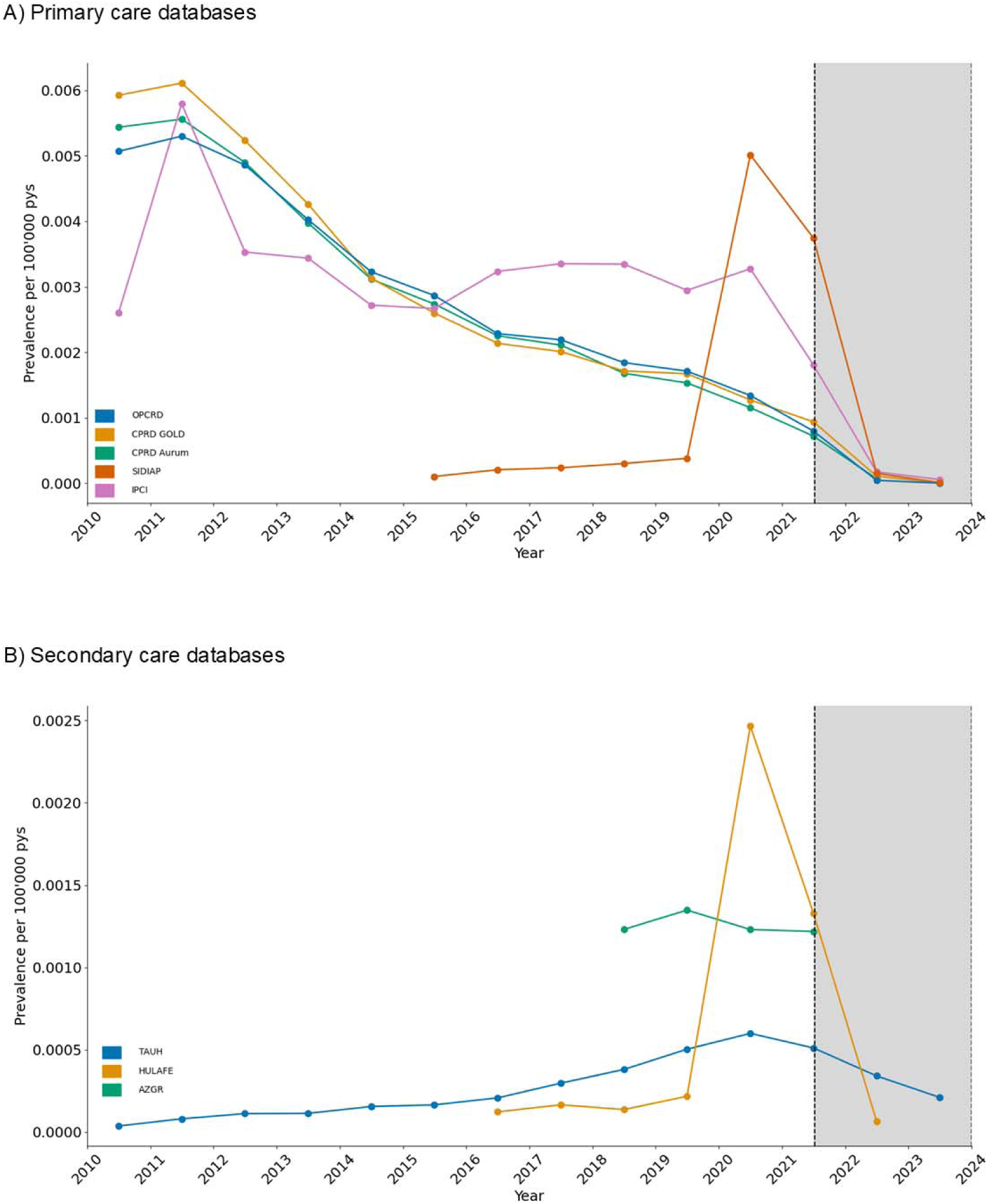
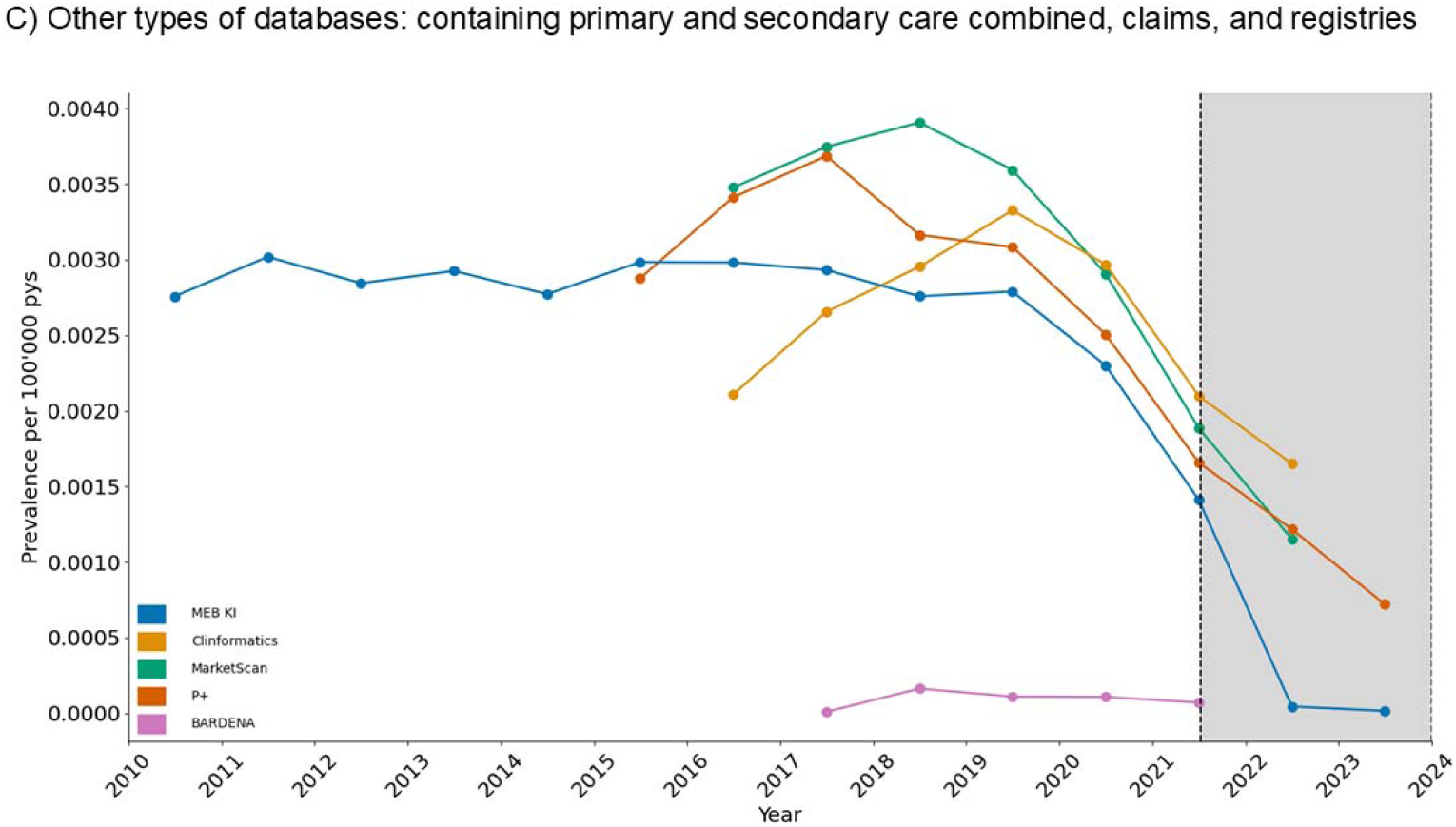
Prevalence of varenicline in the databases presenting. **a** ≥30% drop in prevalent use after shortage was announced (area in grey): A) Primary care databases, B) Secondary care databases and C) Other types of databases. Claims databases contain records from primary and secondary care data. Secondary care databases shown are all hospitals.

Despite not reaching the ≥33% threshold, most primary care databases show a decrease in the incidence of varenicline, whilst incident use of its alternative, nicotine supplements, increased after July 2021 in IPCI, OPCRD, THIN FRANCE, presented a peak in 2022 in THIN BELGIUM and THIN UK, or show a stabilisation in the incident use after a decrease trend in CPRD AURUM and GOLD. (**Supplementary Figure 2A**) When assessing prevalence of varenicline and nicotine in primary care, similar patterns of use were observed in all databases. (**Supplementary Figure 2B**)

#### Common infection antibiotics

Shortage of amoxicillin alone and in combination with clavulanate was announced by the EMA on the 27th of January 2023. We observed a shortage in prevalence of use of amoxicillin in 22 databases out of 46, whilst in amoxicillin with clavulanate was 16 databases out of 34. (**Figure 6**) The shortage of both, amoxicillin alone and in combination with clavulanate, was more prevalent in primary care (55% [6 out of 11] presented a shortage). We observed shortages of amoxicillin alone and in combination with clavulanate in 46% (11 out of 24) and 43% (6 out of 14) of hospitals, respectively. In the other databases, 45% (5 out of 11) presented a shortage in amoxicillin, and 44% (4 out of 9) when combined with clavulanate.

**Figure 6.**
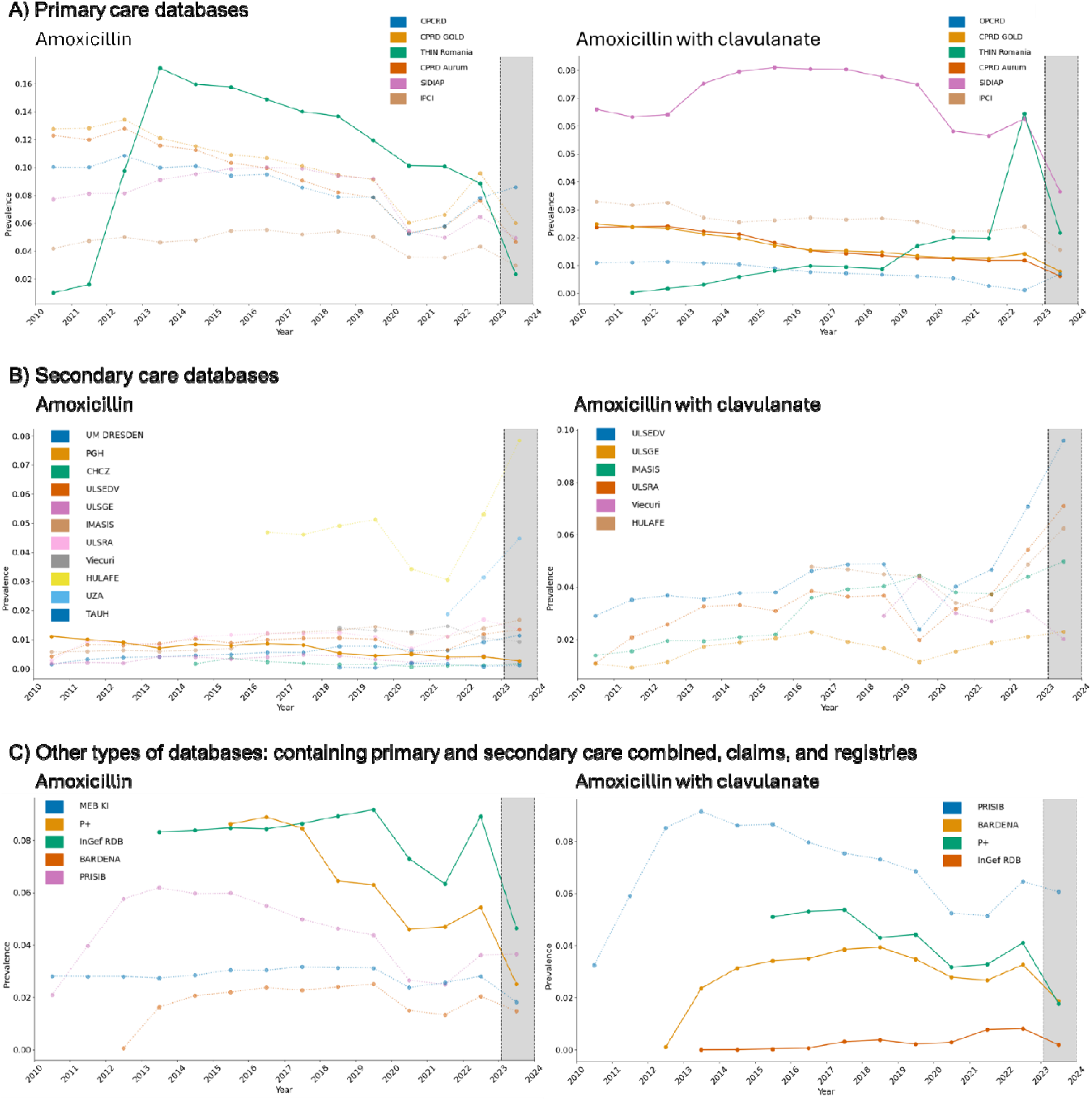
Prevalence of amoxicillin alone or combined with clavulanate in the databases presenting. **a** ≥30% drop in prevalent use after shortage was announced (area in grey): A) Primary care databases, B) Secondary care databases and C) Other types of databases. Claims databases contain records from primary and secondary care data. Secondary care databases shown are all hospitals.

Prevalence of amoxicillin alone and in combination with clavulanate generally decreased in the observed primary care databases, and most of them showed a marked drop in 2020 during the COVID-19 pandemic. Nine of 11 primary care data bases showed an upsurge in 2022 that partially recovered levels before the pandemic onset, however, prevalence of use in 2023 decreased again. (**Supplementary Figure 3A**).

Ten hospitals out of 24 did not show any prevalent use of amoxicillin-clavulanate but had a prevalent use of amoxicillin alone. (**Supplementary Figure 3B**) Prevalent use of amoxicillin-clavulanate increased between 2020-2022, after the COVID-19 pandemic onset, in 8 out of 14 hospitals, whilst for amoxicillin alone this occurred in 12 out of 24 hospitals.

Prevalence of use for amoxicillin alone or combined with clavulanate in 9 of the 11 other databases also showed a decrease, with the exception of HULAFE and for amoxicillin-clavulanate in MAITT. (**Supplementary Figure 3C**).

When the alternative antibiotics used for common infections were observed, prevalence of azithromycin increased in 6 out of 11 primary care databases, but also presented a drop in 2020-2021, whilst clarithromycin decreased in all 11 databases. Similar decreasing trends could be observed for phenoxymethylpenicillin. Additionally, prevalence of use for benzylpenicillin was very low in primary care, below 0.16%. In secondary care, prevalence of use for azithromycin increased in 18 out of 24 databases, clarithromycin presented a generally stable trend in 50% of hospitals (in 12 out of 22), use of phenoxymethylpenicillin was uncommon (only observed in 9 databases of 24), and benzylpenicillin presented a prevalence of use of <0.2% except in HUS that increased from 2010 to 2023 (maximum observed prevalence of 1.3%) and in BZMC that increased from 2020 to 2023 (maximum observed prevalence of 0.7%). In the other types of databases, azithromycin decreased in 8 of 11, with a small upsurge in 2022 in 6 databases; whilst clarithromycin and phenoxymethylpenicillin prevalence generally decreased over time. **Supplementary figure 4** contains the prevalence of alternative antibiotics for common infections.

### Characterisation of users of drugs in shortage

We received results of the large-scale patient-level drug utilisation study of 42 databases. In this section, we focus on discussing the drugs on shortage that presented larger differences in the characterisation. Full results are available in a shiny app: https://dpa-pde-oxford.shinyapps.io/MegaStudy_webinar_DUS/

#### Smoking cessation treatments

Demographics of incident and prevalent users of varenicline show that the median age was similar before and after the shortage period. However, a change in the proportion of sex was observed in UK primary care databases (CPRD Aurum and OPCRD), a Spanish hospital (HULAFE) and in one of the US claims data (P+) for incident (**Supplementary Figure 5A**) and prevalent users (**Supplementary Figure 5B**).

A primary care database from the Netherlands (IPCI) shows a drop in cumulative dose after the shortage date in both, incident and prevalent users (**Supplementary Figure 5C and D**, respectively). Moreover, TAUH (Finland, secondary care) and THIN Belgium (Belgium, claims data) show a reduction in duration after the shortage date (**Supplementary Figure 5E**), whilst SCIFI-PEARL (Sweden, EHR P+S) saw a reduction in initial quantity after the shortage date (**Supplementary Figure 5F**).

#### Thrombolytic agents

After the announcement of alteplase shortage, median age of incident prescription of alteplase were observed increased in AP-HM, CPRD Aurum, InGef RDB and OLVA MS, whilst slightly decreased in ATENEA, IMASIS and UM DRESDEN (**Supplementary Figure 6A**). However, when stratified by age groups, HUS, which give the impression that was stable, show an increased prevalence of the 19-64 age group and a decrease in the 65+ age group (**Supplementary Figure 6B**).

In the year of the shortage (2022), an increase in incident male users can be seen in 6 of 10 observed databases, but 7 databases presented a decrease of male users in 2023 compared with the proportion in the previous year (**Supplementary Figure 6C**). Trends of prevalent patients are similar (**Supplementary Figure 6D**). We observed a reversal in the trend of alteplase usage between sexes in the IMASIS, HUS and UM DRESDEN databases after alteplase shortage announcement. (**Supplementary Figure 6C and D**).

In the AP-HM and UM DRESDEN databases (which contain secondary data), an increase in the incidence of alteplase administered to patients with pulmonary embolism (PE) in the last 30 days was observed during the drug shortage period. Conversely, in the HUS database, this pattern showed a decrease (**Supplementary Figure 6E**). Comparing with PE diagnoses from any time prior, a similar trend to that observed in recently diagnosed patients can be seen in the HUS and UM DRESDEN databases. However, in the AP-HM database, a decrease in the incidence of alteplase administered to patients with PE diagnosed at any prior time was observed. (**Supplementary Figure 6F**).

In France, Finland, and Germany, an increase in the prevalence of alteplase use is observed despite its shortage (**Supplementary Figure 6G**). In the AP-HM and UM DRESDEN databases (which contain secondary data), an increase in the incidence of alteplase administered to patients with PE in the last 30 days was observed during the drug shortage period. Conversely, in the HUS, IMASIS and OLVA MS databases, this pattern showed a decrease (**Supplementary Figure 6H**). Comparing with PE diagnoses from any time prior, a similar trend to that observed in recently diagnosed patients can be seen in the HUS, UM DRESDEN, IMASIS and OLVA MS databases. However, in the AP-HM database, a decrease in the incidence of alteplase administered to patients with PE diagnosed at any prior time was observed (**Supplementary Figure 6I**).

Conversely, we did not observe a significant change in myocardial infarction (MI) and ischemic stroke indications for alteplase (*data shown in the shiny app*).

#### Disease-modifying antirheumatic drugs (DMARDs)

Characteristics of sarilumab users are reported in **Supplementary Figure 7**. After the announcement of sarilumab shortage, median age of incident prescription was observed to have increased in HULAFE and InGef RDB, whilst SCIFI-PEARL show an increase in age in the same year of the shortage that is later maintained (**Supplementary Figure 7A**). In the year of the shortage, an increase in incident male users can be seen in most of the databases, but it is most prominent in HULAFE and CPRD-Aurum. Trends of prevalent patients are similar (**Supplementary Figure 7C**).As for the indications, a significant decrease of rheumatoid arthritis (the most frequent diagnosis) can be observed in incident patients in the year of the shortage in CRPD-Aurum, HUS and SCIFI-Pearl databases. (**Supplementary Figure 7D**). Prevalent users presented similar trends. Additionally, a decrease in cumulative dose of incident users can be observed in IPCI (**Supplementary Figure 7G**).

## Discussion

Our study contains information on the use of 57 drugs across 52 databases that were mapped to the OMOP-CDM.^9^ To our knowledge, this is the largest network cohort study conducted to date, and the most comprehensive analysis of drug shortages internationally. The massive use of Real-World Data opens a way of addressing this serious problem with a more objective and evidence-based approach. ^11–13^

We studied drug shortages across drugs listed in the EMA shortages catalogue for more than a year.^7^ We defined a drop of ≥33% in incidence or prevalence of use after the EMA announced a shortage in their catalogue as a confirmation of the announced shortage across different countries. Prior efforts in detecting shortages using trends of use in the US have suggested that a ≥33% drop is an appropriate definition to detect drug shortages. ^11, 12, 14^

In our analyses, varenicline showed a decline compatible with a shortage in both incidence and prevalence in most of the databases of the study in which its use was observed. Additionally, we observed a drop in cumulative dose after the shortage date in both, incident and prevalent users in the database from the Netherlands (IPCI), a reduction in duration after the shortage date in TAUH (Finland, secondary care) and THIN Belgium (Belgium, claims data) databases, and a reduction in initial quantity after the shortage date in SCIFI-PEARL (Sweden, EHR P+S) were most probably the reflection of drop in incidence/prevalence of varenicline usage and can only be considered as surrogate findings. Varenicline shortage in June 2021 was caused by a disruption in its production: batches of Champix (varenicline commercialised by Pfizer) were found to contain levels of N-nitroso-varenicline exceeding acceptable EU limits.^15^ This led to the recall of several batches, and a subsequent pause in production 3 months later, which resulted in a shortage of the medication affecting the UK, the US and all EU member states except Bulgaria.^15, 16^ Despite a varenicline shortage, we did not observe an increase in the use of nicotine replacement products, which are the most readily available alternative. This might be because nicotine is not the only pharmacological intervention for smoking cessation. In addition to nicotine replacement therapy, other alternatives include bupropion, cytisine, and nortriptyline.^17, 18^ Additionally, e-cigarettes are becoming more popular ^19^, and some countries have included them in their guidelines ^20^. Nicotine replacement therapies are also easily accessible over the counter. Therefore, the electronic health records and prescription data are probably not the best source to assess the usage of nicotine replacement therapies.

The following drugs that had many databases with an observable decline in prevalent use ≥33% was amoxicillin and amoxicillin-clavulanate. We also observed that the shortages were more prevalent at primary care settings than the others. Amoxicillin and amoxicillin-clavulanate are antibiotics widely used for common infections.^21^ Shortage of amoxicillin and amoxicillin-clavulanate were caused by issues in manufacturing plus an increase in demand. The combination amoxicillin-clavulanate was used especially at the start of the COVID-19 pandemic to treat secondary bacterial infections such as pneumonia, at a time when no evidence of risk-benefit of COVID-19 treatments were yet available.^22^ Subsequent decreases in the use of amoxicillin (alone or with clavulanate), as well as of alternative antibiotics, during the COVID-19 pandemic could be attributed to the impact of public health restrictions on non-COVID infections and to a reduced interaction with the health services.^23^ An upsurge in respiratory infections during winter 2022 that especially affected the UK^14^ also contributed to the increase in demand that led to the announcement of the shortage in January 2023.^24^ The reasons announced by EMA for this shortage were both increased demand of antibiotics and manufacturing delays and production capacity issues, including big pharmaceutical companies leaving the antibiotic market to companies producing generic drugs and failing in the development of new antibiotics due to commercial reasons.^25^ Awareness about the risk of serious life-threatening conditions related to shortages of antibiotics should be raised and new more effective measures for drug production and distribution should be taken to address this problem.^26^

In addition of the ≥33% drop in use, shortages can also be studied by observing the changes in the patients’ characteristics:

In our study, median age of alteplase incident use increased in four databases, whilst slightly decreased in three. Additionally, when stratified by age groups, HUS showed an increased prevalence of the 19-64 age group and a decrease in the 65+ age group. These results may indicate that the presence of atherosclerotic cardiovascular diseases due to age can differ between countries, and in some may occur at earlier ages. Despite increasing age is a significant risk factor for ischemic stroke and MI,^27, 28^ we did not observe an incremented use of thrombolytic drugs for the treatment of these two indications. However, we observed an increased incidence of alteplase administered to patients with PE (especially during and after COVID-19 pandemic) in AP-HM and UM DRESDEN databases, which can be explained by the increased risk of PE due to COVID-19 disease.^29^ Acute ischemic stroke, PE, acute MI and occluded catheters are the main indications of thrombolytics. EMA highlighted the increase in demand as the reason of shortages of alteplase and tenecteplase (announced in September 2022).^30, 31^ Therefore, globally increased prevalence of atherosclerotic cardiovascular diseases.^32^ can be counted to be the main contributor for the shortage. For several reasons, treatment with percutaneous coronary intervention is preferred over thrombolytics in the treatment of acute MI and is increasingly more available, affecting the general trend in thrombolytic agents use, which could help interpreting the lack in increased use of alteplase among MI patients.

For sarilumab, we did not detect a ≥33% drop in use except in the incidence estimates of HUS database. However, we observed that the median age of incident sarilumab prescription increased in HULAFE and InGef RDB. Similarly, SCIFI-PEARL show an increase in age in the same year of the shortage that was later maintained. Moreover, a significant decrease of rheumatoid arthritis indication (only approved indication of the drug) can be observed in incident patients in the year of the shortage in CRPD-Aurum, HUS and SCIFI-Pearl databases. The shortage of sarilumab, is a great example of an unexpected reason for a drug shortage. Sarilumab is an IL-6 receptor blocker that is used as a second or third-line treatment option in the treatment of active rheumatoid arthritis (RA) in adult patients who have responded inadequately, or who are intolerant to one or more disease modifying anti rheumatic drugs (DMARDs). Increased global demand for IL-6 receptor blockers (especially due to off-label usage in the treatment of severe patients during the COVID-19 pandemic), caused an unexpected demand which outweighs the production capacity of the company holding the marketing authorization of sarilumab,^33^ and the other approved IL-6 blocker, tocilizumab.^34^ Our findings were consistent with the announced reason of the shortage. Age by itself is an independent risk factor for COVID-19 mortality and severity^35^ and IL-6 receptor blockers were administered during the treatment of COVID-19 only in patients with severe infection. Keeping this in mind; the results of the present study about increased median age in patients using sarilumab and decreased in the indication of rheumatoid arthritis may point out that off-label usage of sarilumab during the pandemic contributed to this unexpected shortage and patients who needed to use IL-6 blockers for the treatment of rheumatoid arthritis could not access the medication and/or switched to other medications.

As a whole, the causes of drug shortages are very complex and multifactorial, therefore, it may be necessary to resort to different solutions depending on the reason behind the shortage. The prevention of drug shortages which are mostly unpredictable is very challenging. Increasing awareness and informing all the stakeholders by health authorities and regulators, providing risk management plans by manufacturers, incentivizing manufacturers in case of shortage of less profitable treatments, fostering the supply of generic medicines, warning of expected challenges in supply as soon as possible, predicting cost implications, preventing speculative stockpiling, allowing the hospital pharmacists to repackage the drugs to use more efficiently, and the usage of substitute alternative medicines that are expected to have similar effectiveness and safety profile can be considered as basic recommendations and solutions to mitigate the impact of shortages.^36^ Practically, there have been also other ideas that have been discussed and sometimes adopted focusing on various aspects of logistics, e.g., the increased production and use of generic drugs, joint massive orders in scale on behalf of EU instead of each country per se etc. However, these ideas are always subject to various political and administrative caveats and there is no widely accepted solution.

### Strengths and limitations of this study

Our study covers a large number of databases from different health care settings and countries, which provides a global picture of the state of drug use and shortages in Europe. Moreover, as a result of the implementation of the OMOP CDM and standard analytics, all databases run the same analytical code, improving the robustness and reliability of the results.^9^ The code is available in GitHub, which provides full transparency. However, we must acknowledge some limitations of the study:

Values below 10 were censored (with the exception of THIN databases, that was censored for <30) and replaced as NA in the results, and therefore we cannot compute the calculations of ≥33% drop in use on this groups.

The data collection, harmonization and analysis introduce some delay between the decrease of prescription and its impact in the data. This may have influenced the interpretation of the most recent shortages. Additionally, the ETL designs of the databases and mapping issues should not be ignored. For instance, the commercialised varenicline product is a ready package containing all required tablets for a defined period of use. Depending on the ETL design, the mapping in the database may be reflected as quantity equal as 1 package, or as 30 tablets.

Incidence and prevalence of use decreased for several drugs in 2020, after the COVID-19 pandemic onset, due to a global reduction in healthcare interactions, including diagnosis and prescriptions. However, some drugs showed an increment after 2021 that could be a rebound effect of health systems trying to catch up with the missing diagnostics and delayed treatments, or in case of short-term drugs, in particular antibiotics, due to an upsurge of respiratory infections. This can have an impact when trying to detect a suggested shortage by calculating the difference in incidence and prevalence 3 years before and after the shortage was announced.

The first and latest data points observed in each database may not contain the full year and therefore those values may be biased. Moreover, our results mostly rely on prescription rather than dispensation data. Doctors may have been prescribing unavailable drugs, which may result in a lag in the reduction of use (until the doctor switched to prescribing the available alternative) and/or as an overestimated incidence or prevalence of use when the shortage was active. Additionally, and as previously mentioned, some of the alternative medicines like nicotine are available over the counter, leading to an underestimation in incidence and prevalence in our data.

Different strategies adopted to mitigate the effects of the declared drug shortages may impact the data we are interpreting. As an example, in response to a shortage, a center may switch to an alternative drug while another is more restrictive in the selection of patients receiving the drug. The effect of the different mitigating strategies should be further studied as it can introduce and exacerbate inequalities in the access to proper care.

### Conclusions

Detection of shortages using estimates of incidence and prevalence of use can be challenging. We compiled and analysed data of annual incidence and prevalence of use for 57 medicines among 52 databases in Europe and the United States, including information on patient characteristics, indication, and dose, between 2010 and 2024. We detected shortages in at least one database regarding the incident and prevalent use of 8 and 9 drugs, respectively, out of the 15 drugs identified as having suggested shortages. Additionally, we observed a change in the users’ characteristics for several drugs. Federated network analysis provides an opportunity to obtain large amounts of information representing multiple countries and care settings, which can help in assessing and combatting against drug shortages. In this line, we have described timely real-world scenarios of drug shortages and those unobserved in various health care settings and countries which helps to better understand how drug shortages play out in real life.

## Supporting information

Annex I

## Data Availability

All data sources were previously mapped to the Observational Medical Outcomes Partnership (OMOP) Common Data Model (CDMv5). Thus, only aggregated data was shared by each of the data partners. Aggregated data is fully available in the two shiny apps of the study: https://dpa-pde-oxford.shinyapps.io/MegaStudy_IncidencePrevalence/ for incidence and prevalence results, and https://dpa-pde-oxford.shinyapps.io/MegaStudy_webinar_DUS/ for characterisation of users results. Access to patient level data are specific to each database and must be requested individually to each data source. Code of the study is fully available in GitHub: https://github.com/oxford-pharmacoepi/MegaStudy

https://dpa-pde-oxford.shinyapps.io/MegaStudy_IncidencePrevalence/

https://github.com/oxford-pharmacoepi/MegaStudy

## Acknowledgments

We acknowledge the immense effort and commitment of the EHDEN data partners, without whom this study would not have been possible. We also thank Montse Camprubi for her support coordinating the study.

## Authors’ roles

Conceptualisation: TB, MPM, DPA, PR. Formal analysis: TB, MPM. Execution of the code: all authors who contributed with data. Funding acquisition: PR, DPA. Data interpretation all authors. Writing original draft: MPM, TB, SS, PN, AR, KM, FSS, NR, AR, EF, JR, PP, MAM, LM, MF, AL, CL, AM, SR, LK, RC, SC, DP, EB, BP. Writing review and editing: all authors. Approving final version of manuscript: all authors.

## Ethical approval

All data partners received IRB approval or waiver in accordance with their institutional governance guidelines, which were collected by study coordinator. This included:

Use of Clinical Practice Research Datalink (CPRD) data for this study was approved via the Research Data Governance (RDG) Process of the UK Medicines and Healthcare Products Regulatory Agency (protocol 23_003389). Ethical approval for Fundación para la Investigación del Hospital Universitario La Fe de la Comunidad Valenciana (HULAFE) was obtained from the Comité de ética de la investigación con medicamentos (number 2024-561-1). Ethical approval for IMIM-Hospital del Mar Barcelona (IMASIS) was obtained by the Parc de Salut Mar Research Ethics Committee CEIm-Parc de Salut Mar (number 2023/11266). Ethical approval number for Hospital District of Helsinki and Uusimaa (HUS) in this study was HUS/325/2023. Ethical approval number for Istanbul University (ITF) was 2302692. Ethical approval number for Netherlands Comprehensive Cancer Organisation (NCR) was K23.338. Ethical approval number for University Medicine Dresden (UM DRESDEN) was SR-EK-181052024. Ethical approval number for Optimum Patient Care Limited (OPCRD) was ADEPT0624. Ethical approval number for Clinical-hospital center Zvezdara (CHCZ) was FWA00024180. Ethical approval number for Fundació Institut d’Investigació Sanitària Illes Balears (PRISIB) was CI-866-24. Ethical approval number for EGAS MONIZ HEALTH ALLIANCE (EMHA ULSRA and EMHA ULSGE) was 4/2023 Ref 19-CE-ICVS/CAC-EMHA.

Ethical approval number for FISABIO-HSRU (VID-CONSIGN) was 2023/382. Ethical approval number for University Hospital Antwerp (UZA) was 5957-EDGE 3372-BUN. Ethical approval number for AZ Groeninge (AZGR) was AZGD2024008. Ethical approval number for Viecuri was 2022_088. Ethical approval number for Charité - Universitätsmedizin (CHA CAN) was EA1/04124. Ethical approval number for THIN databases was 2024-06-R. Ethical approval number for Galilee Medical Center (GMC) and Bnai Zion Medical Center (BZMC) was BNZ0077-22. Ethical approval number for Semmelweis University Clinical Database (SUCD) was BM/14889-1/2024. Ethical approval for The Integrated Primary Care Information Project (IPCI) was obtained by the Integrated Primary Care Information review board (registration number 9/2023).

The remaining databases needed no approval for use of pseudo anonymised secondary data.

