## Supplementary for "Trends of use of drugs with suggested shortages and their alternatives across 52 real world data sources and 18 countries in Europe and North America"

**Supplementary Tables**

**Supplementary Table 1. List of drugs of interest included in the study.**

| Drug of interest | date shortage announced | alternative | shortage resolved | date shortage resolved |
| --- | --- | --- | --- | --- |
| Alteplase,  Tenecteplase | 23/09/2022 | Streptokinase, urokinase | YES | 28/08/2024 |
| Sarilumab | 09/11/2021 | Tocilizumab, abatacept, tofacitinib, baricitinib, upadacitinib, etanercept, infliximab, certolizumab, golimumab, anakinra | YES | 14/09/2023 |
| Tocilizumab | 03/09/2021 | Sarilumab,abatacept, tofacitinib, baricitinib, upadacitinib, etanercept, infliximab, certolizumab, golimumab, anakinra | NO |  |
| Verteporfin | 08/11/2021 | Ranibizumab, bevacizumab | NO |  |
| Varenicline | 08/07/2021 | Nicotine | NO |  |
| C1 esterase inhibitor | 1) 29/06/2017  2) 06/08/2018 | Icatibant, ecallantide, conestat alfa, lanadelumab, berotralstat | YES | 1) 06/11/2017  2) 25/06/2020 |
| Arsenic trioxide | 31/07/2017 | Cytarabine liposomal, cytarabine any, daunorubicin, cytarabine plus daunorubicin, idarubicin, tretinoin oral | YES | 06/09/2018 |
| Belatacept | 17/03/2017 | Mycophenolic acid, sirolimus, cyclosporine, tacrolimus no topical | NO |  |
| Ganirelix | 16/02/2017 | Cetrorelix | YES | 08/11/2022 |
| Cetrorelix^1^ | 10/08/2022 | Ganirelix | YES | 15/02/2023 |
| Imiglucerase | 04/11/2013 | Velaglucerase alfa, taliglucerase alfa | YES | 01/10/2018 |
| Agalsidase beta | 04/11/2013 | Agalsidase alfa | YES | 07/06/2016 |
| Tigecycline | 22/05/2015 | Meropenem, piperacillin plus tazobactam, ceftozolane plus tazobactam | YES | 07/11/2019 |
| Amoxicillin, amoxicillin plus clavulanate | 27/01/2023 | Azithromycin, clarithromycin, phenoxymethylpenicillin, benzylpenicillin | NO |  |
| For completeness, the antibiotics ceftriaxone, cefotaxime, and cefuroxime were assessed despite that no shortage was announced  ^1^ Shortage of cetrorelix was shorter than 365 days, thus, it was included in the study as an alternative. | | | | |

**Supplementary Table 2. Number of drugs in shortage by country with a ≥33% drop in incidence or prevalence.**

|  | Incidence results | | | Prevalence results | | |
| --- | --- | --- | --- | --- | --- | --- |
| Country | Number of observed drugs | Number of drugs with ≥33% decrease in use | Percentage of number of drugs in shortage vs number observed | Number of observed drugs | Number of drugs with ≥33% decrease in use | Percentage of number of drugs in shortage vs number observed |
| Belgium | 4 | 2 | 50% | 5 | 5 | 100% |
| Estonia | 1 | 1 | 100% | 1 | 1 | 100% |
| Finland | 7 | 2 | 29% | 8 | 1 | 12% |
| France | 8 | 1 | 12% | 7 | 3 | 43% |
| Germany | 9 | 2 | 22% | 10 | 3 | 30% |
| Greece | 3 | 1 | 33% | 2 | 1 | 50% |
| Hungary | 1 | 0 | 0% | 1 | 0 | 0% |
| Israel | 2 | 0 | 0% | 2 | 0 | 0% |
| Italy | 4 | 2 | 50% | 3 | 2 | 67% |
| Netherlands | 6 | 1 | 17% | 6 | 1 | 17% |
| Portugal | 5 | 1 | 20% | 6 | 2 | 33% |
| Romania | 2 | 0 | 0% | 2 | 2 | 100% |
| Serbia | 1 | 0 | 0% | 1 | 0 | 0% |
| Spain | 9 | 3 | 33% | 8 | 4 | 50% |
| Sweden | 5 | 0 | 0% | 6 | 1 | 17% |
| Türkiye | 0 | 0 | 0% | 0 | 0 | 0% |
| UK | 5 | 3 | 60% | 6 | 3 | 50% |
| USA | 13 | 3 | 23% | 13 | 7 | 54% |

**Supplementary Figures**

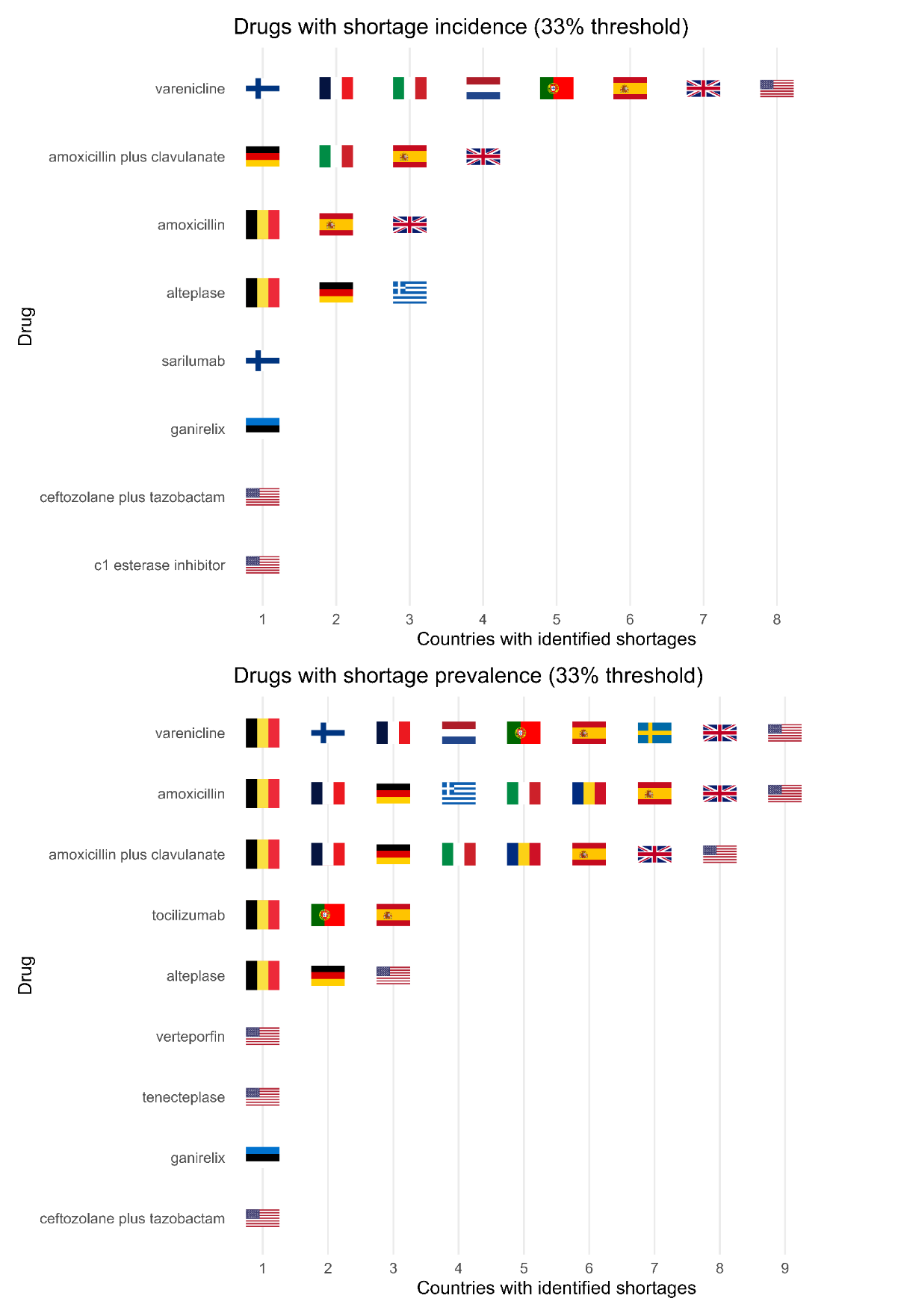

**Supplementary Figure 1. Drugs in shortages and countries where at least one database presented a ≥33% drop in A) incident use or B) prevalent use of the drug.** To be considered as drug in shortage, at least one database of the study in the country had to present a ≥33% drop in use (in incidence or in prevalence). When any of the databases in the country show a ≥33% drop in use after the shortage announcement, we considered it as no shortage observed in the data.

A) Incident use of varenicline and nicotine

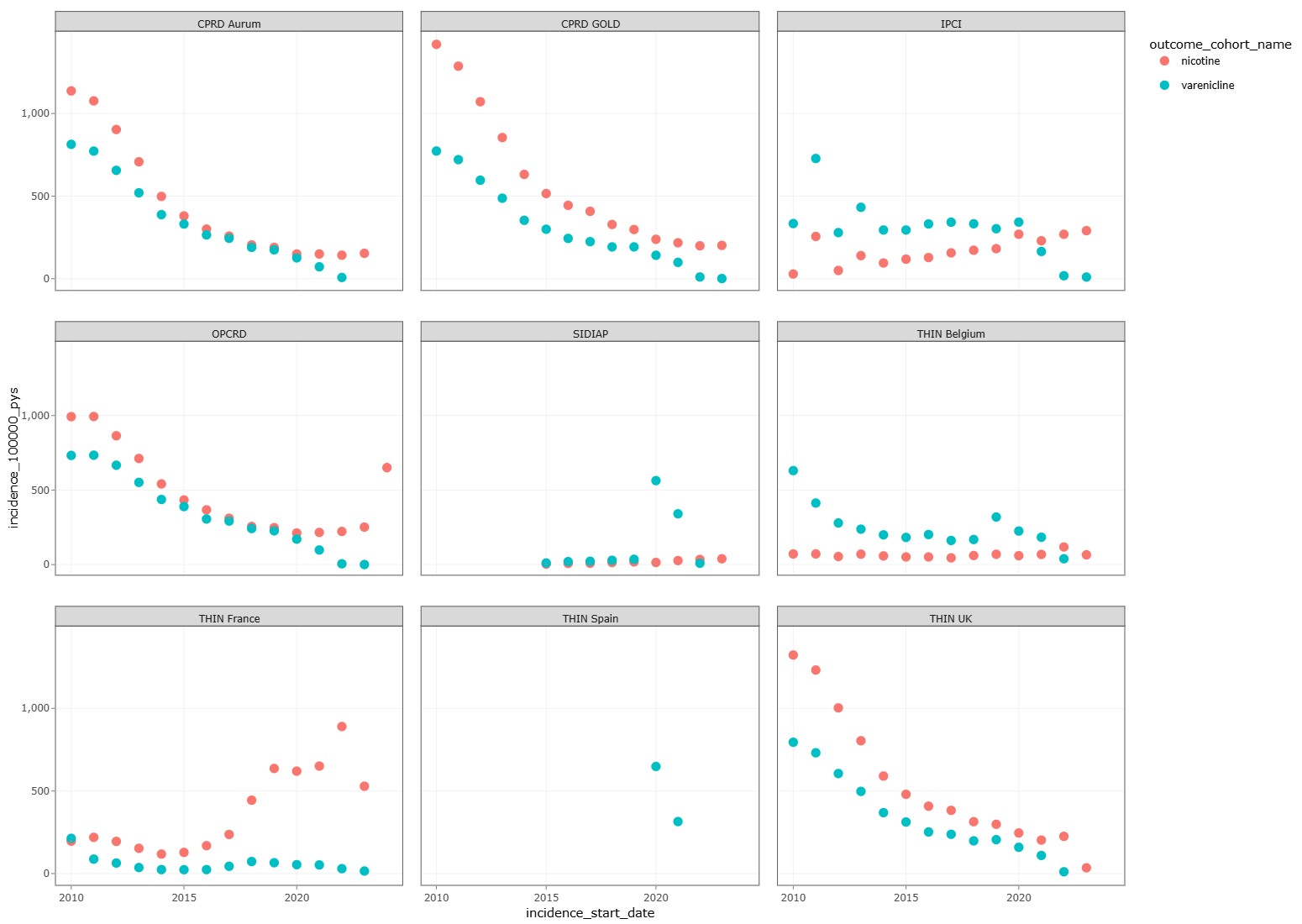

B) Prevalent use of varenicline and nicotine

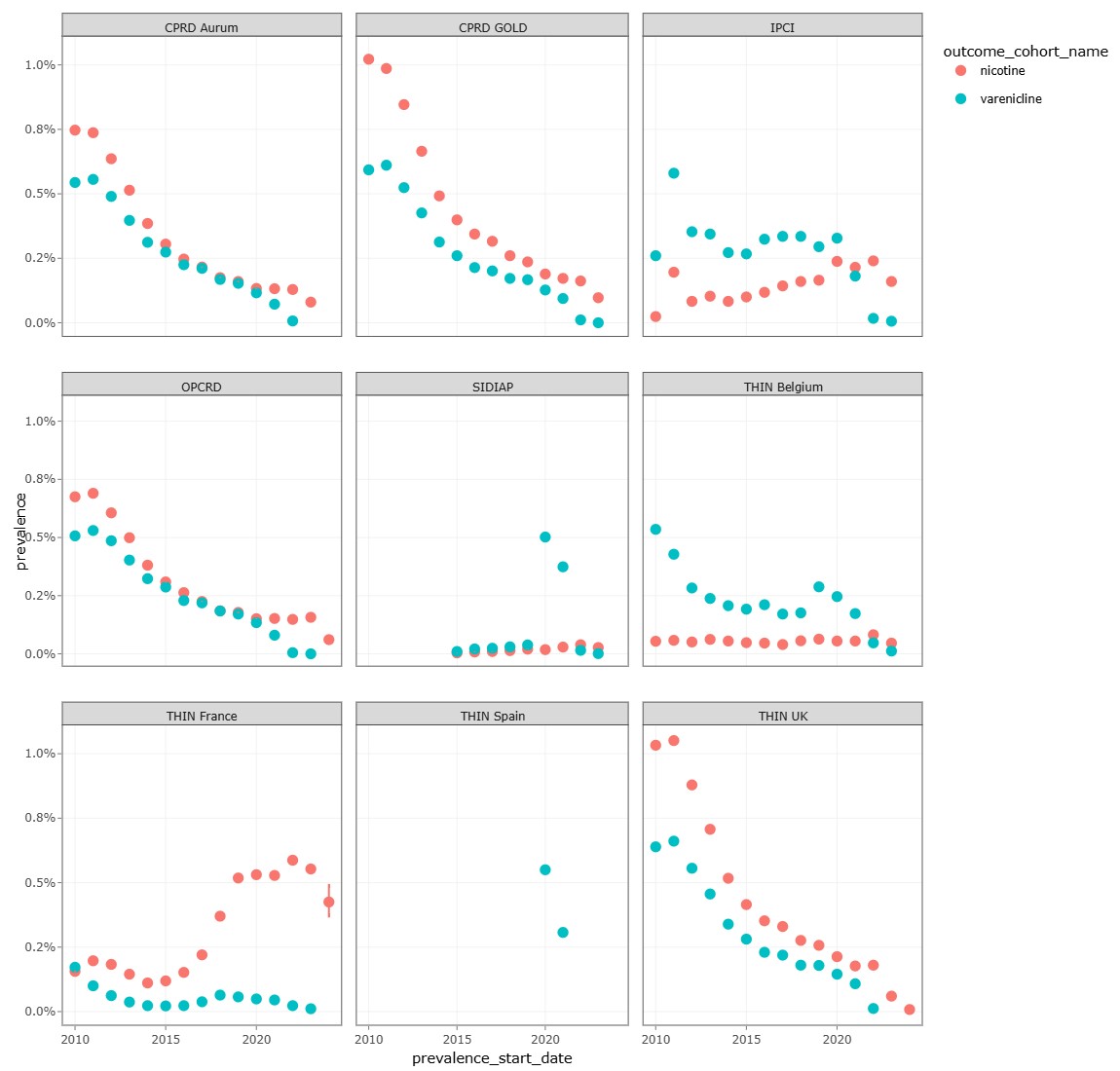

**Supplementary Figure 2. Use of varenicline and its alternative, nicotine supplements, in primary care: A) Incidence rates of use per 100,000 person-years and B) Prevalence of use.**

A) Primary care databases

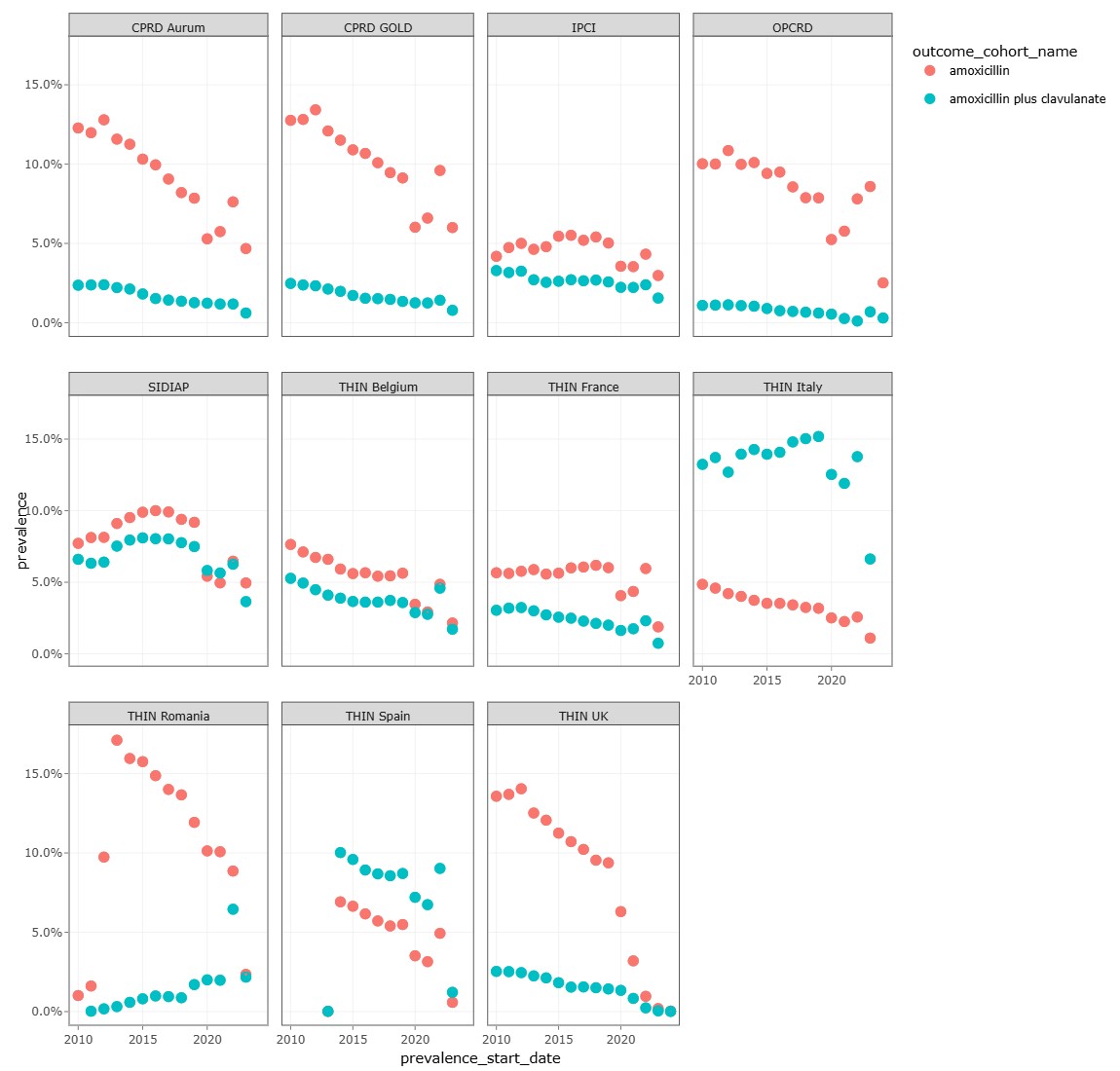

B) Secondary care databases

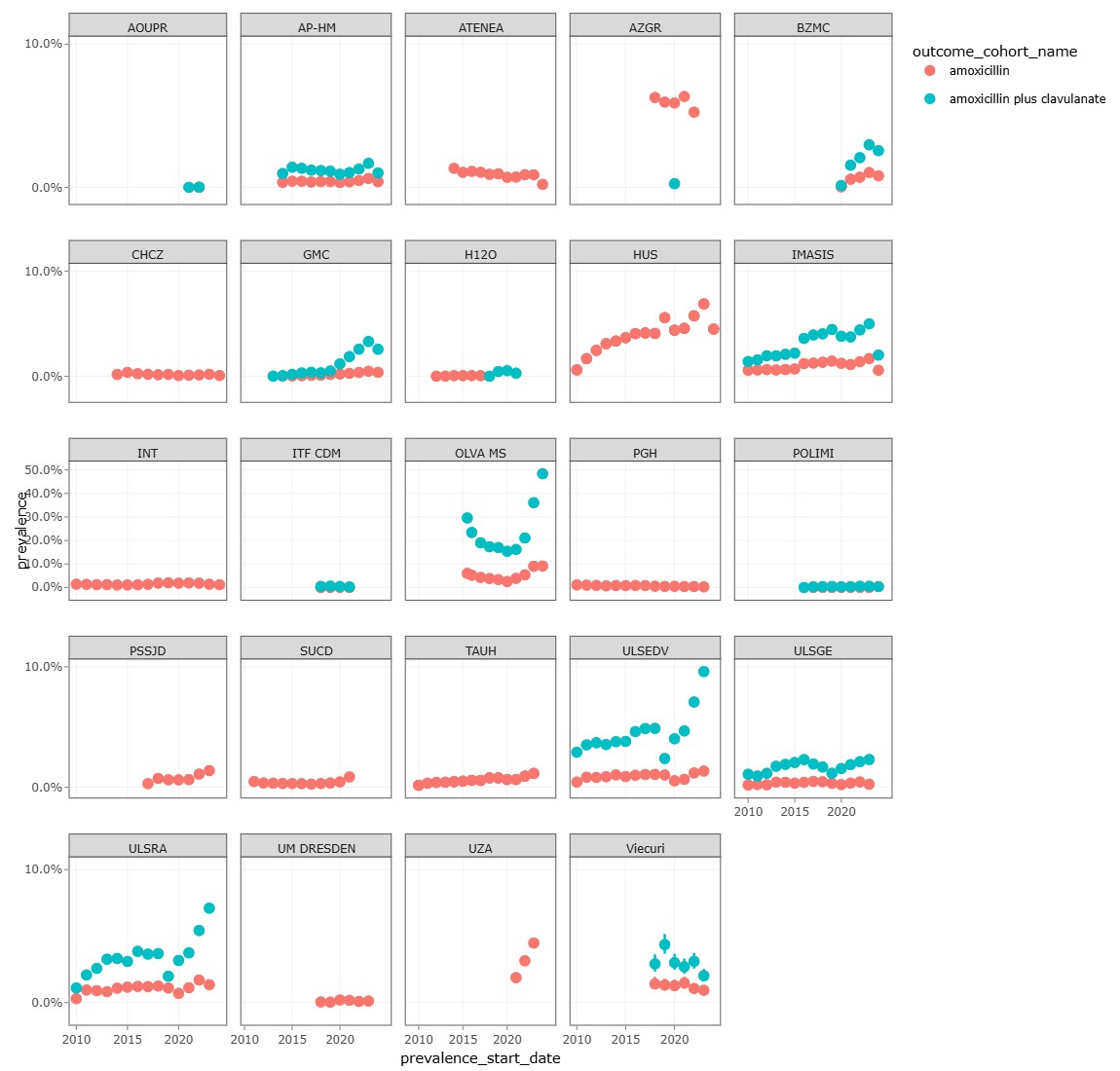

C) Other types of databases: containing primary and secondary care combined

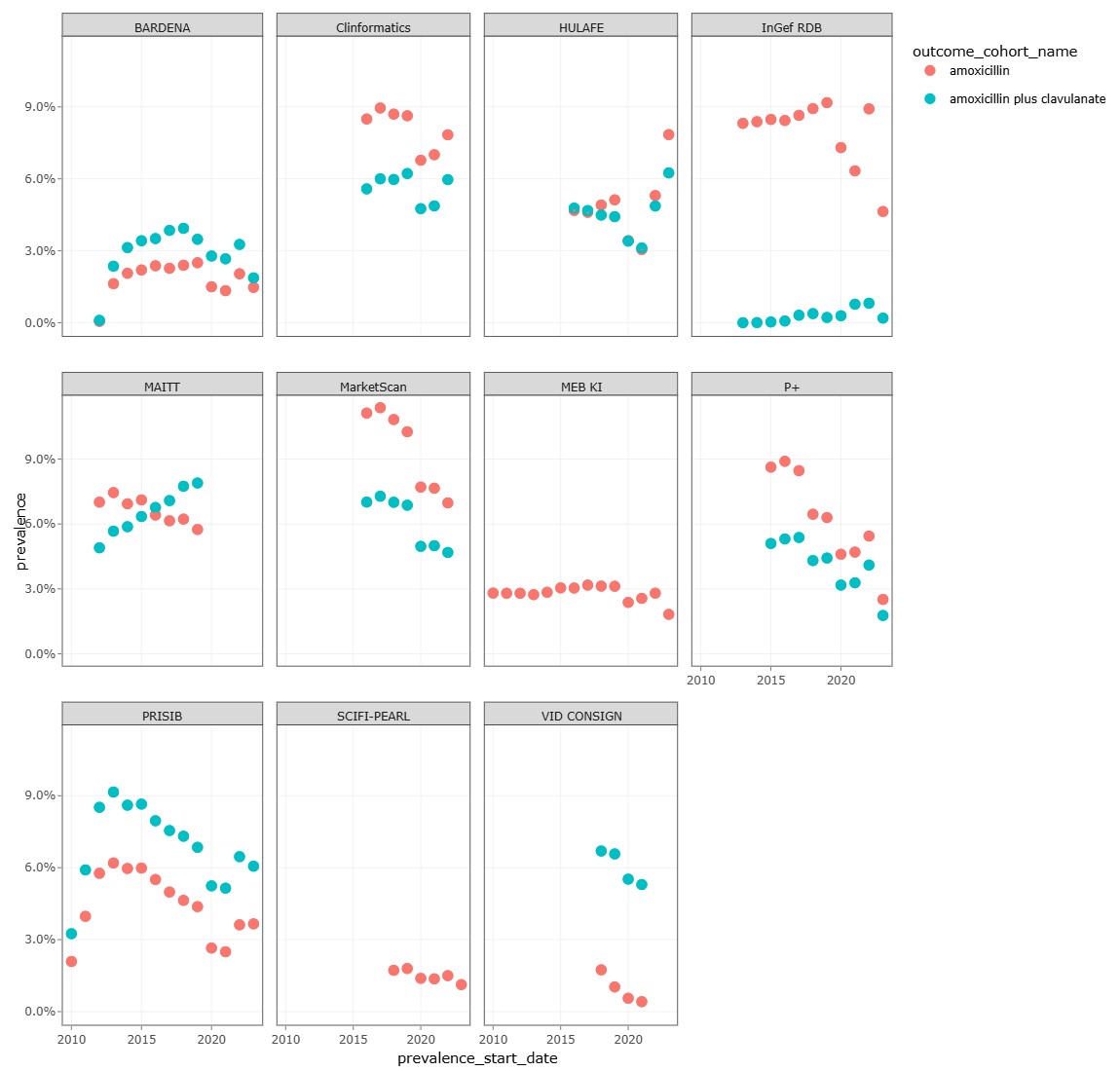

**Supplementary Figure 3. Prevalence of use of amoxicillin alone and in combination with clavulanate in A) Primary care, B) Secondary care and C) Other types of databases.** Claims databases contain records from primary and secondary care data. Secondary care databases shown are all hospitals.

A) Primary care databases

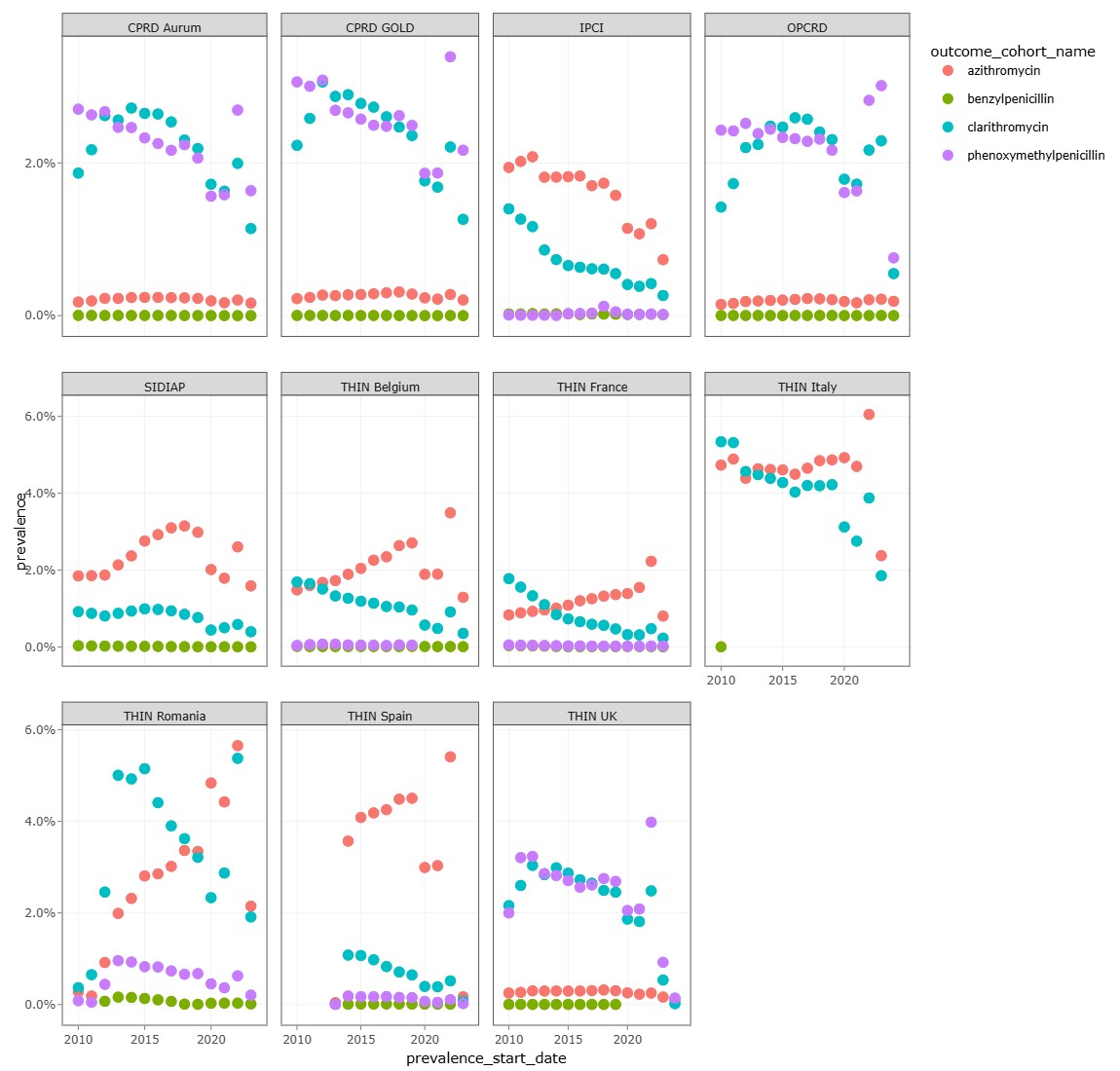

B) Secondary care databases

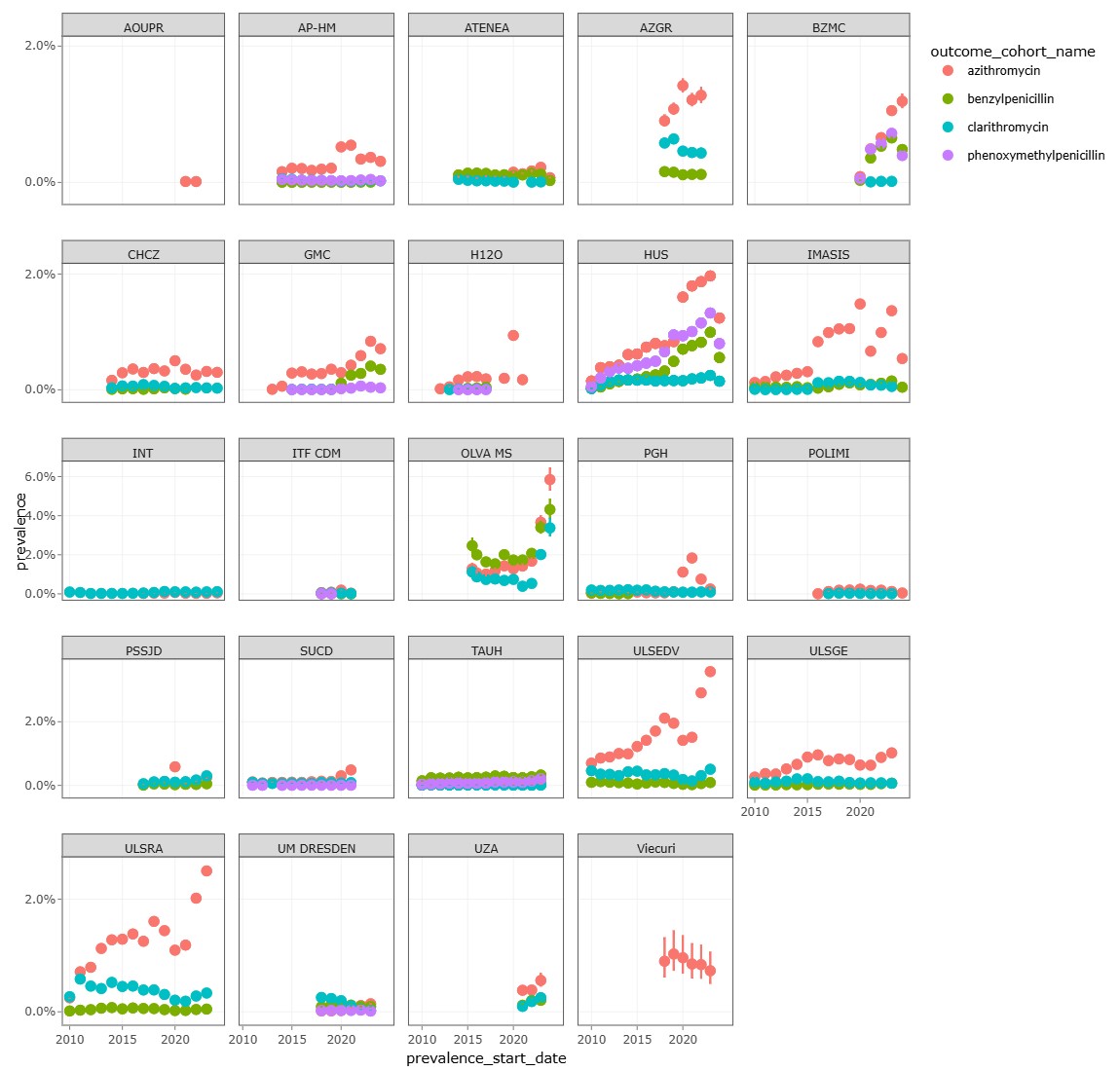

C) Other types of databases: containing primary and secondary care combined

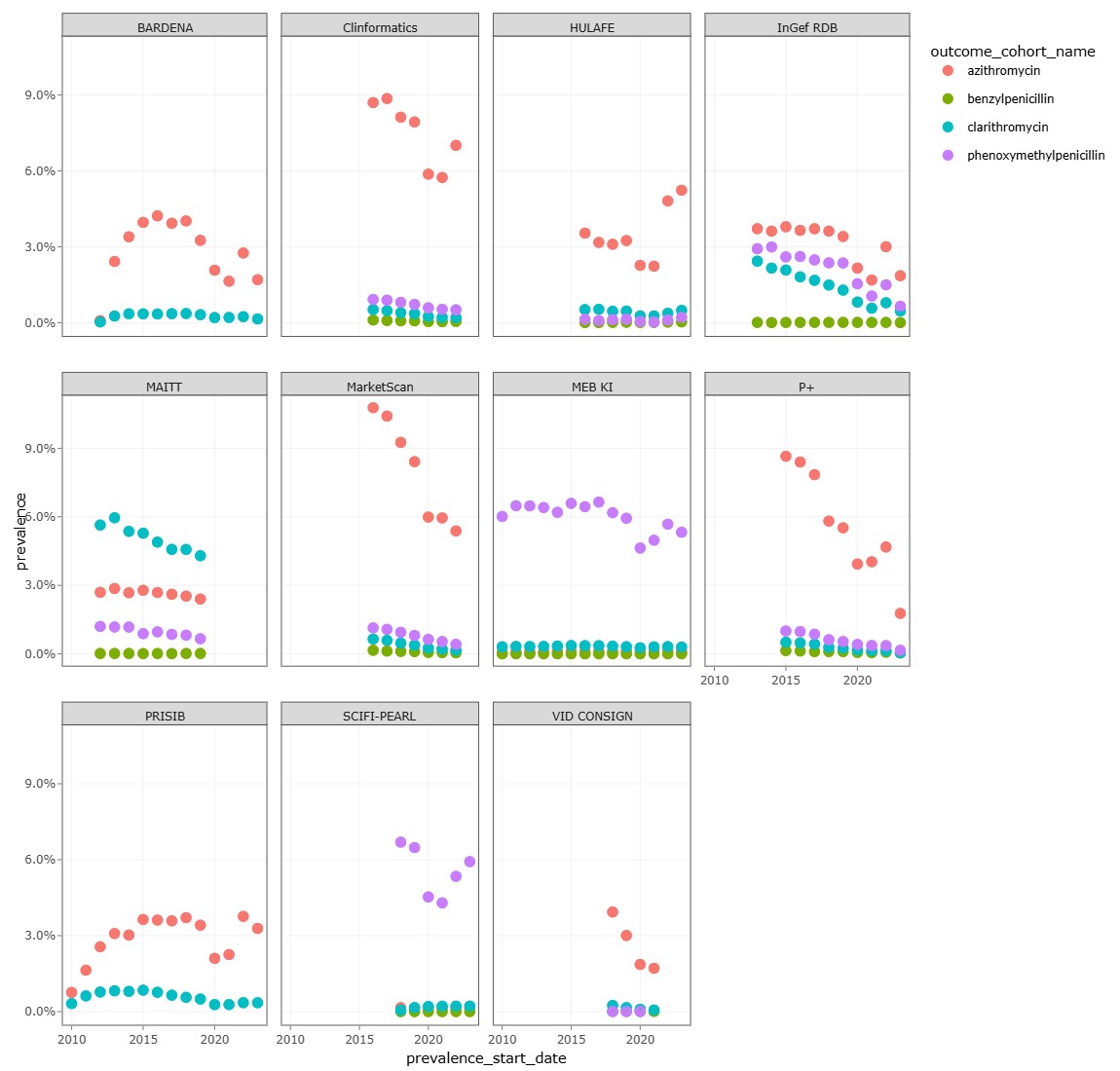

**Supplementary Figure 4. Prevalence of alternative antibiotics for common infections.** Claims databases contain records from primary and secondary care data. Secondary care databases shown are all hospitals.

A) Observed changes in the proportion of sex in incident users after the shortage date

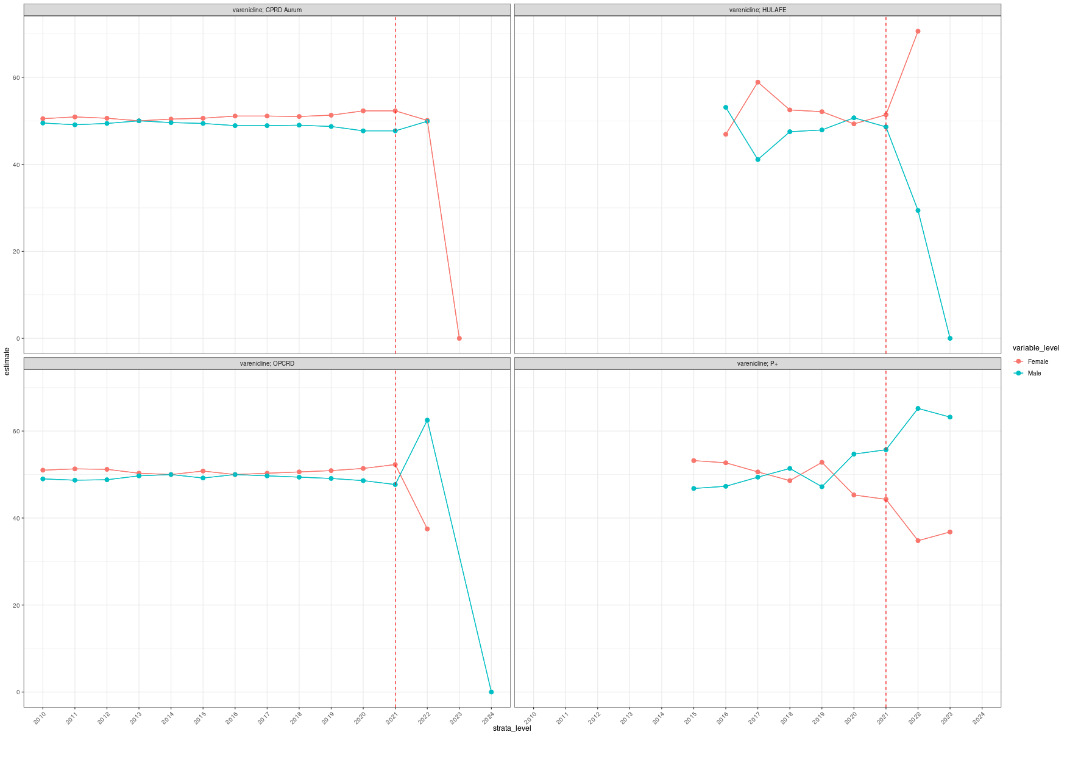

B) Observed changes in the proportion of sex in prevalent users after the shortage date

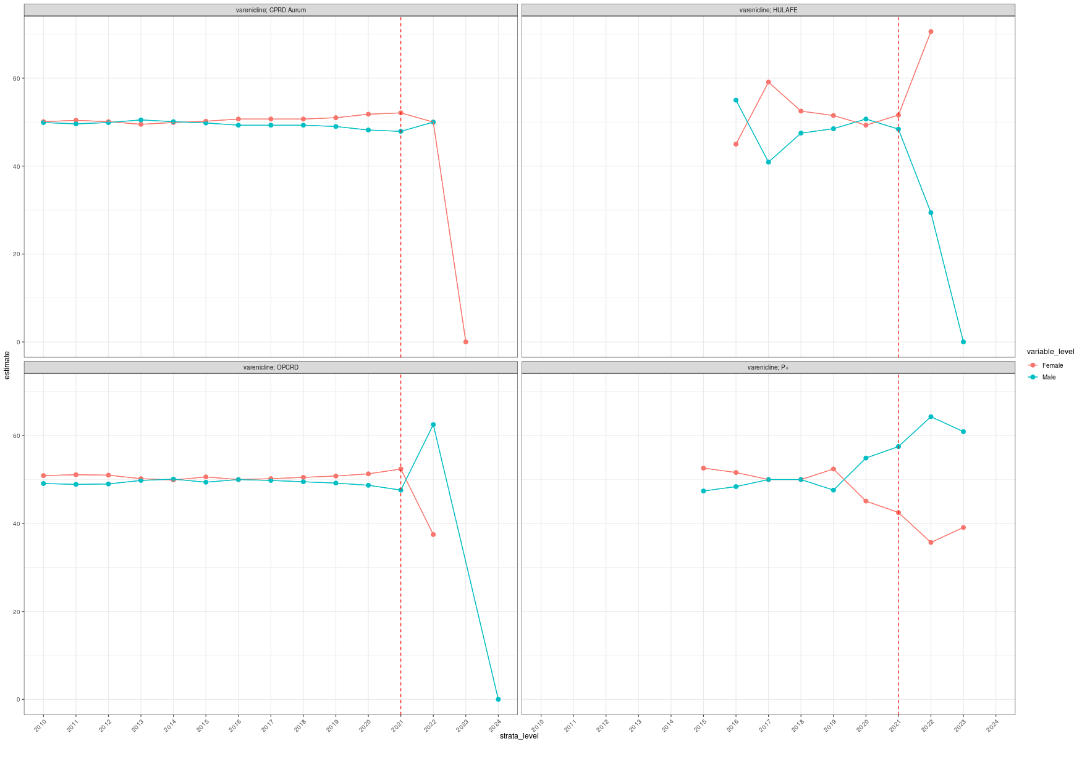

C) Observed changes in the cumulative dose of incident users after the shortage date

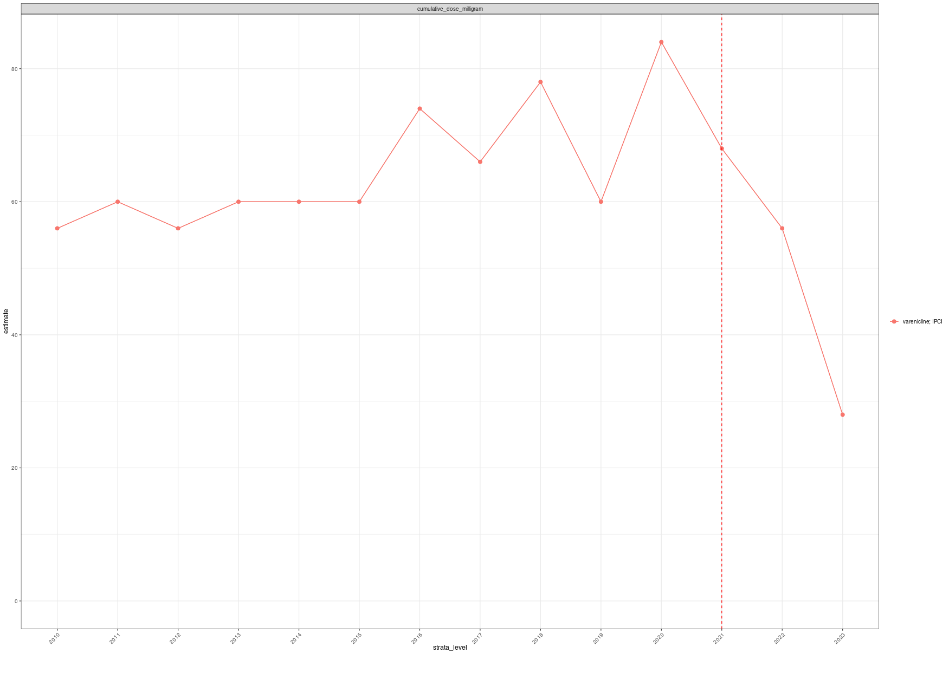

D) Observed changes in the cumulative dose of prevalent users after the shortage date

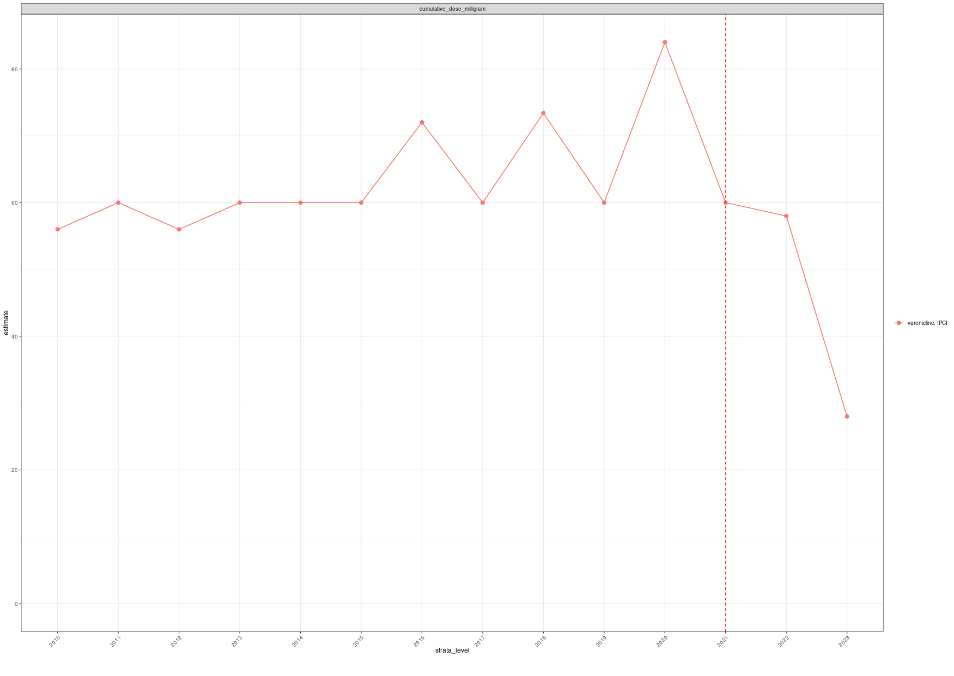

E) Observed changes in duration after the shortage date in incident users

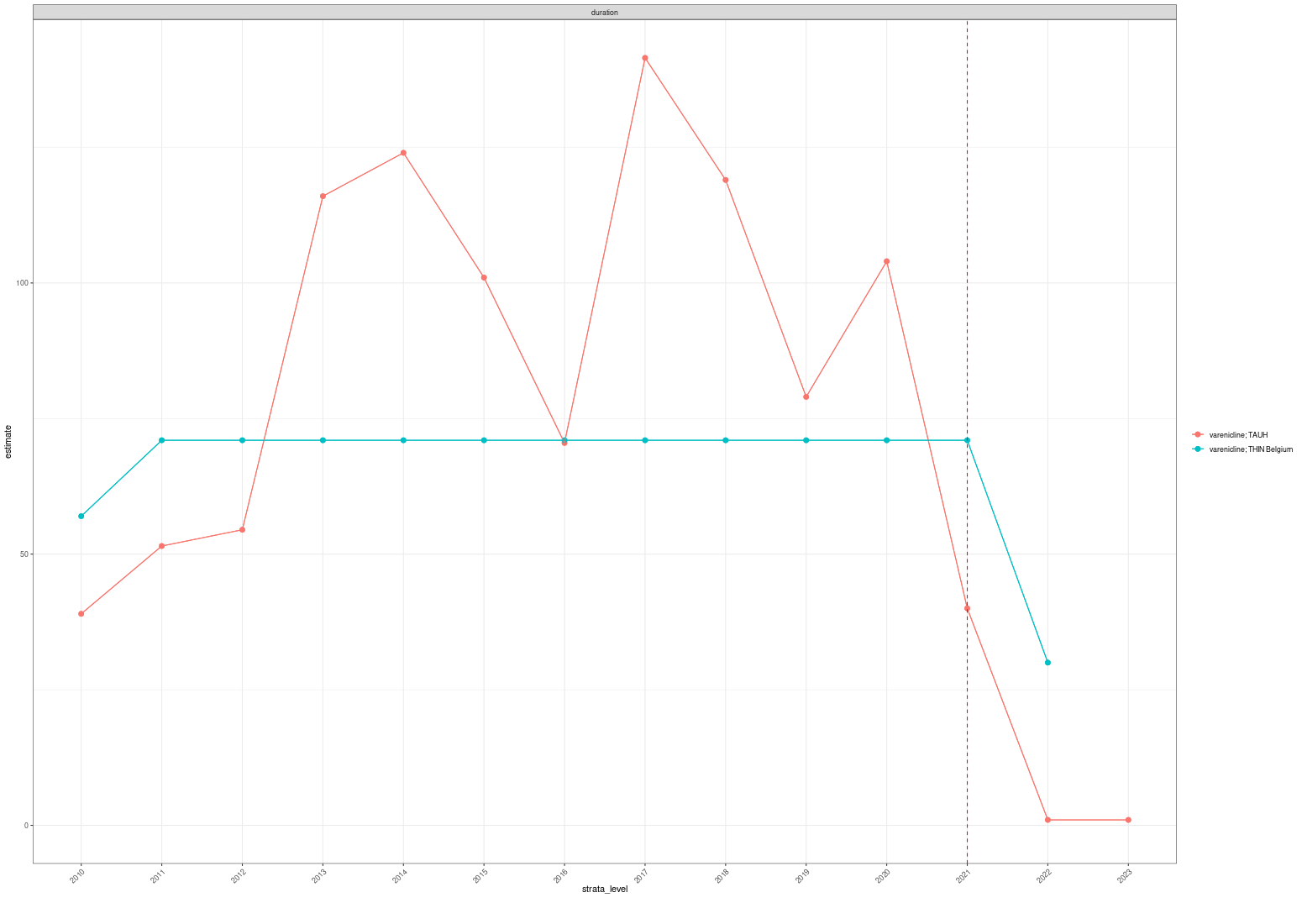

F) Observed changes in duration after the shortage date in prevalent users

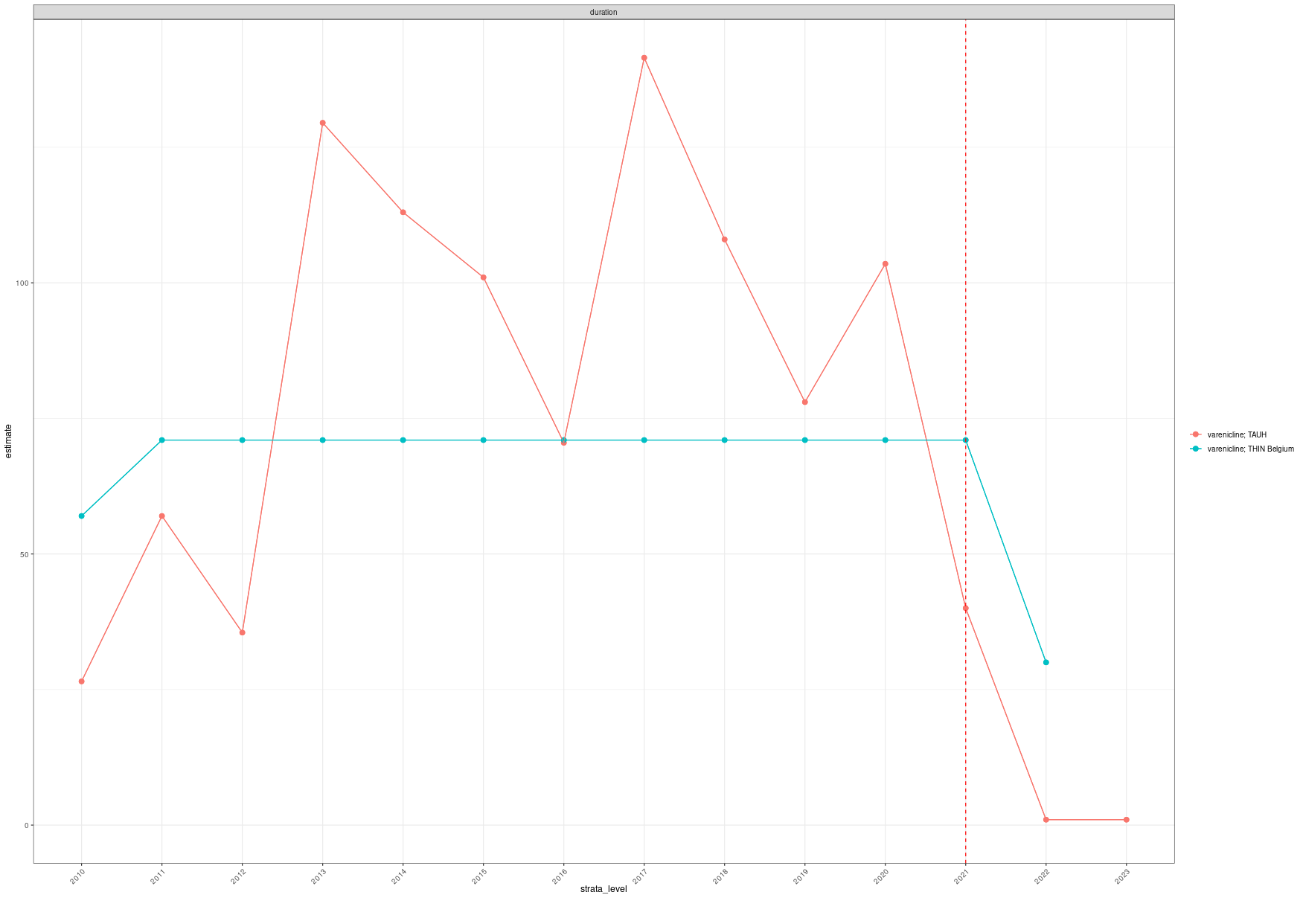

G) Observed changes in initial quantity after the shortage date in incident users

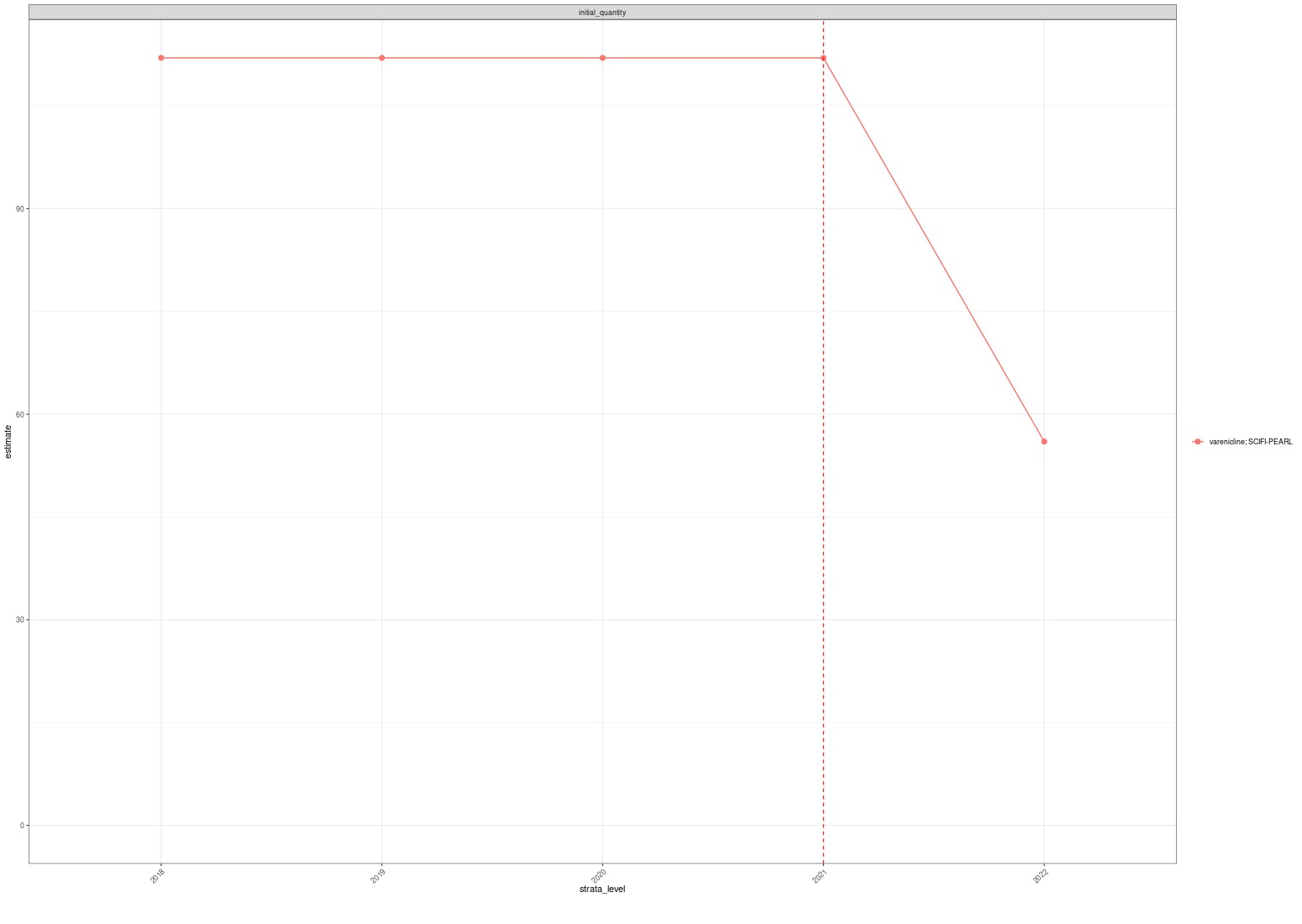

H) Observed changes in initial quantity after the shortage date in prevalent users

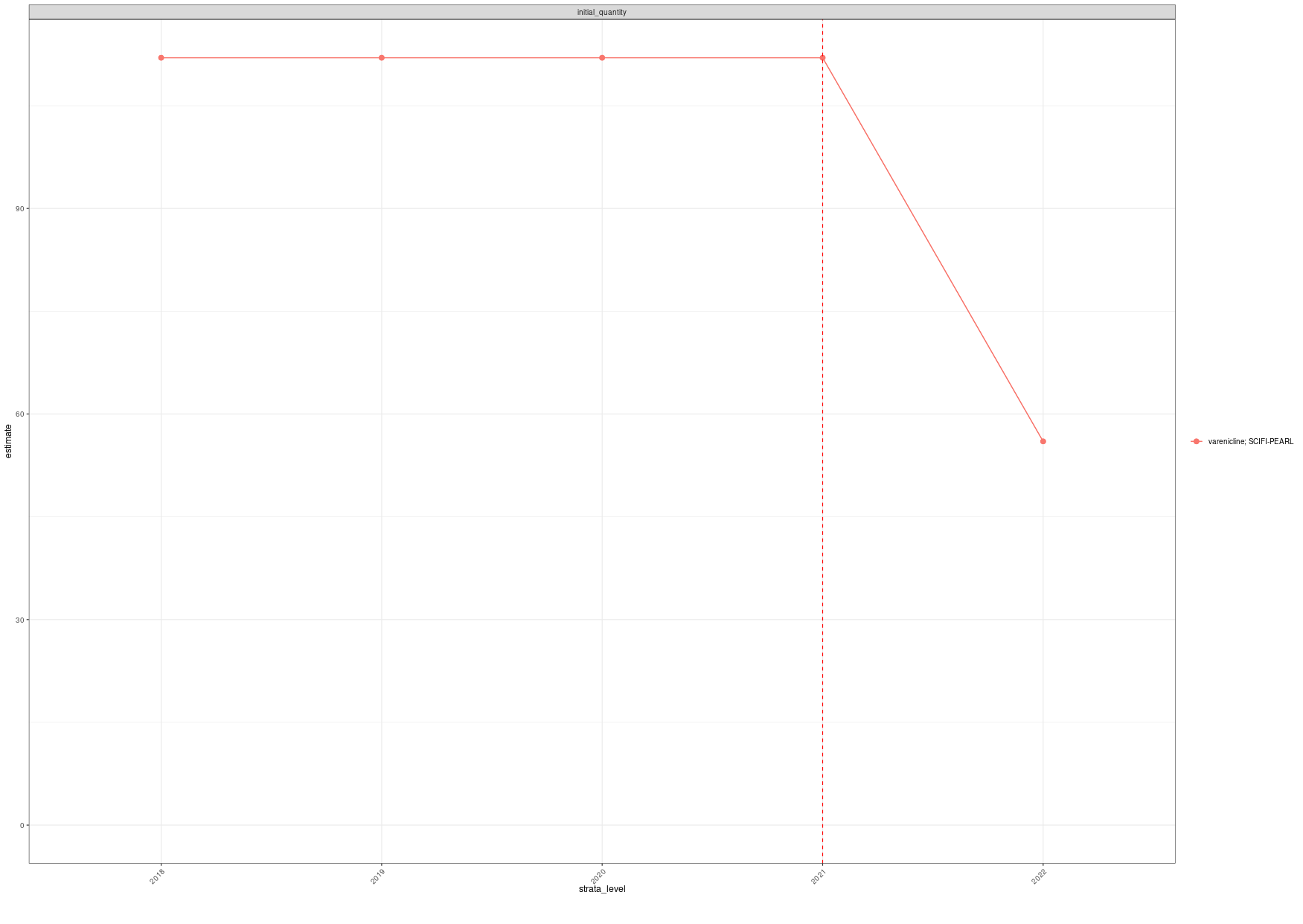

**Supplementary Figure 5. Observed changes in large-scale patient-level drug utilisation study of varenicline: Observed changes in the proportion of sex in A) incident users and B) prevalent users; Observed changes in the cumulative dose of C) incident users and D) prevalent users; Observed changes in duration after the shortage date in E) incident users and F) prevalent users; and Observed changes in initial quantity after the shortage date in G) incident users and H) prevalent users.** The red dotted line indicates the year of shortage of each drug of interest.

A) Observed changes in the median age of incident users after the shortage date

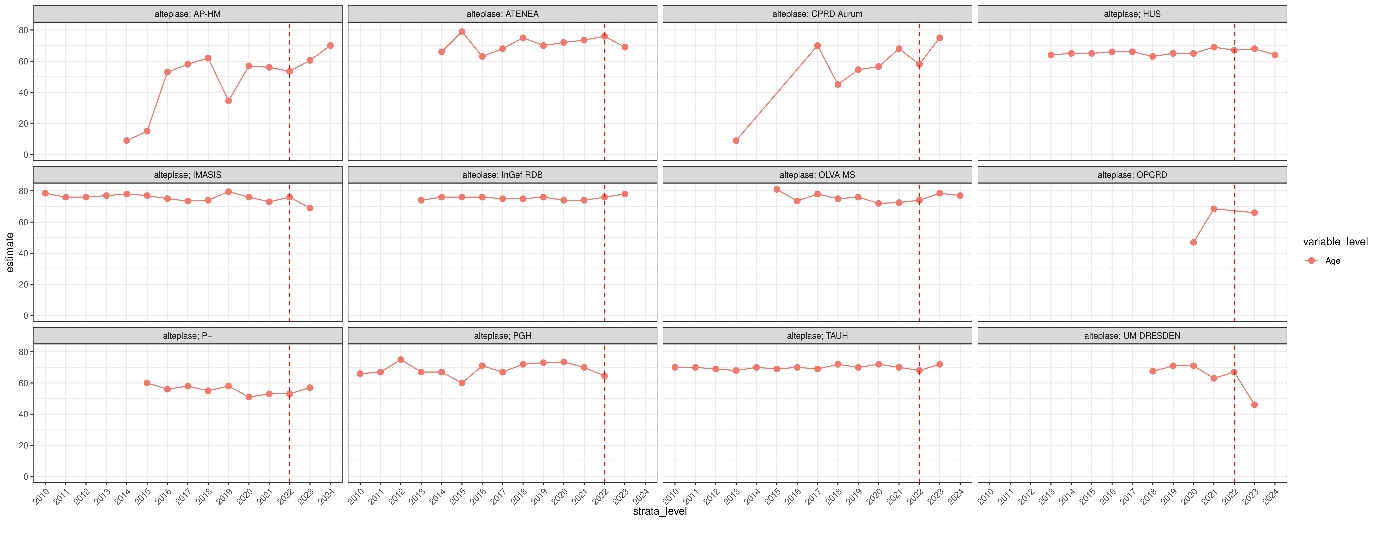

B) Observed changes in proportion of age groups of incident users after the shortage date

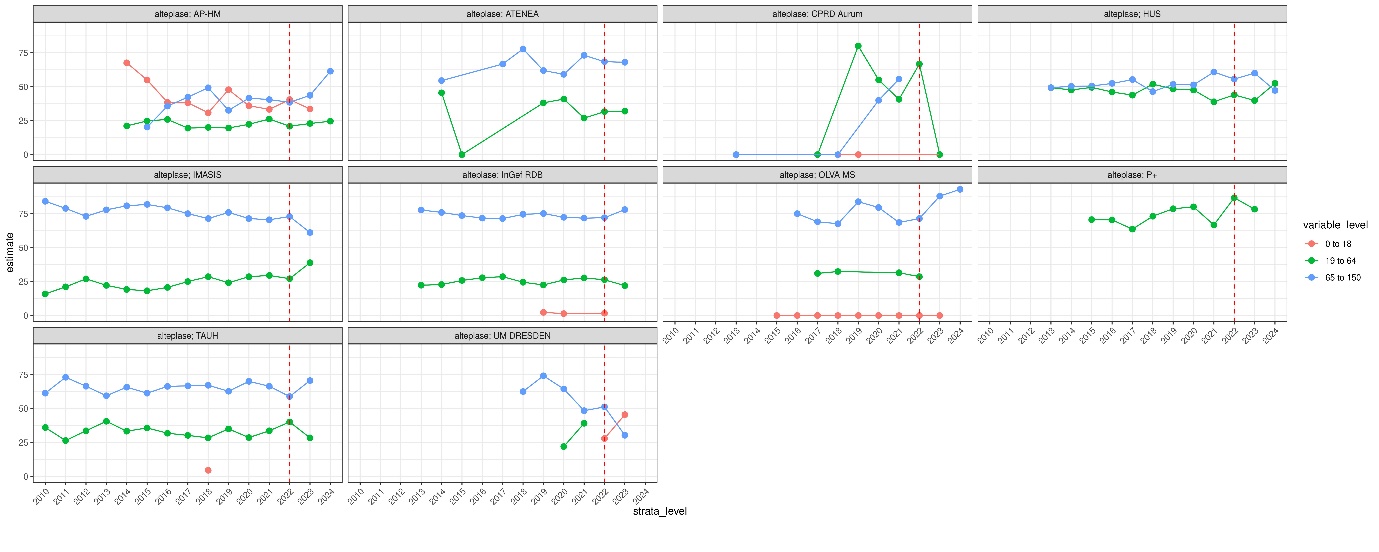

C) Observed changes in proportion of sex of incident users after the shortage date
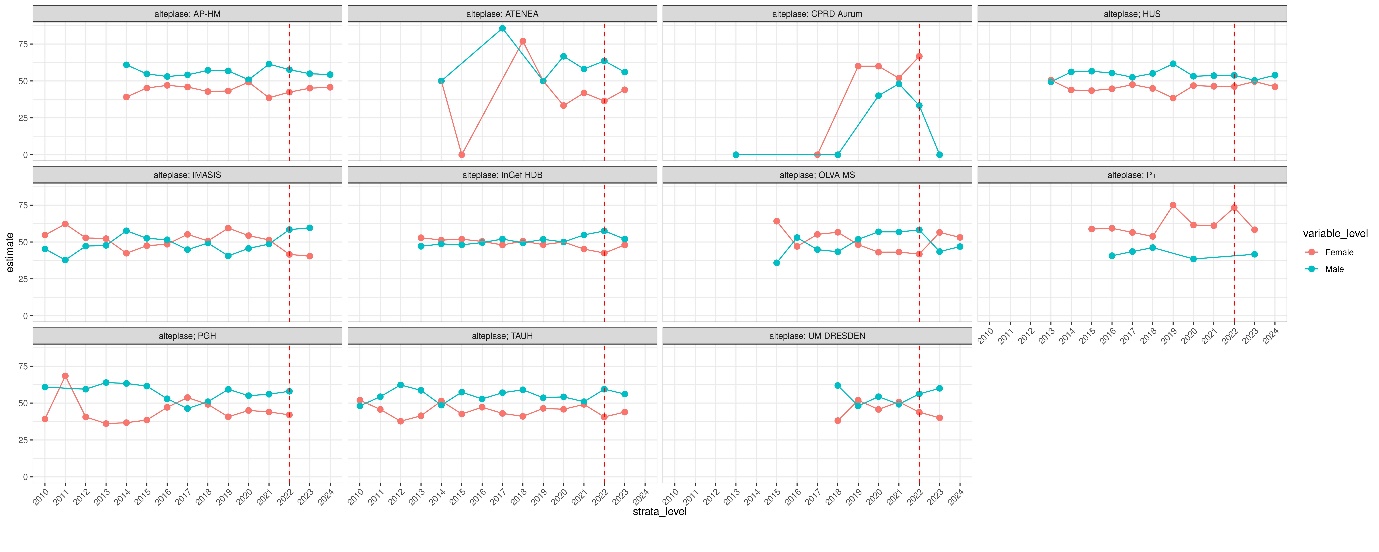

D) Observed changes in the proportion of sex in prevalent users after the shortage date

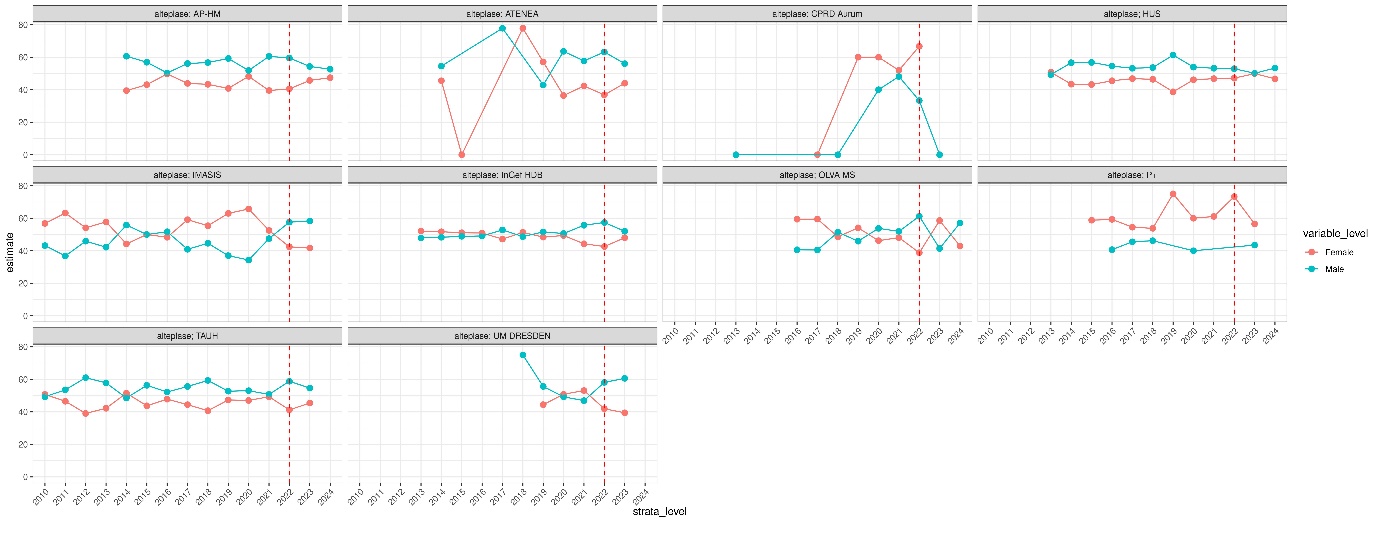

E) Indication for pulmonary embolism 30 days before incident prescription of alteplase

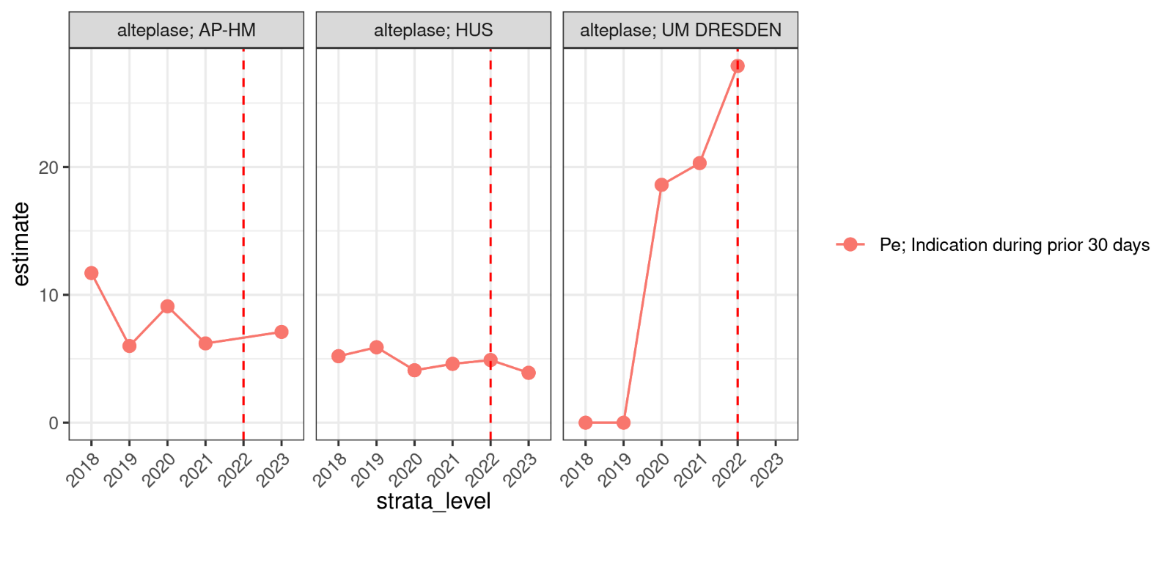

F) Indication for pulmonary embolism any time before incident prescription of alteplase

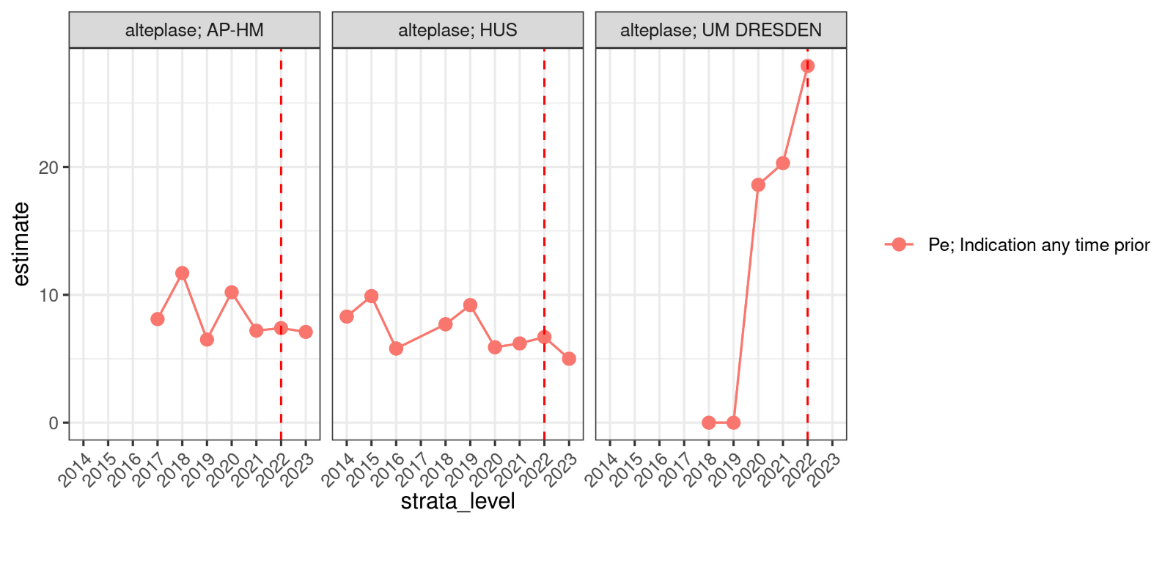

G) Indication for pulmonary embolism at index date before prevalent prescription of alteplase

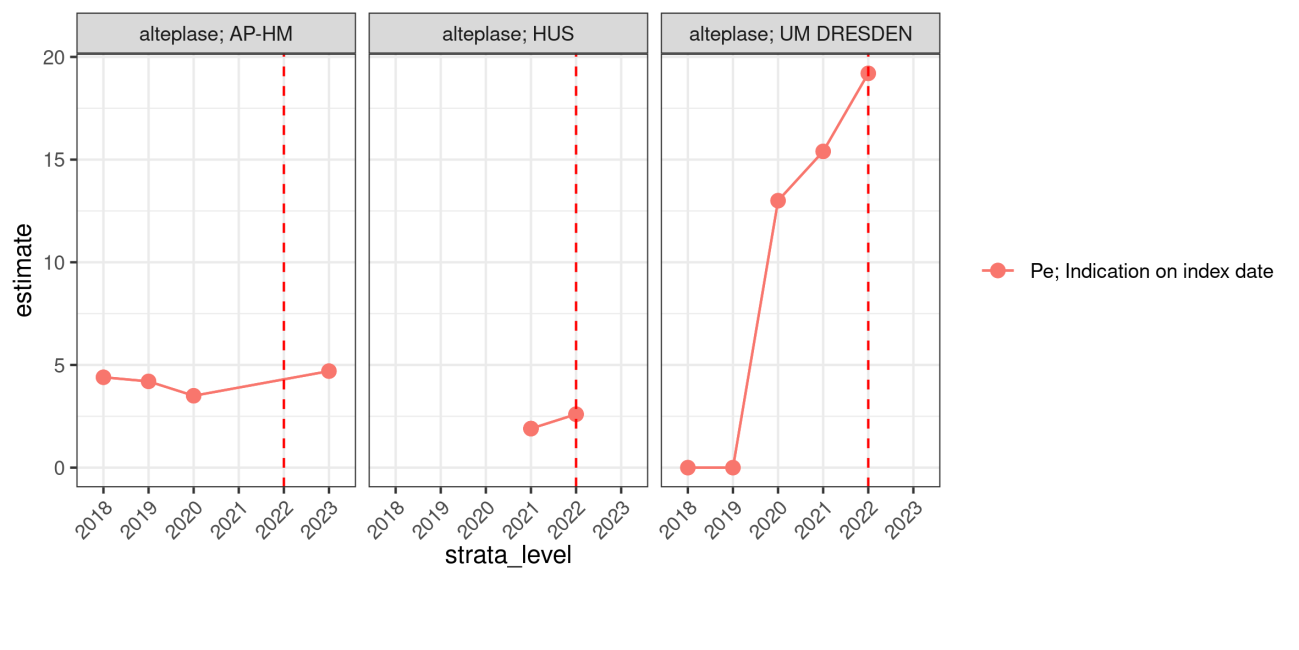

H) Indication for pulmonary embolism 30 days before prevalent prescription of alteplase

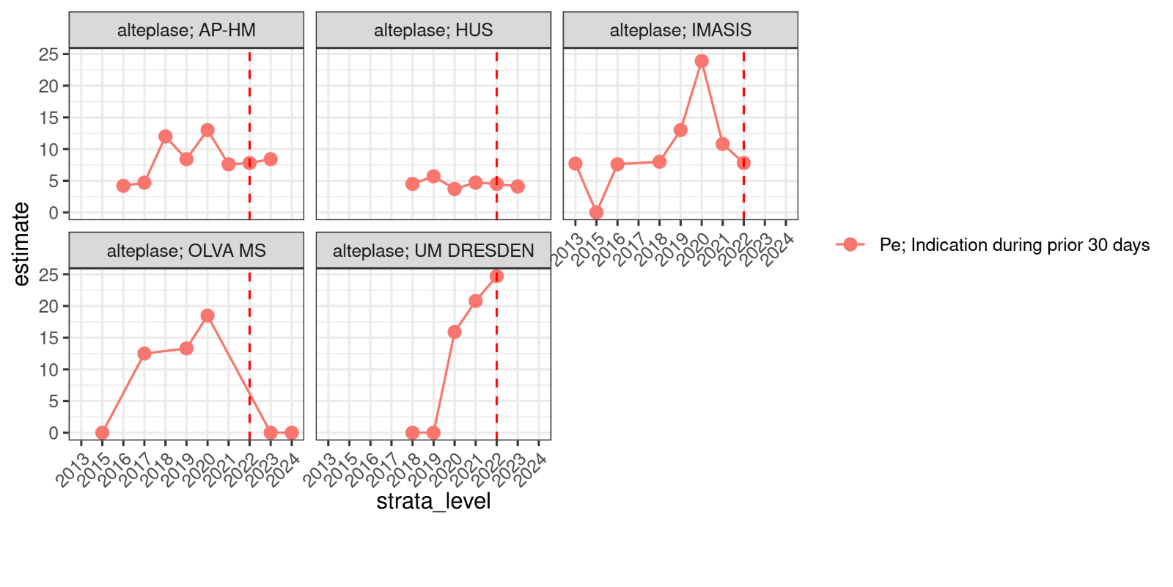

I) Indication for pulmonary embolism any time before prevalent prescription of alteplase

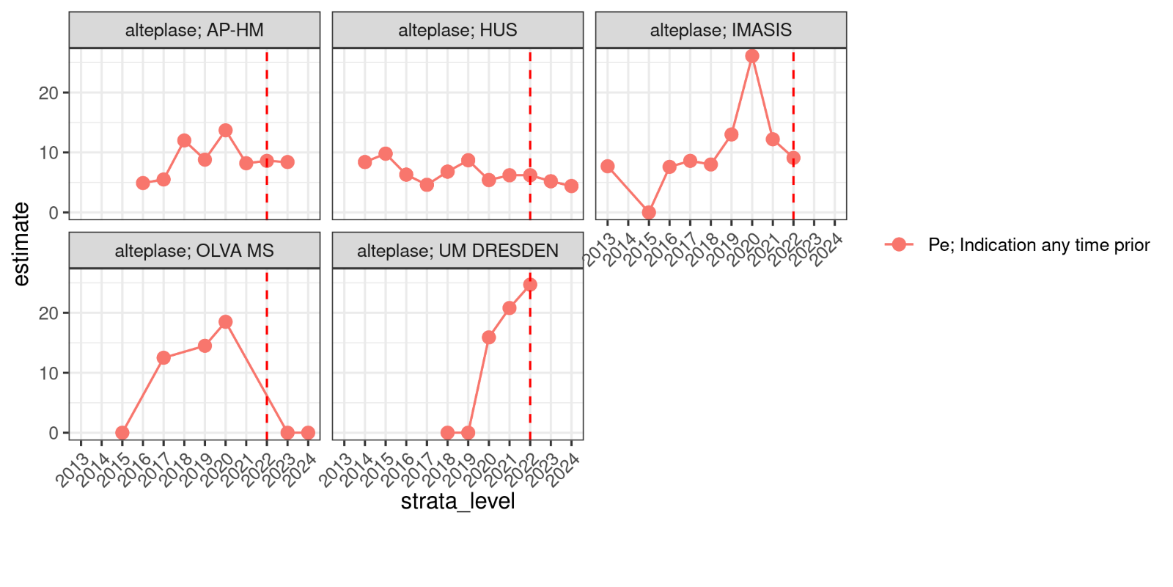

**Supplementary Figure 6. Observed changes in large-scale patient-level drug utilisation study of alteplase: Observed changes among incident users after the shortage date in A) the median age, B) age groups and C) proportion of sex; D) Observed changes among proportion of sex in prevalent users; Indication for pulmonary embolism E) 30 days or F) any time before incident prescription of alteplase; Indication for pulmonary embolism G) at index date, H) 30 days before or I) any time before prevalent prescription of alteplase.** The red dotted line indicates the year of shortage of each drug of interest. Abbreviations: PE, pulmonary embolism.

A) Observed changes in the median age of incident users after the shortage date

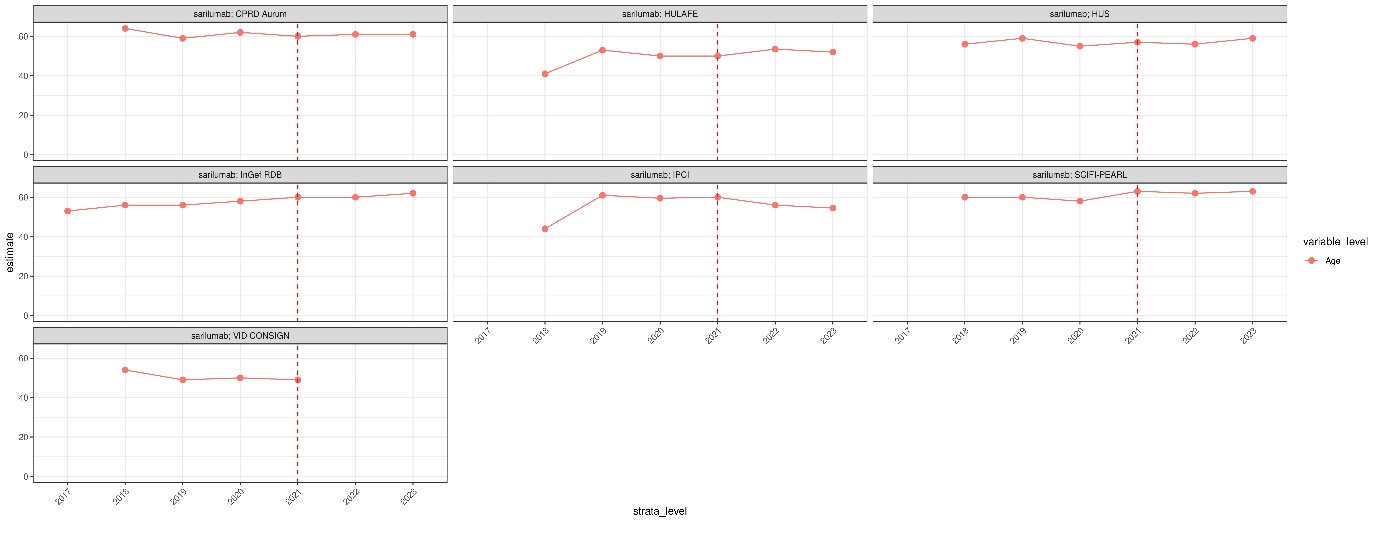

B) Observed changes in proportion of age groups of incident users after the shortage date

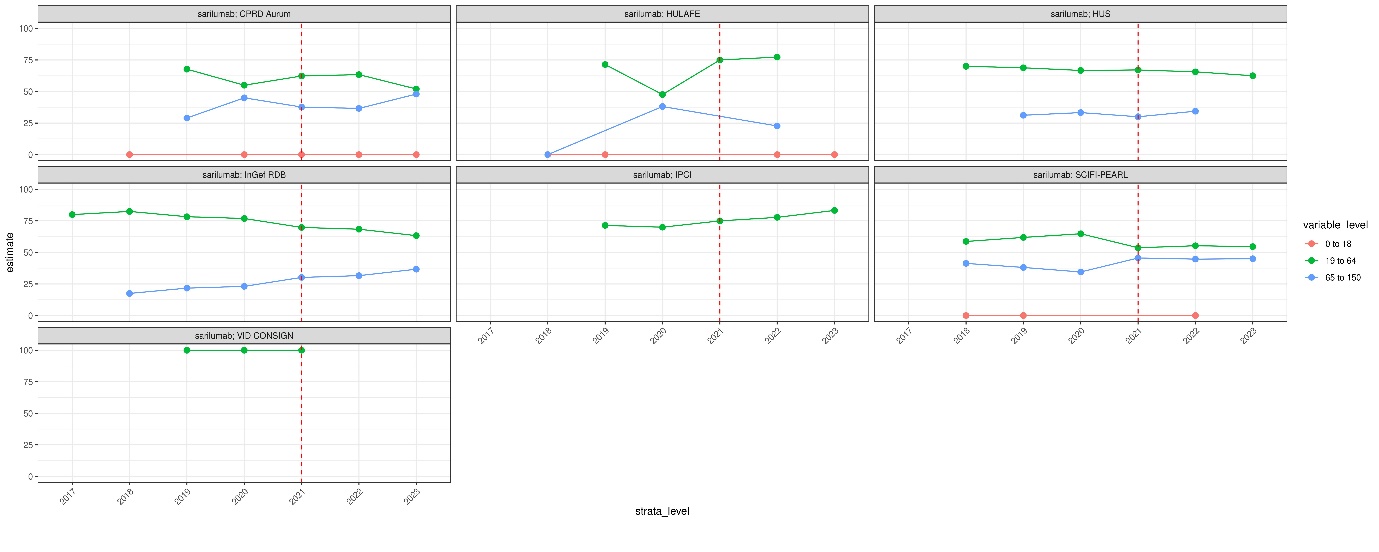

C) Observed changes in proportion of sex of incident users after the shortage date

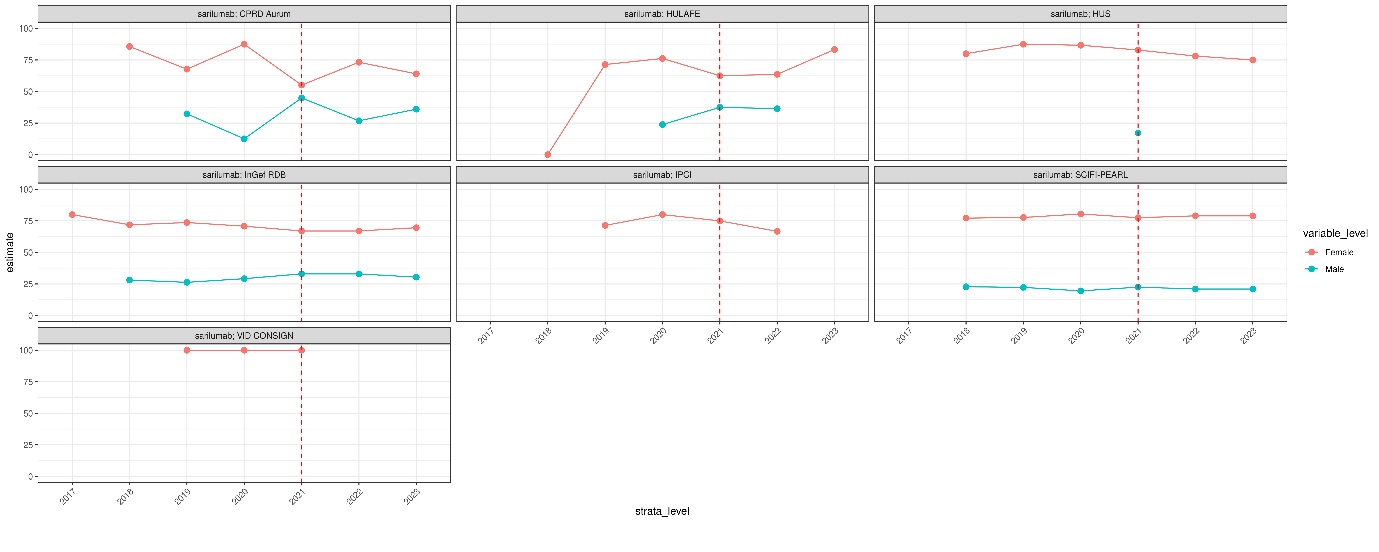

D) Observed changes in indication for rheumatoid arthritis of incident users after the shortage date

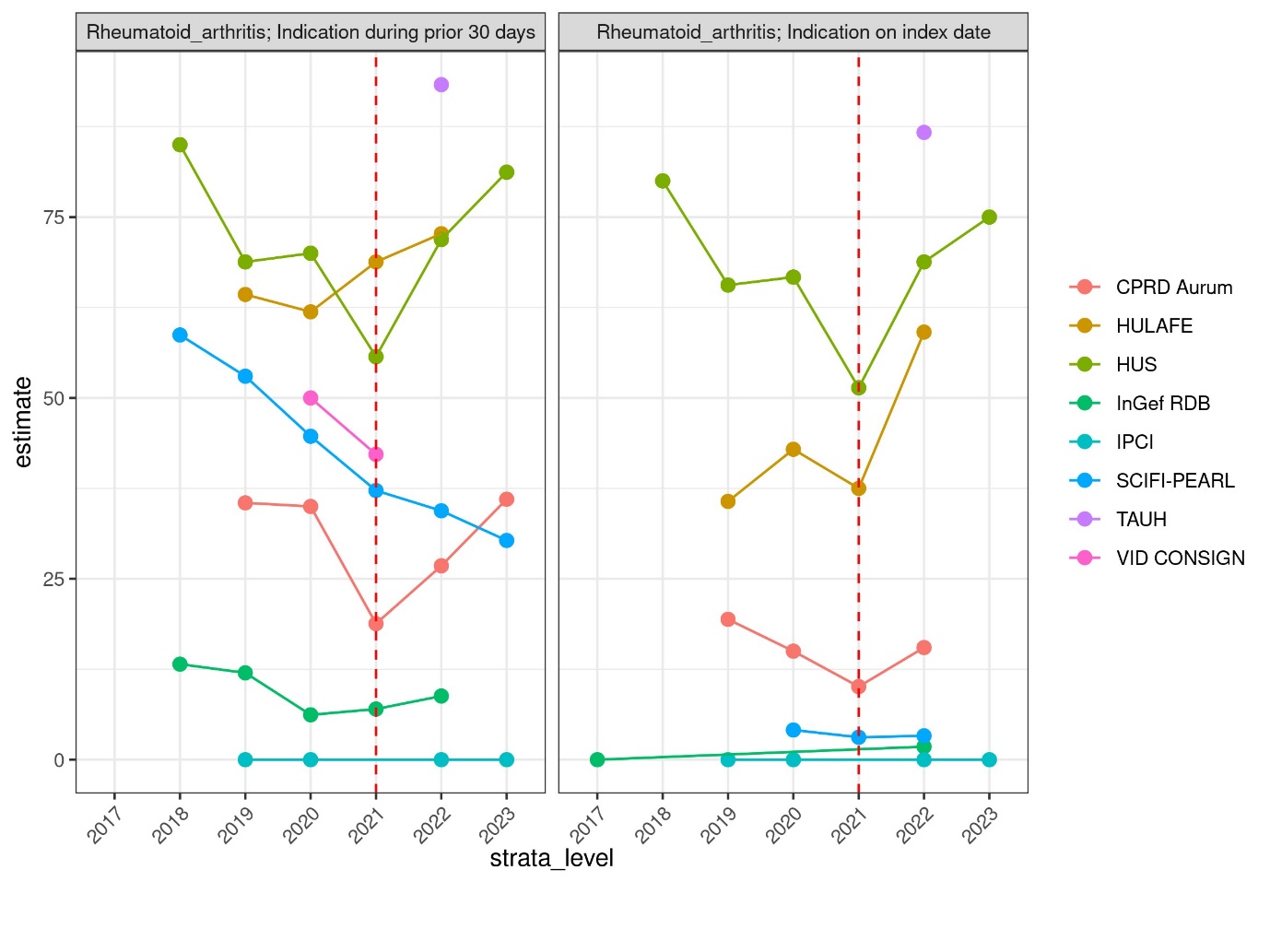

E) Observed changes in duration of incident users after the shortage date

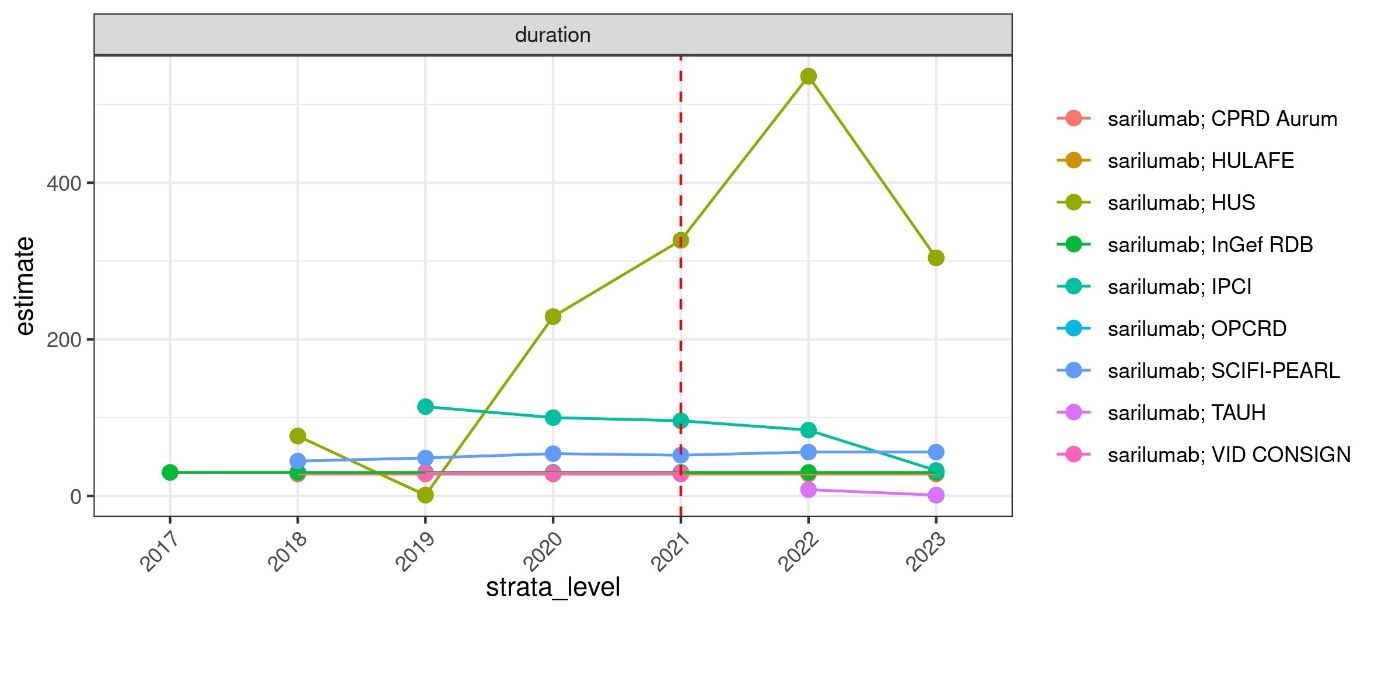

F) Observed changes in initial quantity of incident users after the shortage date

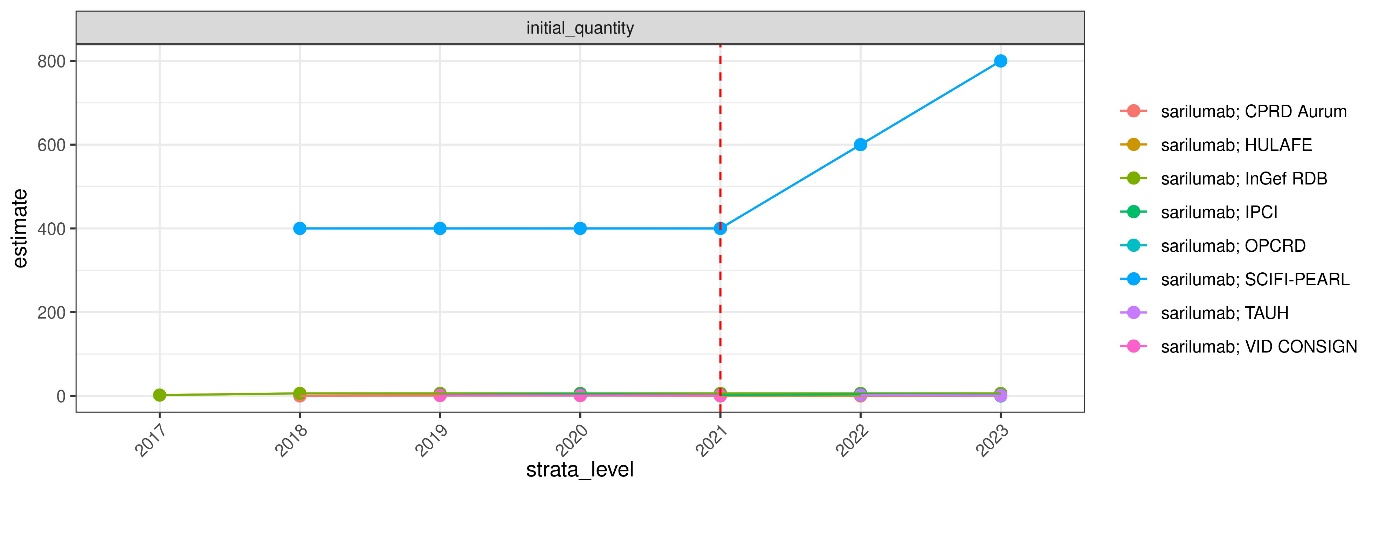

G) Observed changes in cumulative dose of incident users after the shortage date

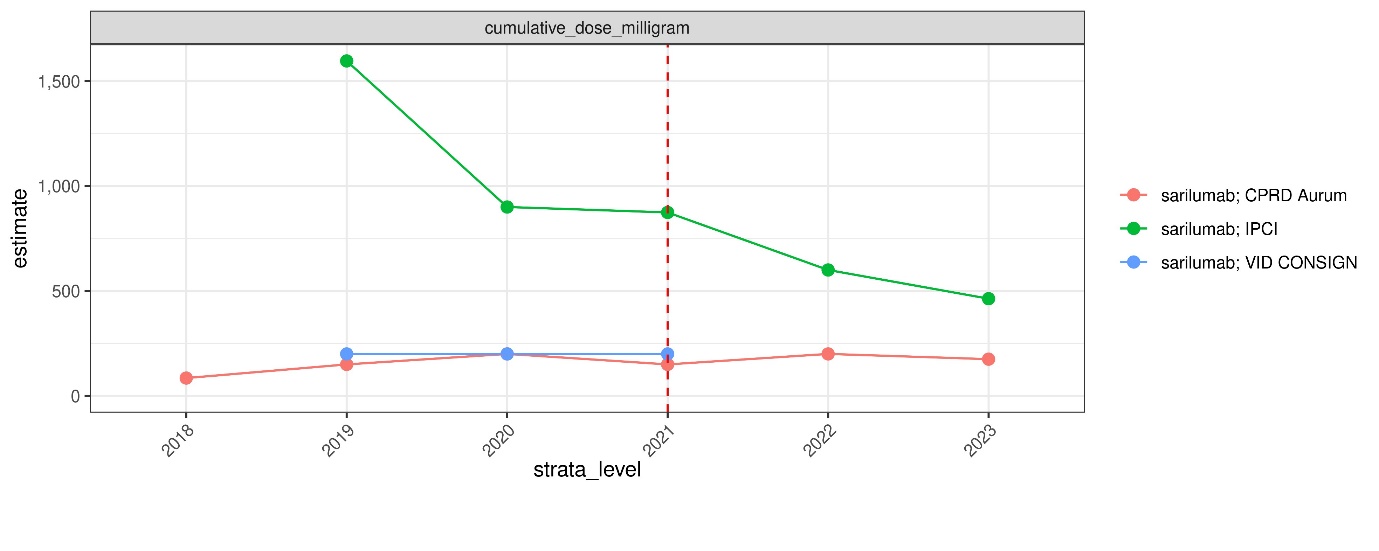

**Supplementary Figure 7. Observed changes in large-scale patient-level drug utilisation study of sarilumab: Observed changes among incident users after the shortage date in A) the median age, B) age groups, C) proportion of sex, D) indication for rheumatoid arthritis, E) duration of sarilumab, F) initial quantity, and G) cumulative dose.** The red dotted line indicates the year of shortage of each drug of interest.
