## Supplementary material for "Trends of use of drugs with suggested shortages and their alternatives across 52 real world data sources and 18 countries in Europe and North America": Annex I

Short descriptions of each database

There is a total of 52 databases in the manuscript:

ULSRA:

Unidade Local de Saúde da Região de Aveiro (ULSRA), formerly named ULSBV.

ULSRA is part of Egas Moniz Health Alliance.

ULSEDV:

Unidade Local de Saúde de Entre Douro e Vouga (ULSEDV)

ULSEDV is part of Egas Moniz Health Alliance.

ULSGE:

Unidade Local de Saúde de Gaia/Espinho (ULSGE)

ULSGE is part of Egas Moniz Health Alliance.

L’Assistance Publique - Hôpitaux de Marseille (AP-HM, France) CDM-APHM

The Assistance Publique – Hôpitaux de Marseille (AP-HM) is a public university hospital system with 4 hospitals, 3,400 beds, and more than 12,000 health care professionals. The AP-HM is one the largest health centers in France (after Paris and Lyon). For adults and children, the AP-HM provide hospital care services going from primary care to secondary ones including specialized ad innovative treatments.

The database contains information concerning inpatient and outpatient visits.

Approximately 300,000 hospitalizations (including ambulatory care) are recorded every year at the AP-HM, involving approximately 210,000 patients. Our information system includes multiple data sources with electronic medical record (Axigate), treatment prescription and dispensings (Pharma), oncology treatment (Chimio), Biology and codifications of medical acts and diagnosis for hospitalized patients (PMSI (Programme de Médicalisation des Systèmes d’Information).

The PMSI is the French medico-administrative database restricted to hospitalized patients that contains data (diagnsosis, acts) that allow to group into diagnosis related-groups (DRG). All the stays are coded using the International Classification of Disease (ICD-10th version). Only ICD-10^th^ version codes were used in this study. All these data are collected and stored for more than 10 years with more than 1 billion pieces of data.

Data involved in our CDM start from 2014.

COMNET AOU Policlinico (Modena, Italy)

The Modena Cancer Center (Centro Oncologico Modenese - COM) of the AOU Policlinico (Modena, Italy) annually takes care of several thousand patients affected by solid tumors. All patients are followed within specific diagnostic-therapeutic paths for the pathology. All histopathological data, radiological staging documents (US, CT scan, MRI, PET/SPECT), all therapies (chemo, immunotherapies, radiotherapy, target therapies, hormonal therapies) and survival outcomes are stored in an electronic medical platform (COMnet). This platform is also integrated with laboratory assessments (heamatological. biochemical, microbiological, serological). The COMNet patients records cover nearly the 100% of the tumor cases in Modena Province and the 15% in the Emilia Romagna Region

Hospital del Mar Electronic Health Record Information System (IMASIS, Spain)

IMASIS is the Electronic Health Record (EHR) system of the Parc Salut Mar Barcelona (PSMar), which is a complete healthcare services organization including two general hospitals, one mental health care centre and one social-healthcare centre, which are offering specific and different services in the Barcelona city area (Spain). IMASIS includes clinical information since 1990 and from different healthcare settings such as inpatients, outpatients, emergency room and major ambulatory surgery. Drugs includes information from administrations, dispensations and prescriptions. IMASIS is linked to several other sources of information such as the Spanish National Mortality Database, the Catalan prescription outpatient's database and the Hospital del Mar Cancer Registry. Currently, the database contains hospital-based information on more than 1.7 million patients, and it is included in the Real-World Data Catalogue of the European Medicines Agency (<https://catalogues.ema.europa.eu/node/1022>).

POLIMI (Foundation IRCCS Ca' Granda Ospedale Maggiore Policlinico):

Foundation IRCCS Ca' Granda Ospedale Maggiore Policlinico, known simply as Policlinico of Milan, is a general hospital that can count on important excellence in different areas of care with a strong interdisciplinary focus. Given its nature as IRCCS – Institute for Research, Hospitalization and Health Care - in addition to care, it carries out biomedical and health research activities of a clinical and translational nature, involving the rapid transfer of therapies from the laboratories to the bedside of the sick person. The research activity is conducted in the different fields of medicine, from neurology to cardiology, from transplantation to hematology, to excellence of care in gynecology, neonatology, geriatrics and rare diseases.

Our DWH was born a few years ago with the aim of helping researchers in identifying patient cohorts and in obtaining large amounts of data for their studies more easily. A few years later, thanks to the EHDEN Project, we were also able to introduce the CDM OMOP.

Currently the DWH contains data from Hospitalization, Outpatients visits, Laboratory analysis, Therapies, Radiology, Anatomic Pathology and a REDCap instance for non-profit studies.

AOUPR (Hospital de Parma, Italy):

The University Hospital of Parma (AOU), together with the Local Health Unit Company (AUSL), is a highly specialized multi-specialist academic hospital, providing the full repertoire of diagnostic, therapeutic and rehabilitation services to the provinces of Parma, Piacenza and Reggio Emilia, is the Trauma and Neurosurgical Reference Center for north-western Emilia. As part of its scientific research activities, AOU carries out basic, translational, clinical and epidemiological research in a wide range of disciplines, also thanks to the close collaboration with the University of Parma. The clinical data warehouse (CDW) has been recently harmonised to OMOP CDM and currently includes data on admissions, clinic, day hospital and day surgery, diagnoses and interventions (surgery, rehabilitation, drug delivery), laboratory data, birth events, cardiac surgery interventions, blood transfusions, cancer and mortality registries.

ATENEA_OMOP (Denia, Spain):

The Health Department of Denia belongs to the Health Care System of the Region of Valencia and provides health services (from primary to tertiary care) to 150,000 inhabitants, reaching 300.000 during summer season. The health department includes a Hospital and 20 primary care centres, that work as an integrated network, supported by an unified EHR system powered by Oracle’s MILLENNIUM. Its level of integration, including both processes and data, has been recognized by achieving the HIMMS Stage 6 in the eight-stage (0-7) EMRAM maturity model. Starting from the central health database the hospital composed a data warehouse composed by different datamarts created for specific purposes. One of these has been created for the OMOP data model in the context of the EHDEN Project. It includes data starting from 2009 onwards and receives periodic updates, the last performed in 2024. It includes information about all the services offered by the Hospital including Emergencies, In and Out patient's activity, medication and drugs, surgery, oncology, paediatrics, etc.

AZGR:

AZ Groeninge is a progressive supraregional hospital located in Kortrijk ([www.azgroeninge.be](http://www.azgroeninge.be)) in Belgium. In 2021, staff provided approximately 470.000 consultations, 36.700 overnight hospitalizations, 75.000 ambulant hospitalizations, 45.400 emergencies and 45.000 procedures. In addition to expert and warm patient care, research and development opportunities are endorsed within the organization, with particular focus on (1) Sustainable healthcare; (2) Technological developments and (3) Clinical research studies. The AZ Groeninge database is a dataset for selected medical conditions extracted from the electronic patient record, including non-structured (textual) data, for which a context specific NLP algorithm is used, and related auxiliary data sources holding all medical, paramedical and nursing data available on single patient level. For each clinical context, a specific set of clinically relevant datapoints are mapped to OMOP-CDM. In particular, data is currently available for the following clinical populations: immuno-oncology, breast oncology, lung cancer and cardiology (particular focus on hearth failure and atrial fibrillation).

BARDENA:

Bardena is a data infrastructure that gathers periodically data from the Health Care System of the Region of Navarra that provides health services (from primary to tertiary care) to 660,000 inhabitants directly.

The data warehouse covers information from Primary and Specialized Care (in and outpatient) as well as Emergencies, including visits, diagnostics and treatments, procedures, vaccines, Drug prescriptions and Laboratory among others. It included 1.97 millions pseudonymized patients.

Data is from 2012 onwards and different tables are updated daily and weekly. Pharmacy data is available from 2012 onwards and contains drugs which are prescribed or administered in the hospital or primary care centers. All the data is also standardised into the OMOP CDM format.

BDR (Bulgarian Diabetes Register, Sofia, Bulgaria):

All the information needed for the Register is extracted from the outpatient records, collected by the Bulgarian National Insurance Fund (NHIF). In Bulgaria the outpatient records are produced by the General Practitioners (GPs) and the Specialists from Ambulatory Care for every contact with the patient. The outpatient records are semi-structured files with predefined XML-format.

The most important indicators like Age, Gender, Location, Diagnoses are stored in explicit tags. The Case history is presented in the Anamnesis. Patient status includes a summary of the patient state, symptoms, syndromes, patients’ height and weight, body mass index (BMI), blood pressure and other clinical descriptions.

The values of clinical tests and lab data are enumerated as free text in another section. A special section is dedicated to the prescribed treatment, where treatments reimbursed by NHIF are structured and mapped.

The data repository contains more than 1’600’000 pseudonymised outpatient records for more than 502’000 patients with diabetes.

Bnai Zion Medical Center (BZMC)

The Bnai Zion Medical Center's database is a comprehensive electronic medical record (EMR) system for a large-scale medical facility. Located in Haifa, a city in northern Israel, Bnai Zion Medical Center is a 474-bed hospital serving a population of 500,000 people. Annually, the center handles approximately 280,000 visits, including 50,000 emergency department visits and 200,000 outpatient visits. The facility performs about 14,000 surgical procedures and manages around 3,500 births each year. The medical center employs a workforce of 1,800 staff members. The patient population is insured by the country's four health maintenance organizations (HMOs). Bnai Zion Medical Center provides a wide range of comprehensive inpatient and outpatient services, including emergency room care, operating room services, elective and emergency hospitalizations, imaging, and various specialist consultations.

Charité Cancer (CHA CAN, Berlin, Germany):

The Charité Berlin provides the dataset `CHA CAN` that consists exclusively of clinical data from oncology. It originates from the integrative tumor center "Charité Comprehensive Cancer Center (CCCC)" and includes approximately 215.000 pseudonymized patients diagnosed with various types of cancer. The data covers patient demographics, conditions, procedures, medications and observations over a period of roughly three decades. The source data is mapped from semi-structured records of a third-party, closed-source documentation software.

CHCZ:

Clinical Hospital Center Zvezdara is one of the largest tertiary health care facilities in Belgrade, Serbia, and covers almost all fields of medicine. The database contains medical health records from 2014 onward.

Clinical Practice Research Datalink (CPRD) GOLD (UK):

CPRD GOLD comprises de-identified primary care data for more than 11 million patients in around 700 GP practices and is collected electronically by the National Health System (NHS) as part of their care and support. GPs, gatekeepers to healthcare within the NHS, record information on demographics, prescriptions, laboratory values, diagnoses including those that were made in hospital or outpatient consultation (and fed back to the GP) as well as medical symptoms and lifestyle factors. Patients with CPRD GOLD are representative of the UK population.

Clinical Practice Research Datalink (CPRD) Aurum (UK):

CPRD Aurum comprises de-identified primary care data for 44.9 million patients in over 1700 GP practices in England and is collected electronically by the National Health System (NHS) as part of their care and support. GPs, gatekeepers to healthcare within the NHS, record information on demographics, prescriptions issued in primary care, laboratory values, diagnoses (including those that were made in hospital or outpatient consultation and fed back to the GP) as well as medical symptoms and lifestyle factors. Only coded information is included (free text is not included). Patients with CPRD are representative of the UK population. Coverage period: Jan 1995- May 2023.

Ref: Wolf A, Dedman D, Campbell J, Booth H, Lunn D, Chapman J, Myles P. Data resource profile: Clinical Practice Research Datalink (CPRD) Aurum. Int J Epidemiol. 2019 <https://doi.org/10.1093/ije/dyz034>

Galilee Medical Center (GMC)

Galilee Medical Center (GMC) is a 732-bed facility located in northern Israel. It is the sole medical center in its region, serving a diverse population of approximately 650,000 individuals. The center provides extensive inpatient and outpatient services, including emergency room care, operating room services, elective and emergency hospitalizations, imaging, and specialist consultations. The patient population is ethnically diverse, comprising Ashkenazi Jews, Sephardic Jews, Muslims, Christians, and others, and is insured by the country's four health maintenance organizations (HMOs). GMC maintains a comprehensive, nonselective electronic medical record (EMR) database, capturing a complete range of clinical information.

HULAFE (Valencia, Spain):

The Health Department Valencia-La Fe belongs to the Health Care System of the Region of Valencia and provides health services (from primary to tertiary care) to 300,000 inhabitants directly. The Department has a central unit located at La Fe Hospital and has deployed a de-identified EMR that has achieved stage 6 in the eight-stage (0-7) EMRAM maturity model. The data warehouse is composed of the aggregation of 22 datamarts, comprising data from different origins (emergency care settings, outpatient, hospitalization, clinical reports, primary care, surgical unit, intensive care, hospital-at-home care, casuistry attended, etc) for 2.5 million patients, where 300.000 patients belong to La Fe Hospital.

Data is from 2012 onwards and different tables are updated daily and weekly. Pharmacy data is available from 2016 onwards and contains drugs which are prescribed or administered in the hospital or primary care centers. Data comes from La Fe Hospital, one specialty centre and 20 primary care centers. All the data is also standardised into the OMOP CDM format.

### HUS (Helsinki University Hospital, Finland)

HUS is the biggest health care provider in Finland providing specialist and secondary care for a catchment area of about 2.2 million people (<https://www.hus.fi/en/about-us>). As a university hospital, HUS continuously develops and evaluates treatment methods and activities. HUS is responsible for organizing specialized healthcare in the Uusimaa region. In addition, the treatment of many rare and severe diseases is nationally centralized to HUS. All patients who visit the HUS hospitals are recorded in the HUS IT system (EPIC). All visits, all procedures, and treatments, including drug prescriptions and administrations are recorded systematically in electronic format (<https://www.hus.fi/en/research-and-education/tutkijan-palvelut/data-services>). HUS uses more than a hundred different operational IT systems.

For secondary use, the individual level data is pooled into a data lake. Data relevant for research is then collected through an ETL-process and provided to dedicated secure analytics environments for a specific research purpose (project).2 Certifications: ISO 13485:2016, ISO 9001:2015, EHDEN Data Partner.

The HUS OMOP database contains granular data from 3.1 million patients.

INGEF FDB (PROD) [Name recorded in EHDEN Portal – InGef RDB]:

InGef RDB contains administrative healthcare claims data from over 50 German statutory health insurances (SHIs) from all federal states of Germany. Data available in the InGef Research Database include partly coarsened information on demographics (quarter of birth, sex, quarter of death if applicable and region of residence on administrative district level) as well as diagnoses and operations in the inpatient setting and diagnoses and reimbursed drugs in the outpatient setting. In addition, costs from the SHI perspective are available for all healthcare sectors. Lag time of complete data availability is about nine months.

Notes relevant to this study:

- Data is included from 2013.01.01 until 2023.03.31
- Since outpatient diagnoses are only recorded on a quarterly basis, they are mapped over non-standard concepts in the OBSERVATION table. Therefore, all analyses of diagnoses only consider inpatient diagnoses

INT Database (DataWareHouse of the INT – National Cancer Institute,Milano, Italy):

INT is an Italian oncology center, located in the Lombardy region (Northern Italy) and approximately 30% of patients come from other Italian regions.

The institutional Data Warehouse (DWH) was transcoded in OMOP. The DWH is the institutional database where information relating to all the activities that patients carry out at our institution is centralized.

We have about 13500 hospitalizations per year for 460 hospital beds. Lab exams are about 850000 per year. Other outpatient services are about 370000 per year.

As mentioned, we are a mono-specialistic institute dedicated to diagnosing and curing cancer.

So, in our DWH we have cancer patients, but the majority are cases performing diagnostic exams (or other outpatient services) with negative diagnosis of cancer.

In synthesis, the DWH is used to:

- identify potential patients for clinical trials
- retrospectively count the potential number of patients for new prospective studies
- understand the feasibility of retrospective observational studies
- provide data for European and national data sharing studies
- have indications on institutional strategic decisions
- facilitate institutional management control activities

The Integrated Primary Care Information (IPCI, NL)

The IPCI database started in 1992 and is collected from EHR records of patients registered with their GPs throughout the Netherlands. The selection of 551 practices, of which 320 are currently still actively contributing, is representative for the entire country. The database contains records from in total 2.8 million patients (approximately 1.3 million are still active) out of a Dutch population of 17 million. The observation period for a patient is determined by the date of registration at the GP and the date of leave/death. The observation period start date is refined by many quality indicators, e.g. exclusion of peaks of conditions when registering at the GP. All data before the observation period is kept as history data. Drugs are captured as prescription records with product, quantity, dosing directions, strength and indication. The duration of the drug exposure is determined for all drugs by: 1. The amount and dose extracted from the signature or if instruction is "see product instructions" we use the DDD and quantity; 2. Duration available in the record; 3. If option 1 and 2 is not possible we use the DDD derived duration, or default to 30 days otherwise. Drugs not prescribed in the GP setting might be underreported. Indications are available as diagnoses by the GPs and, indirectly, from secondary care providers but the latter might not be complete.

ITF_CDM:

Istanbul University (IU) Istanbul Faculty of Medicine uses a bespoke EHR system (IU- Hospital Information Management System (HBYS)). All IU hospitals' medical records are stored in a central data repository. All doctor visits, diagnostics, treatments, prescriptions, and other relevant information are entered into the record.

Database type: hospital

Number of patients: 899515

CDM version: 5.3.1

The data is sourced from IU Istanbul Faculty of Medicine Hospital

Last CDM database update: Sep 2021

ICD10- Snomed standard mappings used

Lab codes mapped to LOINC codes

Procedure codeds mainly mapped to SNOMED

Drugs barcodes mapped to RxNorm and RxNormext.

Units and Route codes mapped to UCUM

MAITT (University of Tartu, Estonia)

MAITT is a dataset is composed from three national health databases in Estonia: digital prescription, claims, and EHR. The dataset contains information about primary care, secondary care (inpatient and outpatient) visits. It contains 10% random sample from Estonian population (approximately 150 000 persons). For each individual in the sample, it contains all pseudonymised records from these three databases from 2012-2019. Drug information is derived from drug dispensing records. These records contain product info, quantity, dosing directions, strength and indication. Drug exposure length is calculated from quantity and dosing directions or if available using duration on the record. If these are not available, we use the most frequent duration for the ingredient from available durations or 30 days.

UZA (University Hospital Antwerp):

UZA is an academic hospital in the city of Antwerp, Belgium, affiliated with Antwerp University. The dataset included for the EHDEN Mega Study consists of longitudinal structured hospital data of >4500 patients with breast cancer, lung cancer, CLL, MM and treated with immune-checkpoint inhibitors. It includes pharmacy data, minimal hospital data (diagnoses, comorbidity), procedures, devices, admission data, selected lab results, vital parameters, cancer staging, death.

Merative MarketScan® (MarketScan, US):

MarketScan databases (Commercial Claims and Encounters and Medicare Supplemental) capture person-specific clinical utilization, expenditures, and enrolment across inpatient, outpatient, prescription drug, and carve-out services from a selection of large employers, health plans, and government and public organizations in the United States. These medical databases include private sector health data from approximately 100 payers. Historically, more than 500 million claims records are available in the MarketScan databases. These data represent the medical experience of insured employees and their dependents for active employees, early retirees, Consolidated Omnibus Budget Reconciliation Act (COBRA) continues and Medicare-eligible retirees with employer-provided Medicare Supplemental plans. The study used MarketScan databases converted to the OMOP common data model as of October 2022, which included 177 million patients at that time.

Netherlands Cancer Registry (NCR) (IKNL, The Netherlands):

The Netherlands Cancer Registry is a national registration providing statistics on cancer in The Netherlands. The registry is maintained by the Netherlands Comprehensive Cancer Organisation (IKNL). The recording of data in this database is performed by registration employees of IKNL. The Netherlands Cancer Registry is the only oncological hospital registry in The Netherlands with data on all cancer patients. Data are available on national level from 1989 onwards. The data from 1993 on has been converted into the OMOP database and the OMOP database is restricted to patients 18 years or older at time of diagnosis. The Netherlands Cancer Registry comprises information on newly diagnosed cancer patients in the Netherlands, including cancer diagnosis, tumour staging (according to the TNM-classification developed and maintained by the Union for International Cancer Control (UICC)), tumour site (topography) and morphology (histology) (according to the WHO International Classification of Diseases for Oncology (ICD-O-3)), and treatment received directly after diagnosis. Death records are linked once a year using the municipality registration. The most recent linkage of death records was performed in January 2024.
Reference: [Netherlands Cancer Registry (NCR) (iknl.nl)](https://iknl.nl/en/ncr)

OLVA MS:

The OLV hospital is a general hospital, located in Aalst, Asse and Ninove, in Belgium. It is one of the largest non-university hospitals in the Flemish region with over 800 accredited hospital beds. This database contains structured pharmacy data from 2016-2024 and minimal hospital data, including procedures and diagnosis from 2016 to 2023. In total 51.262 patients are included.

Hospital 12 de Octubre – H12O

The 12 de Octubre Hospital, a high complexity center, is the specialized health center of reference for approximately 450,000 patients of the southern area of Madrid. It is one of the largest hospitals in Spain in terms of surface area and number of beds and can solve most of the patients' health problems, since it has practically all the medical and surgical specialties, and central services with state-of-the-art technological equipment, HIMSS 6. The hospital has developed the INFOBANCO project, a platform for the reuse of clinical data from the EHR based on standards such as OpenEHR, OMOP, i2b2 and terminologies such as SNOMED or LOINC. Thus, this platform has information from more than one million patients on their diagnoses, procedures, interventions, outpatient visits, emergencies, admissions, medication and laboratory.

Optimum Patient Care Research Database (OPCRD):

OPCRD collects anonymised coded data from over 1,200 general practices in England, Scotland and Wales using all the three major clinical management systems. Data for OPCRD is collected monthly from contributing practices. OPCRD includes data for more than 25 million individuals, of whom over 10 million are currently registered patients. It is representative of the UK population in terms of age, sex, ethnicity and socio-economic status. The coded data collected includes diagnoses, prescriptions, immunisations, and basic demographic characteristics. The OPC Collaborative Clinical Network (OPC) has grown to become a global leader in the provision of technologically enhanced health care data and clinical research services and the network continues to grow month-on-month. OPCRD has been converted to the OMOP CDM, enhancing its interoperability with other health databases and facilitating more extensive and robust clinical research.

Optum's de-identified Clinformatics® Data Mart Database (Clinformatics ®, US)

Optum's de-identified Clinformatics® Data Mart Database is a database comprised of administrative health claims for members of a large national managed care company in the United States. These administrative claims are submitted by providers and pharmacies for payment and are verified, adjudicated, adjusted, and de-identified prior to inclusion in Clinformatics®. In addition to medical claims, pharmacy claims, and lab results, Clinformatics® includes data tables related to member inpatient confinements and member eligibility data. The study used the Clinformatics® as of December 2022, converted to the OMOP common data model, which included 77.2 million patients at that time.

Papageorgiou General Hospital (PGH)

The Papageorgiou General Hospital (PGH) database contains healthcare data for about 1.5 million patients. It contains clinical, administrative and lab exams data from the year 2000 on and it covers the northern Greece/central Macedonia area as PGH is one of the biggest public hospitals in this region. PGH database includes data from patients with a wide range of conditions (oncological, pathological, surgical etc.)

IQVIA Pharmetrics+:

PharMetrics is the largest non-payer-owned integrated claims database of commercial insurers in the US. This de-identified, integrated database includes all paid medical and pharmacy claims for more than 70 million members from more than 100 health plans across the US. It includes both inpatient and outpatient claims, diagnoses and procedures based on ICD and Current Procedural Terminology codes, as well as retail and mail order pharmacy claims.

PRISIB:

This database is the result of the harmonization and merging of different data sources across the Public Health System of the Balearic Islands (IBSalut) including data from 7 hospitals, 61 primary care areas and around 1,300,000 active patients, which encompass the whole of the population of the archipelago. Hospital data includes diagnostic and procedure codes from the discharge reports. Primary care data includes visits, measurements, diagnoses, procedures, prescriptions and laboratory results.

Reuma.pt:

Reuma.pt (Rheumatic Diseases Portuguese Registry) is an observational, prospective, long-term registry of rheumatic patients residing in Portugal. Reuma.pt is a project from the Portuguese Society of Rheumatology that has been active since 2008. It includes the semi-structured clinical data of more than 35,000 patients, and 300,000 visits. The registry functions as an electronic health record and has protocols that permit the registration of 14 rheumatic diseases (rheumatoid arthritis, psoriatic arthritis, juvenile idiopathic arthritis, spondylarthritis, systemic lupus erythematosus, auto-inflammatory syndromes, osteoporosis, early arthritis, vasculitis, osteoarthritis, systemic sclerosis, myositis, Sjögren’s syndrome, mixed connective tissue disease). Data includes diagnoses, demographics, procedures, prescriptions, and laboratory results.

SCIFI-PEARL

The SCIFI-PEARL (Swedish Covid-19 Investigation for Future Insights – a Population Epidemiology Approach using Register Linkage) project is a nationwide linked multi-register, regularly updated, observational study for timely response over time to scientific questions regarding SARS-Cov-2 (severe acute respiratory syndrome coronavirus 2) and the COVID-19 (Coronavirus disease 2019) pandemic, with broad scientific aims that cover diverse aspects of COVID-19 and vaccination as well as the pandemic impact on society, healthcare, drug use and availability. Data span the years 2015 (2018 for prescription drug data) to currently 2023.
Through its coverage of the entire Swedish population, the SCIFI-PEARL database captures all Swedish residents with regularly updated data (approximately quarter-yearly). It links to a broad range of national and regional healthcare data for a comprehensive longitudinal view of the population and patient experience before, during and after the pandemic.

The Information System for Research in Primary Care (SIDIAP)

SIDIAP is a database of population-wide primary care electronic health records of the population of Catalonia, North-East Spain. The database contains pseudo-anonymized records for >8 million people since 2006, with 5.8 million people active in June 2021 (75% of the Catalan population). SIDIAP is representative of the general population living in Catalonia in terms of age, sex and geographic distribution.

Parc Sanitari Sant Joan de Déu - PSSJD (Spain)

Parc Sanitari Sant Joan de Déu is a comprehensive healthcare institution located in Barcelona, Spain, specializing in mental health and social care services. The centre serves a population of approximately 1.6 million people in the metropolitan area and provides a wide range of services, including inpatient and outpatient mental health care, social care for vulnerable populations, and community-based programs. The institution operates a fully integrated health information system that supports clinical, administrative, and research activities, ensuring a high level of coordination across its various services. Parc Sanitari Sant Joan de Déu is involved in multiple research initiatives, particularly in the field of mental health, and contributes to various regional and international projects. The data warehouse at Parc Sanitari includes comprehensive information covering mental health assessments, inpatient admissions, outpatient visits, medication records, and social care activities.

SUCD:

The Semmelweis University (SU) Clinical Database has medical records data of inpatient and outpatient visits of all hospitals affiliated with the SU in Budapest, Hungary starting from 2011. It includes diagnoses, procedures, drugs and laboratory test results in a structured form and also all healthcare-related free text-based data. As we have a National Health Insurance System, it also serves as a basis of claims.

MEB KI:

The MEB-KI dataset is a nationwide, multi-register resource encompassing the entire population of individuals aged 65 and older in Sweden. It includes data from 2006 onwards, with approximately bi-annual updates, covering a cumulative sample size of around 3.1 million individuals. The core of the database is formed by linked data from several key registers: the Total Population Register, the National Patient Register, the National Prescribed Drug Register, the National Social Services Register, and sociodemographic data from Statistics Sweden.

TAUH (Tampere University Hospital):

Tampere University Hospital (TAUH) is the hospital of the Wellbeing Services County of Pirkanmaa, the second biggest public healthcare provider in Finland. TAUH provides expertise in nearly all medical specialties and serves a population base of 0.5 million.

The TAUH database covers the specialized healthcare of all age and social groups in the region. The TAUH OMOP database has data for 893,000 patients, and contains patient demographics, inpatient and outpatient visits, diagnoses, hospital-administered drugs, electronic prescriptions, surgeries, imaging and other procedures, laboratory measurements, and pathology diagnoses.

THIN®

THIN® (The Health Improvement Network) is Cegedim Health Data’s European medical databases network of electronic health records. These data are transmitted by a network of voluntary physicians who firmly believe that supporting this kind of longitudinal data observatory benefits research and medical progress. THIN® currently covers over 72 million electronic health records across several European countries: the UK, France, Germany, Italy, Spain, Belgium and Romania. Its data is available to all life science stakeholders. They are used by leading healthcare authorities and research centers, and also by academics for numerous scientific publications. THIN® therefore contributes to advancements in patient care and outcomes, in the interests of public health, in compliance with current regulations, including GDPR.

THIN® includes information on demographics, prescriptions, laboratory values, symptoms, diagnoses, including those that were made in secondary care (and fed back to the GP). There is an average history of more than seven years.

THIN® is built on a common data model (applicable in each country), which is mapped to the OMOP format in FR, UK, RO, IT, SP, and BE, which are part of this study.

UM Dresden:

The Data Integration Center (DIC) at Dresden University Hospital is part of the Center for Medical Informatics at Dresden University Hospital. The DIC currently provides access to all in-patient and outpatient visits with its conditions, procedures, laboratory findings, patient demographics (gender, age and birthdate) and medications (limited to the in-patient medication prescriptions and administrations). Our data set in the DIC contains around 58.000 in-patient cases and 250.000 outpatient cases per year. We have 1.3 Mio patients in the data source. A subset of the data in the DIC is provided as FHIR resources based on the core data set basic modules defined by the German Medical Informatics Initiative (https://www.medizininformatik-initiative.de/en/basic-modules-mii-core-data-set). These FHIR resources are transformed to OMOP CDM with the restriction of visits from 2018.

PCASF

The Prostate Cancer Registry of South-west Finland includes all men with suspicion and established diagnosis of prostate cancer in South-West Finland. As one tenth of the population in Finland resides in South-West Finland, yearly some 1500 men are entered to the database including 500 men with prostate cancer diagnosis. The database includes all data from EHRs in specialized health care.

VID_CONSIGN:

The Valencia Health System Integrated Database (VID) is a set of multiple, public, population-wide electronic databases for the Valencia Region, the fourth most populated Spanish region, with about 5 million inhabitants and an annual birth cohort of 48 000 newborns, representing 10.7% of the Spanish population and around 1% of the European population. The VID provides exhaustive longitudinal information including sociodemographic and administrative data (sex, age, nationality, etc.), clinical (diagnoses, procedures, diagnostic tests, imaging, etc.), pharmaceutical (prescription, dispensing) and healthcare utilization data from hospital care, emergency departments, specialized care (including mental and obstetrics care), primary care and other public health services. It also includes a set of associated population databases and registries of significant care areas such as cancer, rare diseases, vaccines, congenital anomalies, microbiology (including COVID-19 test results registry) and others, and also public health databases from the population screening programmes. All the information in the VID databases can be linked at the individual level through a single personal identification code. The databases were initiated at different moments in time, but all in all the VID provides comprehensive individual-level data fed by all the databases from 2008 to date. FISABIO-HSRP is not the owner of the VID data, and access to the data is project-dependent. Therefore, the OMOP instances are created on a project-by-project basis. The VID-CONSIGN OMOP instance described here has been created using CONSIGN project data, where 1.96 Million of females in fertile age are studied from start of 2018 to end of 2021.

Viecuri:

VieCuri Medical Center, Rheumatology Database (The Netherlands):

VieCuri Medical Center, is a general Dutch hospital located in Venlo, specializes in rheumatology among other areas and holds STZ certification for high-quality care.

The Rheumatology Database includes clinical data for 4,900 patients treated from January 2018 to August 2023. The database has been designed to support clinical research and to contribute to broader epidemiological studies across Europe and will be extended in the following years to include more patients and specializations.

Key features of the database are:

- Patient Demographics: Age, gender, and region of residence.
- Clinical Diagnoses: Focus on rheumatoid arthritis (RA), psoriatic arthritis (PsA), and gout.
- Treatment Data: Information on prescribed medications (biologics, DMARDs, NSAIDs).
- Laboratory Results: Data on biomarkers such as CRP, ESR, rheumatoid factor, and serum urate levels.
- Outcomes: Measures of patient progress using DAS28.
- Follow-up Records: Data on follow-up visits and chronic condition management.

ULSM:

Unidade Local de Saúde de Matosinhos (ULSM) is the oldest healthcare cluster in Portugal that provides all levels of care to a well-defined region in Portugal. The database starts in 1999 ULSM is part of Egas Moniz Health Alliance.
